## Supplementary material for "Machine Learning Models for Predicting Stroke Risk Among Patients with Coronary Heart Disease": Latex Files: Manuscript.pdf

### Materials and methods

#### Data Source and Preprocessing

This study employed secondary data on coronary heart disease obtained from the IEEE DataPort repository, one of the most well-known repositories that has made high-quality datasets available for cardiovascular and epidemiological studies [33]. The dataset is based on the Behavioral Risk Factor Surveillance System survey and comprises responses from 253,680 people. It consists of 22 variables, including demographic, lifestyle, behavioral, clinical, healthcare access, and overall health status variables. Among the total respondents, 248,960 participants had no history of Stroke, whereas 4,720 participants reported a positive Stroke status. Due to the large population and the variety of risk factors it covers, which are most relevant to Stroke prediction, this study is most relevant to the prediction of Stroke. Even though the data was gathered in the United States, the risk factors that were included, including smoking habit, cholesterol, high blood pressure, and diabetes, portray Stroke determinants that are also equally common in most of the low- and middle-income environments. This renders the valuable dataset useful for training generalized prediction models, which can later be modified and tested with smaller regional datasets to improve their applicability in local settings.

To model the data, a systematic preprocessing process was used to ensure data quality and enhance model learning. The missing values were addressed using appropriate techniques, such as multiple imputation or a k-nearest neighbors approach, based on the nature of the missingness and its distribution. Outlier detection has been performed using methods such as the interquartile range and the Mahalanobis distance. Noise reduction used smoothing procedures, feature correlation analysis, and low variance attribute elimination. Moreover, scaling methods such as normalization and standardization were applied to align feature ranges and enable stable, efficient training.

#### Addressing Class Imbalance

Given the significant class imbalance in the dataset (with stroke cases representing only approximately 1.86% of the total sample), we employed the Synthetic Minority Over-sampling Technique (SMOTE) to generate synthetic samples for the minority class. This approach helps prevent model bias toward the majority class and improves the detection of rare but clinically important stroke events.

SMOTE operates by creating synthetic examples along line segments connecting minority class instances in feature space. For each minority class instance $x_i$, we:

1. Find its $k$ nearest neighbors from the minority class (typically $k = 5$)
2. Randomly select one of these neighbors, $x_{zi}$
3. Generate a synthetic sample $\tilde{x}_i$ along the line segment between $x_i$ and $x_{zi}$:

$$\tilde{x}_i = x_i + \lambda \cdot (x_{zi} - x_i) \tag{1}$$

where $\lambda$ is a random number uniformly distributed between 0 and 1. This interpolation creates new instances that lie in the feature space region between existing minority class samples.

To ensure robustness and prevent overfitting, we applied SMOTE in conjunction with stratified $k$-fold cross-validation during model training. Specifically, we:

- Used stratified sampling to maintain the original class distribution in training and test splits
- Applied SMOTE only to the training folds during cross-validation, leaving the test folds untouched to provide unbiased performance evaluation
- Generated synthetic samples until the minority class reached 50% of the majority class size, balancing detection sensitivity with computational efficiency

This combined approach of SMOTE with stratified cross-validation ensured that our models could learn meaningful patterns from the minority class while maintaining generalizability to unseen, naturally imbalanced clinical data.

#### Feature Selection

Feature selection is critical for improving model accuracy and interpretability. We applied a hybrid approach combining statistical and machine learning-based methods.First, univariate feature selection based on mutual information $I(X;Y)$ was applied:

$$I(X;Y) = \sum_{x \in X} \sum_{y \in Y} p(x,y) \log \left( \frac{p(x,y)}{p(x)p(y)} \right) \tag{2}$$

This quantifies the dependency between each feature $X$ and the target $Y$. Next, recursive feature elimination with cross-validation (RFECV) was applied using a base classifier to iteratively remove the least important features until optimal performance was achieved.

#### Machine Learning Algorithms

This study implements a multi-model machine learning framework designed to predict stroke risk among individuals diagnosed with coronary heart disease. The framework integrates deep learning architectures with tree–based and kernel–based ensemble methods. A stacked meta–learner is ultimately used to combine model outputs for improved robustness and predictive stability.

#### Deep Learning Components

##### Keras Multilayer Perceptron (Keras-MLP)

A fully connected feedforward neural network was constructed to model nonlinear interactions among clinical and behavioral risk factors.

Let $X \in \mathbb{R}^{n \times d}$ denote the input dataset with $d$ predictors and binary labels $Y \in \{0,1\}^n$. The computations in the MLP are:

$$\hat{y} = \sigma(W_L h_{L-1} + b_L), \qquad h_l = f(W_l h_{l-1} + b_l), \quad l = 1, 2, \ldots, L-1$$

where:

- $h_l$ is the hidden state at layer $l$
- $W_l$ and $b_l$ are trainable weight matrices and biases
- $f(\cdot)$ is the ReLU activation
- $\sigma(\cdot)$ is the sigmoid function for binary classification

##### Keras 1D Convolutional Neural Network (Keras-CNN1D)

Although CNNs are traditionally used for sequential data, they can also extract feature interactions from tabular data through 1D convolutions. Input features are reshaped to a 1-dimensional sequence:

$$z^{(1)} = \text{Conv1D}(X),$$
$$z^{(2)} = \text{ReLU}(z^{(1)}),$$
$$z^{(3)} = \text{GlobalMaxPool}(z^{(2)})$$

$$\hat{y} = \sigma(W z^{(3)} + b)$$

The convolutional filters learn local patterns and interactions that fully connected models may miss.

#### Scikit-Learn MLP Classifier (SKLearn_MLP)

As an additional neural architecture, the Scikit-Learn MLP classifier is included as a lightweight alternative to Keras models. The feedforward computations follow the same formulation as the Keras MLP but without GPU-accelerated optimization.

##### Tree-Based Ensemble Models

###### Random Forest

Random Forest is a bagging ensemble consisting of multiple decision trees. Each tree is trained on a bootstrap sample, and predictions are combined through majority voting:

$$\hat{y}_{RF} = \text{mode}\{\, h_t(x) \,\}_{t=1}^{T} \tag{3}$$

where $h_t(x)$ is the prediction from tree $t$. The model reduces variance and is robust to overfitting. Feature importance is derived from Gini impurity:

$$GI = \sum_{i=1}^{C} p_i(1 - p_i) \tag{4}$$

###### Decision Tree

A single decision tree classifier is used as a simple, interpretable baseline. It partitions the feature space by selecting splits that maximize information gain. For a node with impurity $I$, the information gain is:

$$IG = I_{\text{parent}} - \sum_{k} \frac{N_k}{N} I_k \tag{5}$$

###### XGBoost (Extreme Gradient Boosting)

XGBoost builds trees sequentially, where each new tree attempts to correct errors from previous trees. The model prediction is:

$$\hat{y} = \sum_{m=1}^{M} \eta \, h_m(x) \tag{6}$$

where:

- $h_m$ is the $m$-th boosted tree,
- $\eta$ is the learning rate.

The objective minimized is:

$$L = \sum_{i} l(y_i, \hat{y}_i) + \sum_{m} \Omega(h_m) \tag{7}$$

with a regularization term $\Omega$ controlling tree complexity.

###### LightGBM

LightGBM is a gradient boosting framework that uses leaf-wise tree growth for greater efficiency. It optimizes the same boosting framework as XGBoost but with:

- histogram-based feature binning,
- gradient-based one-side sampling (GOSS),
- exclusive feature bundling (EFB).

The model prediction follows:

$$\hat{y} = \sum_{m=1}^{M} \eta \, h_m(x) \tag{8}$$

#### Kernel-based and Probabilistic Methods

**Support Vector Machine (SVM)**

The SVM classifier maps input data into a high-dimensional space using an Radial Basis Function (RBF) kernel.

The RBF kernel is defined as:

$$K(x_i, x_j) = \exp\big(-\gamma \|x_i - x_j\|^2\big) \tag{9}$$

The decision function is:

$$\hat{y} = \text{sign}\left(\sum_{i=1}^{n} \alpha_i y_i K(x_i, x) + b\right) \tag{10}$$

**Logistic Regression**

Logistic Regression serves as a linear baseline model. The predictive function is:

$$\hat{y} = \sigma(w^T x + b) \tag{11}$$

It estimates the log odds of stroke risk and is valuable for identifying statistically significant predictors.

**Naive Bayes**

The Naive Bayes classifier assumes conditional independence between features. For a set of predictors $x_1, x_2, \ldots, x_d$, the posterior probability is:

$$P(y = 1 \mid x) \propto P(y = 1) \prod_{i=1}^{d} P(x_i \mid y = 1) \tag{12}$$

#### Stacked Ensemble Framework

To leverage the complementary strengths of all models, a stacked ensemble is used as the final estimator. Each base model $k$ produces an output probability $\hat{y}_k$. These are combined as features for a meta-learner (Logistic Regression):

$$\hat{y}_{\text{stack}} = \sigma\Big(\sum_{k=1}^{K} w_k \hat{y}_k^{\mathcal{M}} + b\Big) \tag{13}$$

where:

- $w_k$ are learnable combination weights,
- $K$ is the number of base models.

This approach reduces variance, stabilizes predictions, and strengthens generalization.

#### Model Training and Validation

The dataset was split into training (70%) and testing (30%) sets with stratification to preserve class balance. K-fold cross-validation ($k = 5$) was used during training to tune hyperparameters for all models. Binary cross-entropy loss was used for neural networks:

$$L_{\text{BCE}} = -\frac{1}{n} \sum_{i=1}^{n} \Big[y_i \log(\hat{y}_i) + (1 - y_i) \log(1 - \hat{y}_i)\Big] \tag{14}$$

Ensemble models were optimized using grid search over hyperparameters such as number of trees, learning rate, and maximum depth. Early stopping was applied to prevent overfitting.

#### Model Comparison and Evaluation

To rigorously assess the performance of the proposed hybrid framework, a comprehensive evaluation strategy was implemented, encompassing both standard classification metrics and graphical diagnostic tools. Model comparison was conducted across the deep learning network, individual ensemble models such as Random Forest and XGBoost, and the stacked ensemble, ensuring a robust assessment of predictive accuracy, reliability, and generalization.

**Accuracy** measures the proportion of correct predictions among all predictions and is defined as:

$$\text{Accuracy} = \frac{TP + TN}{TP + TN + FP + FN} \tag{15}$$

where $TP$ and $TN$ are true positives and true negatives, and $FP$ and $FN$ are false positives and false negatives. While accuracy is intuitive, it may not fully capture performance in imbalanced datasets.

**Precision** quantifies the proportion of correctly predicted positive instances among all predicted positives:

$$\text{Precision} = \frac{TP}{TP + FP} \tag{16}$$

**Recall** (sensitivity) measures the proportion of actual positive instances that are correctly identified:

$$\text{Recall} = \frac{TP}{TP + FN} \tag{17}$$

**Specificity** assesses the proportion of actual negative instances correctly identified by the model:

$$\text{Specificity} = \frac{TN}{TN + FP} \tag{18}$$

**F1 Score** is the harmonic mean of precision and recall:

$$\text{F1 Score} = 2 \cdot \frac{\text{Precision} \cdot \text{Recall}}{\text{Precision} + \text{Recall}} \tag{19}$$

**Matthews Correlation Coefficient (MCC)** offers a robust measure for imbalanced datasets:

$$\text{MCC} = \frac{(TP \cdot TN) - (FP \cdot FN)}{\sqrt{(TP + FP)(TP + FN)(TN + FP)(TN + FN)}} \tag{20}$$

The SHAP value $\phi_i$ for feature $i$ is defined as:

$$\phi_i(f, x) = \sum_{S \subseteq N \setminus \{i\}} \frac{|S|!(|N| - |S| - 1)!}{|N|!} [f_x(S \cup \{i\}) - f_x(S)] \tag{24}$$

where:

- $N$ is the set of all features
- $S$ is a subset of features not including $i$
- $f_x(S)$ is the model prediction using only the feature subset $S$
- $|S|!(|N| - |S| - 1)!/|N|!$ is the weighting factor that accounts for different subset sizes

The final model prediction $f(x)$ for an instance $x$ can be expressed as the sum of SHAP values:

$$f(x) = \phi_0 + \sum_{i=1}^{M} \phi_i(x) \tag{25}$$

where $\phi_0$ is the base value (expected model output over the training dataset) and $\phi_i(x)$ is the SHAP value for feature $i$ for instance $x$.

In this study, we computed SHAP values for our best-performing models (LightGBM and XGBoost) to identify global feature importance and provide local explanations for individual predictions. We generated SHAP summary plots to visualize feature importance across the entire dataset and force plots to explain individual predictions.

##### LIME (Local Interpretable Model-agnostic Explanations)

LIME explains individual predictions by approximating the complex model locally with an interpretable model. For a given instance $x$, LIME generates perturbed samples around $x$ and trains a simple, interpretable model (typically linear) on these samples weighted by their proximity to $x$.

The LIME explanation is obtained by solving:

$$\xi(x) = \arg\min_{g \in G} \mathcal{L}(f, g, \pi_x) + \Omega(g) \tag{26}$$

where:

- $g$ is an interpretable model from class $G$ (e.g., linear model)
- $\mathcal{L}$ is a loss function measuring how well $g$ approximates $f$ in the locality defined by $\pi_x$
- $\pi_x(z)$ is a proximity measure between instance $z$ and $x$
- $\Omega(g)$ penalizes the complexity of $g$ to ensure interpretability

In practice, for tabular data, we used the following weighted linear regression:

$$\min_{w} \sum_{z \in Z} \pi_x(z) \left(f(z) - w^T z\right)^2 + \lambda \|w\|_1 \tag{27}$$

where:

- $Z$ is the set of perturbed samples around $x$
- $\pi_x(z) = \exp(-D(x, z)^2/\sigma^2)$ is the exponential kernel with width $\sigma$
- $D(x, z)$ is the distance between $x$ and $z$ (Euclidean distance for continuous features, Hamming distance for categorical features)
- $\lambda$ controls the L1 regularization for sparsity

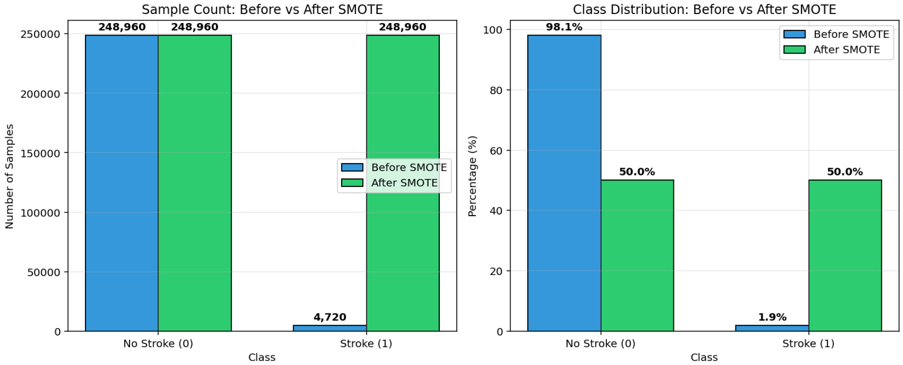


**Fig 1.** Comparison of class distribution before and after applying SMOTE.

#### Predictors of Stroke Among Patients with Coronary Heart Disease

The logistic regression model evaluating determinants of stroke among patients with coronary heart disease showed excellent overall model fit, as indicated by a statistically significant likelihood ratio test ($p < .001$). The model achieved a pseudo-$R^2$ of .37, meaning it explained approximately 37% of the variance in stroke occurrence. Several predictors were statistically significant and clinically meaningful, demonstrating strong associations with stroke risk.

**Table 1. Logistic Regression Predicting Stroke Among Patients with Coronary Heart Disease.**

| Predictor | B (Coef.) | SE | z | p | 95% CI | OR |
|---|---|---|---|---|---|---|
| Constant | 5.325 | 0.250 | 21.334 | < .001 | [4.836, 5.814] | 205.40 |
| Heart attack | 2.448 | 0.109 | 22.390 | < .001 | [2.233, 2.662] | 11.56 |
| High BP | –0.655 | 0.075 | –8.734 | < .001 | [–0.801, –0.508] | 0.52 |
| High cholesterol | –0.713 | 0.070 | –10.233 | < .001 | [–0.850, –0.577] | 0.49 |
| Cholesterol check | –0.646 | 0.077 | –8.423 | < .001 | [–0.796, –0.495] | 0.52 |
| BMI | –0.013 | 0.005 | –2.468 | .014 | [–0.023, –0.003] | 0.99 |
| Smoker | –1.196 | 0.082 | –14.518 | < .001 | [–1.357, –1.035] | 0.30 |
| Diabetes | –0.298 | 0.056 | –5.357 | < .001 | [–0.407, –0.189] | 0.74 |
| Physical activity | –1.408 | 0.064 | –22.151 | < .001 | [–1.532, –1.283] | 0.24 |
| Fruit intake | –1.005 | 0.063 | –16.009 | < .001 | [–1.128, –0.882] | 0.37 |
| Vegetable intake | –1.084 | 0.063 | –17.178 | < .001 | [–1.207, –0.960] | 0.34 |
| Heavy alcohol consumption | –2.202 | 0.182 | –12.098 | < .001 | [–2.558, –1.845] | 0.11 |
| Any healthcare coverage | –0.734 | 0.089 | –8.232 | < .001 | [–0.909, –0.559] | 0.48 |
| No doctor due to cost | –2.044 | 0.139 | –14.660 | < .001 | [–2.318, –1.771] | 0.13 |
| General health rating | –0.134 | 0.023 | –5.720 | < .001 | [–0.180, –0.088] | 0.87 |
| Mental health days | –0.007 | 0.003 | –2.152 | .031 | [–0.014, –0.001] | 0.99 |
| Physical health days | –0.004 | 0.003 | –1.258 | .209 | [–0.011, 0.002] | 1.00 |
| Difficulty walking | –0.980 | 0.104 | –9.433 | < .001 | [–1.183, –0.776] | 0.38 |
| Sex | –1.038 | 0.063 | –16.394 | < .001 | [–1.162, –0.914] | 0.35 |
| Age | 0.015 | 0.009 | 1.745 | .081 | [–0.002, 0.032] | 1.02 |
| Education | –0.159 | 0.019 | –8.538 | < .001 | [–0.196, –0.123] | 0.85 |
| Income | –0.112 | 0.014 | –8.273 | < .001 | [–0.139, –0.086] | 0.89 |

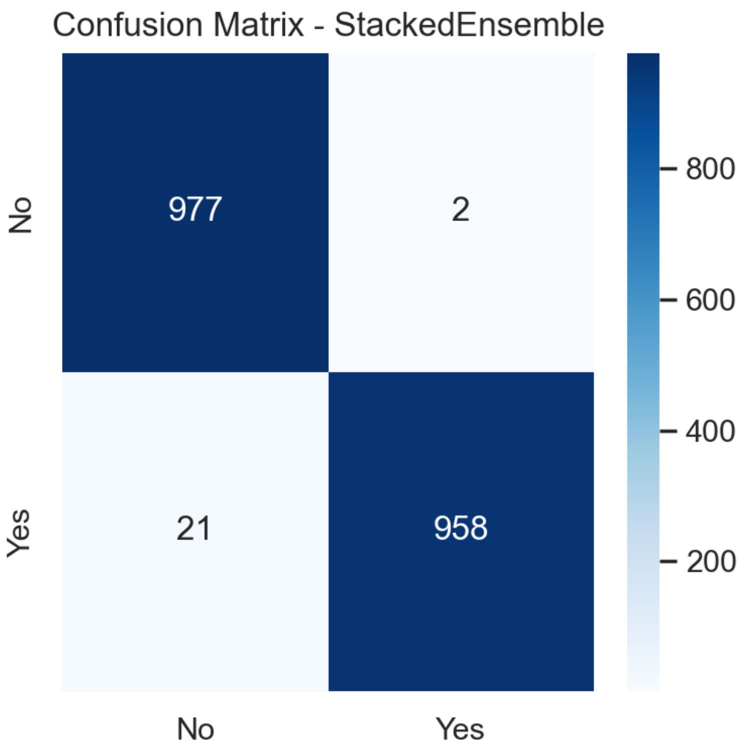


**Fig 2. Confusion Matrix for Stacked Ensemble Model**

higher rate of incorrect high-risk classifications. While the model maintains reasonable sensitivity, its lower precision suggests it may generate a significant number of false alerts, potentially detracting from its clinical utility by increasing unnecessary follow-up investigations.

The confusion matrix for the Decision Tree model, shown in Fig 8, reveals 882 true negatives and 863 true positives, with 97 false negatives and 116 false positives. This performance profile indicates a model with fair discriminative power but notable overfitting vulnerability, as evidenced by appreciable errors in both directions. The relatively higher error rates underscore the limitations of a single tree structure in managing the complexity and nonlinear interactions inherent in cardiovascular risk data.

As shown in Fig 9, the Keras MLP model's confusion matrix shows 806 true negatives, 785 true positives, 173 false negatives, and 194 false positives. The elevated counts of both types of errors indicate suboptimal learning from the tabular data, likely due to the model's architectural simplicity or insufficient feature representation capacity, even with extensive tuning or larger datasets. This results in decreased reliability for precise clinical stratification.

The Logistic Regression model's performance, depicted in Fig 10, yields 903 true negatives, 476 true positives, 503 false negatives, and 76 false positives. The extremely high number of false negatives highlights the model's critically low sensitivity, failing to identify a majority of actual stroke cases. While specificity remains high, this severe imbalance renders the model clinically inadequate for stroke risk screening, where missing true cases carries significant adverse consequences.

The confusion matrix for the Scikit-learn MLP classifier, presented in Fig 11, shows a model with moderate, balanced, yet suboptimal predictive performance for stroke risk in

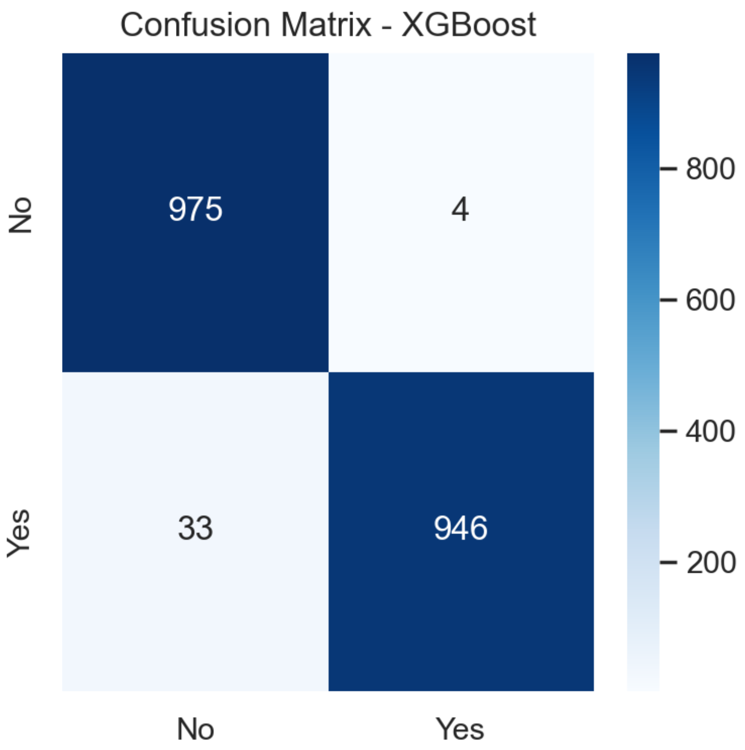


**Fig 3. Confusion Matrix for XGBoost Model**

patients with coronary heart disease. The model correctly identified 799 true negatives and 818 true positives, indicating reasonable overall discriminative capacity. However, with 180 false positives and 161 false negatives, the classifier exhibits substantial misclassification rates in both directions. The substantial number of false positives suggests reduced specificity, potentially leading to unnecessary clinical investigations and increased healthcare resource utilization. Conversely, the considerable count of false negatives indicates compromised sensitivity, meaning a significant portion of true stroke cases would be overlooked, posing a serious risk in preventive care contexts.

Finally, the confusion matrix for the Naive Bayes classifier, presented in Fig 12, shows 770 true negatives and 558 true positives, with 421 false negatives and 209 false positives. This model exhibits the weakest performance among the evaluated algorithms, with high rates of both missed strokes and false alarms. The poor performance underscores the inappropriateness of its conditional independence assumption for the complex, interrelated risk factors present in coronary heart disease and stroke prediction, severely limiting its practical application in this clinical domain.

#### Comparison of Machine Learning Algorithms for predicting Stroke among patients with heart disease

The machine learning results demonstrate apparent performance differences across the evaluated algorithms, with the stacked ensemble model providing the strongest and most stable predictive ability for Stroke among patients with coronary heart disease, as shown in Table 2. The stacked ensemble achieved the highest overall accuracy and one of the highest precision scores, reflecting its ability to combine the strengths of

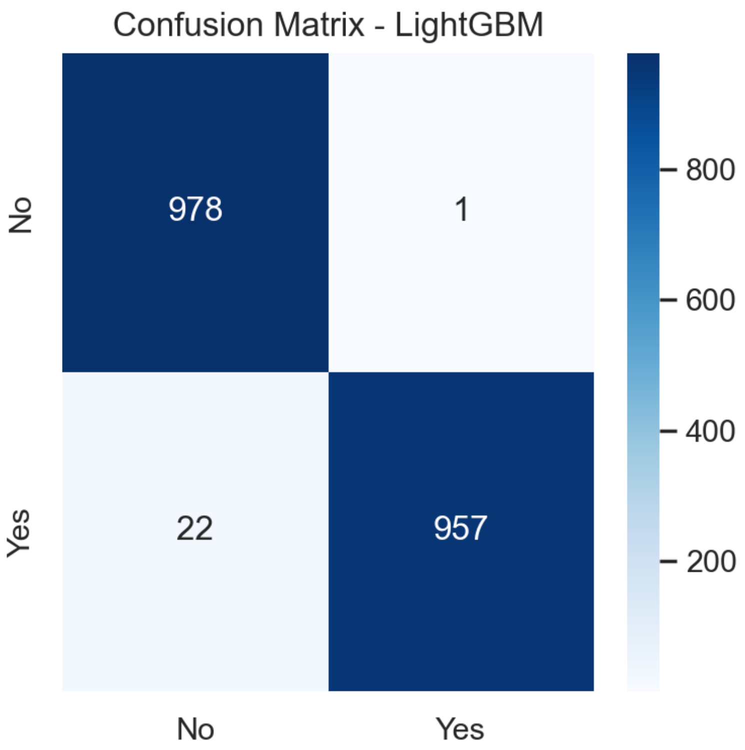


**Fig 4. Confusion Matrix for LightGBM Model**

individual models while reducing their weaknesses. Its high recall and specificity further confirm that it effectively identifies both Stroke and non-stroke cases with minimal error. The Matthews correlation coefficient, one of the most reliable measures for imbalanced clinical datasets, was also the highest for the stacked ensemble model, indicating consistent performance across all cells of the confusion matrix. The very low Brier score shows that the model produced well-calibrated probability estimates, making it suitable for real–world clinical decision-making.

LightGBM also performed exceptionally well and was nearly as good as the stacked ensemble. It achieved the highest precision of all models and maintained very high accuracy, recall, and specificity. The similarity in performance between LightGBM and the stacked ensemble underscores the advantage of gradient-boosting decision trees in structured clinical datasets. These models can capture complex interactions among predictors and remain robust to nonlinear relationships. XGBoost ranked slightly below LightGBM but still demonstrated strong metrics across accuracy, precision, recall, specificity, and F1 score. Its high Matthews correlation coefficient and relatively low Brier score indicate that it remained a reliable and stable model, although not as strong as the top two boosting methods.

Deep learning models showed moderate to strong performance. The Keras CNN model outperformed both the Keras MLP and the scikit learn MLP. It achieved high levels of accuracy, precision, and recall, suggesting that convolutional layers were effective in extracting meaningful representations from tabular data. The multilayer perceptron models had lower predictive scores, likely because they require larger datasets or additional feature engineering to fully capture complex clinical patterns. Their Matthews correlation coefficients and Brier scores were also weaker, indicating

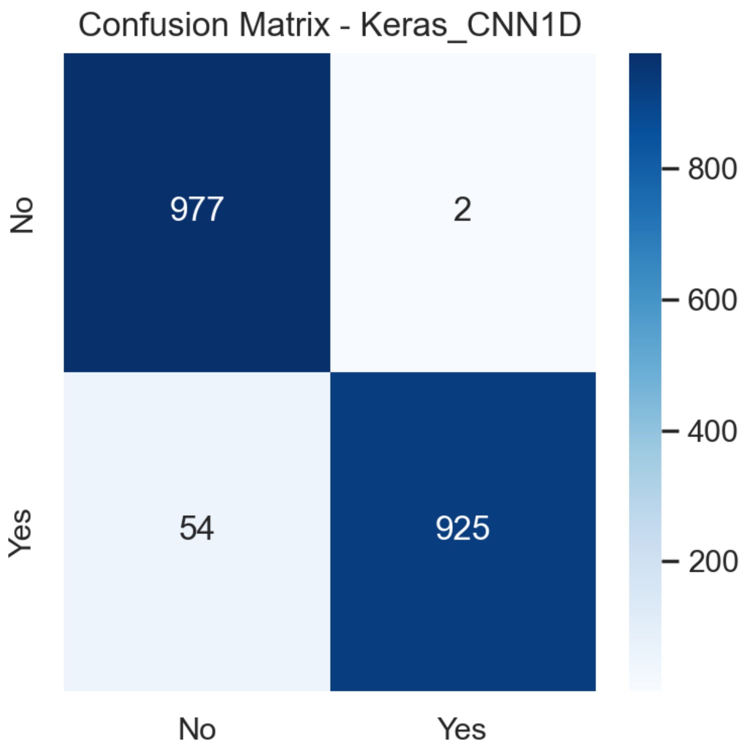


**Fig 5. Confusion Matrix for Keras CNN1D Model**

reduced stability and probability calibration.

Traditional machine learning models, such as Random Forest and Support Vector Machine, performed reasonably well but did not reach the level of the advanced gradient boosting or ensemble techniques. Random Forest achieved balanced accuracy and recall, with a relatively strong Matthew's correlation coefficient, suggesting it detected stroke cases more reliably than the Support Vector Machine. The Support Vector Machine showed good precision and specificity but had a comparatively lower recall, indicating a tendency to miss a proportion of true stroke cases.

Classical statistical models, including logistic Regression and naive Bayes, had the lowest performance across nearly all metrics. Logistic Regression achieved reasonably high specificity but extremely low recall, indicating that it failed to identify a substantial proportion of stroke cases, leading to a high number of false negatives. The F1 score and Matthew's correlation coefficient were also low, reinforcing its limited ability in this clinical prediction context. Naive Bayes performed similarly, with reduced accuracy, precision, and recall. Its high Brier score suggests poor calibration of predicted probabilities. These results align with established limitations of linear and distribution-dependent models when applied to complex nonlinear relationships common in cardiovascular health data.

Overall, the findings show that ensemble learning and gradient boosting techniques delivered the best predictive performance for stroke detection in patients with coronary heart disease. Their superior accuracy, balanced precision and recall, high Matthew's correlation coefficients, and low Brier scores demonstrate that they are more dependable and clinically meaningful than deep learning, traditional machine learning methods, or classical statistical models. Their strong performance supports their suitability for

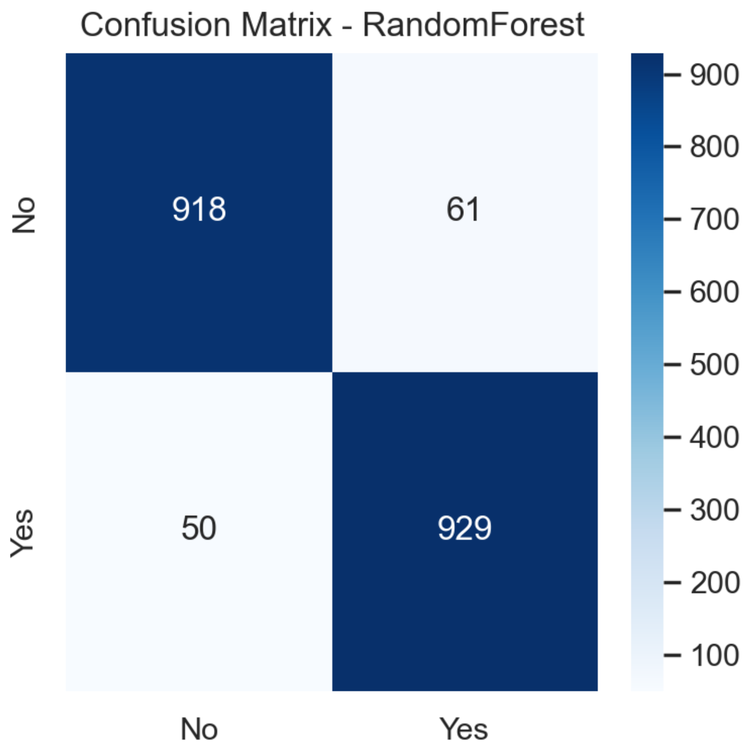


**Fig 6. Confusion Matrix for Random Forest Model**

clinical risk prediction and early intervention strategies aimed at preventing Stroke in high-risk cardiovascular populations.

**Table 2. Performance of Machine Learning Models for Stroke Prediction.**

| Model | Accuracy | Precision | Recall | Specificity | F1 | MCC | Brier Score |
|---|---|---|---|---|---|---|---|
| Stacked Ensemble | .9883 | .9979 | .9785 | .9980 | .9881 | .9767 | .0106 |
| LightGBM | .9883 | .9990 | .9775 | .9990 | .9881 | .9767 | .0108 |
| XGBoost | .9811 | .9958 | .9663 | .9959 | .9808 | .9626 | .0159 |
| Keras CNN | .9714 | .9978 | .9448 | .9980 | .9706 | .9441 | .0234 |
| Random Forest | .9433 | .9384 | .9489 | .9377 | .9436 | .8867 | .0547 |
| Support Vector Machine | .8968 | .9293 | .8590 | .9346 | .8928 | .7959 | .0791 |
| SKLearn MLP | .8258 | .8196 | .8355 | .8161 | .8275 | .6518 | .1382 |
| Decision Tree | .8912 | .8990 | .8815 | .9009 | .8901 | .7826 | .1088 |
| Keras MLP | .8126 | .8194 | .8018 | .8233 | .8105 | .6253 | .1384 |
| Logistic Regression | .7043 | .8623 | .4862 | .9224 | .6218 | .4540 | .1969 |
| Naive Bayes | .6782 | .7275 | .5700 | .7865 | .6392 | .3652 | .2737 |

Table notes: Performance metrics including accuracy, precision, recall, specificity, F1 score, Matthews correlation coefficient (MCC), and Brier score for each machine learning model on stroke prediction.

#### ROC Curve Analysis for Stroke Prediction Models

The receiver operating characteristic curves presented in Fig 13 show apparent differences in the discriminative ability of the machine learning algorithms used to

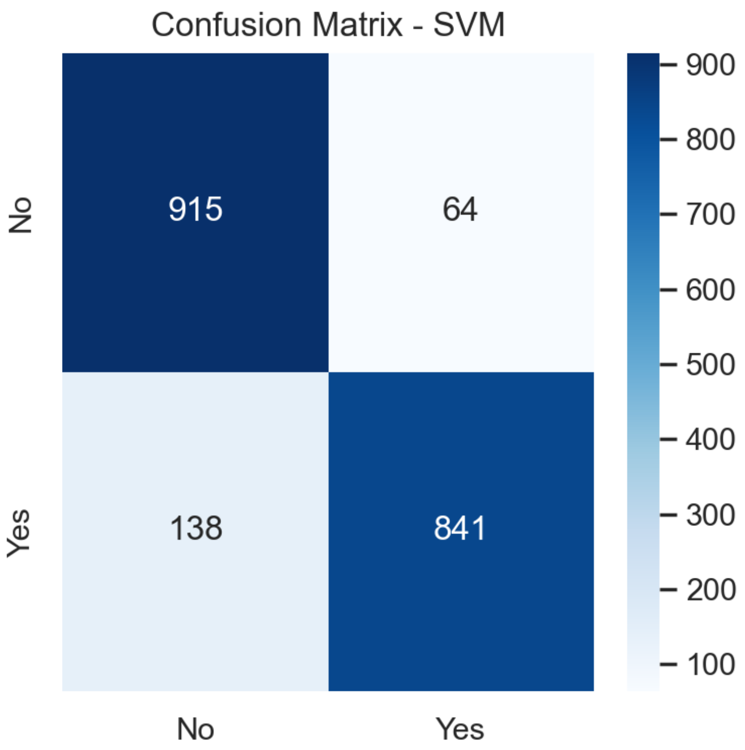


**Fig 7. Confusion Matrix for SVM Model**

predict Stroke among patients with coronary heart disease. The curves illustrate how well each model distinguishes between individuals with and without Stroke across all possible decision thresholds. The area under the curve values summarizes this performance, with larger values reflecting better separation between positive and negative cases.

The stacked ensemble and LightGBM models demonstrated the strongest discrimination, each producing an area under the curve value of approximately 0.994. Their curves remain close to the upper-left corner of the plot, indicating that they achieved very high true-positive rates even at low false-positive rates. XGBoost closely followed, with an area under the curve of 0.991, reflecting similarly strong classification performance. The Keras CNN model achieved an AUC of 0.988, and the Random Forest model reached 0.986. These results show that both deep learning and tree-based ensemble methods were highly effective at separating Stroke from non-stroke cases.

Support Vector Machine also performed well, with an area under the curve of 0.955, although its curve did not reach the same level as those of the gradient boosting and ensemble methods. The multilayer perceptron models showed moderate performance. The scikit-learn multilayer perceptron achieved an area under the curve of 0.904, while the Keras multilayer perceptron achieved 0.883. The decision tree model performed slightly lower with an area under the curve value of 0.891. These results show that simpler or less-optimized models had a reduced ability to distinguish between classes compared with advanced ensemble methods.

Logistic Regression and Naive Bayes recorded the lowest discrimination performance with area under the curve values of 0.740 and 0.739, respectively. Their curves remain close to the diagonal reference line shown in Figure 13, indicating limited capability to

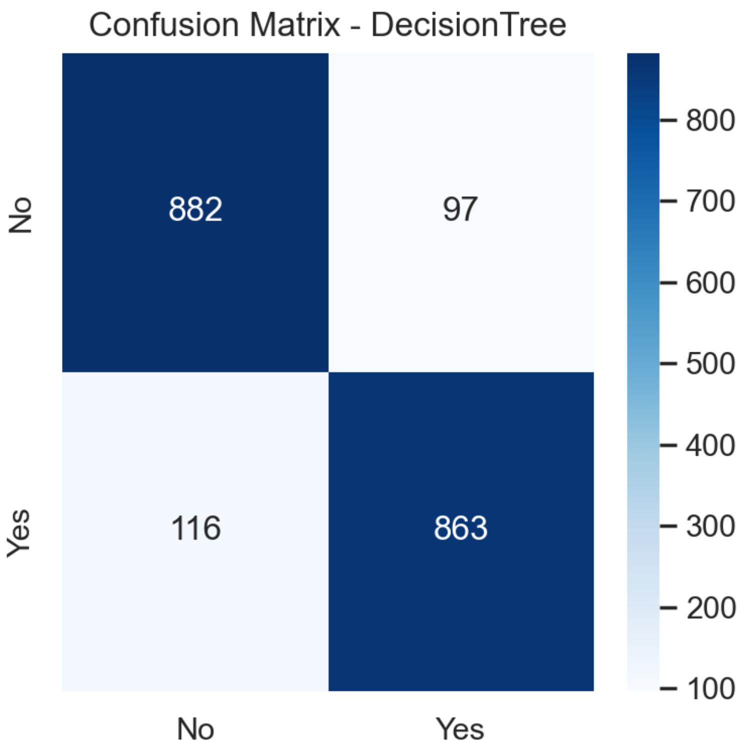


**Fig 8. Confusion Matrix for the Decision Tree Model**

separate Stroke and non-stroke cases. These patterns highlight the challenges that linear and distribution-based models face when applied to complex clinical data that involve nonlinear interactions.

Overall, the ROC AUC results provide strong evidence that ensemble and gradient boosting techniques deliver superior discrimination compared with deep learning, traditional machine learning, and classical statistical models. Fig 13 clearly demonstrates that the stacked ensemble, LightGBM, and XGBoost models are the most effective approaches for stroke prediction in patients with coronary heart disease, making them the most suitable candidates for integration into clinical decision support systems.

#### Precision-Recall Curve Analysis for Stroke Prediction Models

The precision-recall curves in Fig 14 provide further insight into the performance of the machine learning models in predicting Stroke among patients with coronary heart disease. Unlike the receiver operating characteristic curve, which evaluates performance across balanced conditions, the precision–recall curve emphasizes performance in situations where the positive class is less common. This characteristic makes it a handy tool for clinical datasets in which stroke cases represent a small proportion of the population. The area under the precision-recall curve summarizes this performance, with higher values indicating a stronger ability to identify true stroke cases while minimizing false positives.

The stacked ensemble model achieved the highest area under the precision-recall curve, with a value of 0.996. Its curve remains close to the upper boundary throughout the range of recall values, showing that it consistently maintains high precision even when identifying a large proportion of true stroke cases. LightGBM performed almost

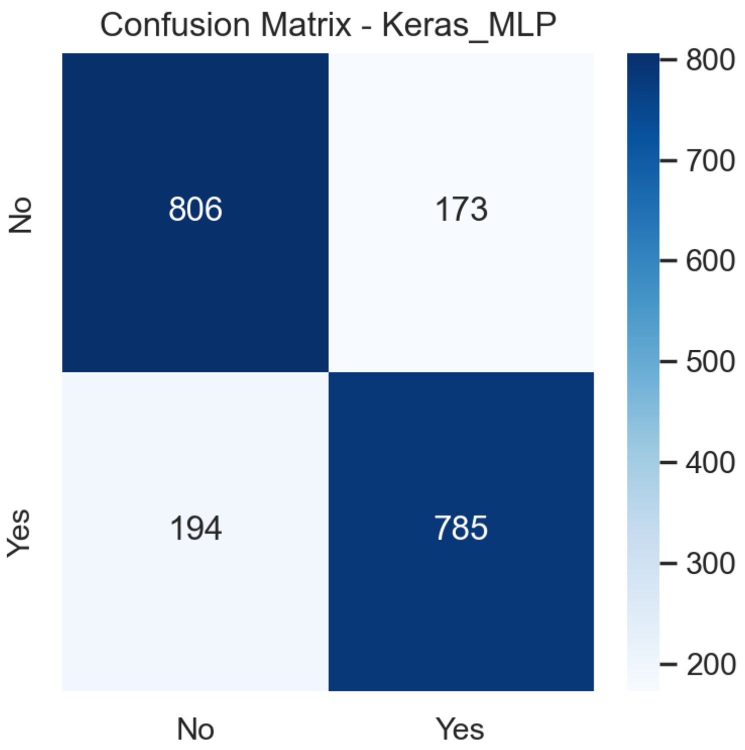


**Fig 9. Confusion Matrix for Keras MLP Model**

identically, with an area under the precision-recall curve of 0.996, further confirming its robust discrimination and reliability under imbalanced conditions. XGBoost achieved an area under the precision-recall curve of 0.995, demonstrating similarly strong performance. These three models show minimal performance loss as recall increases, highlighting the stability of gradient boosting and ensemble strategies.

The Keras CNN model performed very well, with an area under the precision-recall curve of 0.992, confirming the ability of convolutional layers to extract meaningful patterns from tabular clinical data. Random Forest also achieved strong performance with a value of 0.987. Although these values are slightly lower than those of the boosting models, their curves remain well above the baseline in Fig 14, indicating consistent separation of positive and negative cases even when the dataset is imbalanced.

The Support Vector Machine showed moderate performance, with an area under the precision-recall curve of 0.958. Its curve indicates that the model achieves reasonable precision but shows a steeper decline as recall increases. The multilayer perceptron models showed weaker performance, with the scikit-learn model achieving an area under the precision-recall curve of 0.905 and the Keras model achieving 0.886. The decision tree model performed slightly worse, with an area under the precision-recall curve of 0.852. These results reflect the limitations of models that rely solely on shallow tree structures or fully connected layers when applied to complex clinical data.

Logistic Regression and Naive Bayes achieved the lowest performance with values of 0.741 and 0.761, respectively. Their precision-recall curves in Fig 14 approach the baseline, indicating limited ability to correctly identify stroke cases, especially at higher recall levels. These models are less able to maintain precision as they attempt to identify more true stroke cases, thereby reducing their suitability for clinical use in early

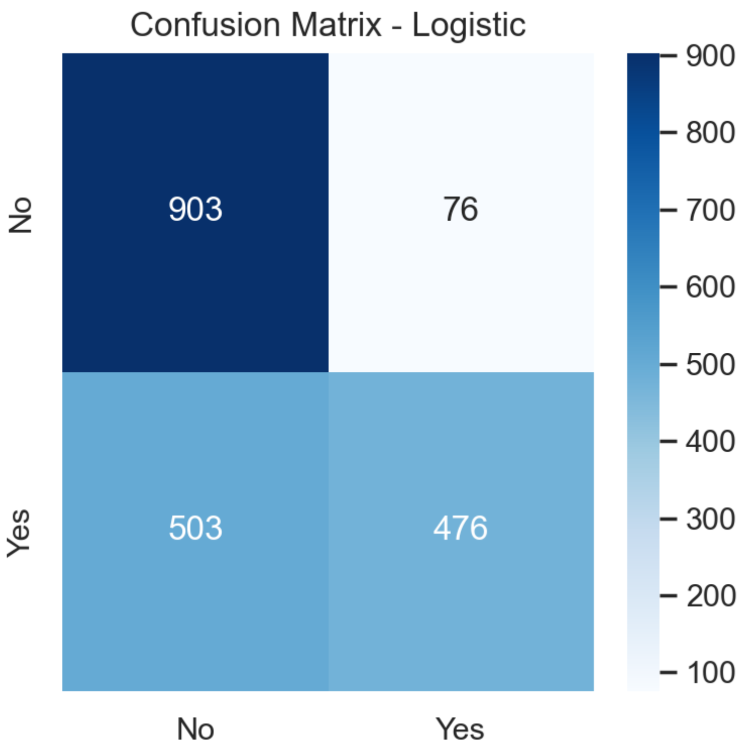


**Fig 10. Confusion Matrix for Logistic Regression Model**

stroke risk detection.

Overall, the area under the precision-recall curves in Fig 14 confirms that the stacked ensemble, LightGBM, and XGBoost models deliver the strongest performance in identifying stroke cases despite class imbalance. Their curves remain consistently high across the full range of recall values, reinforcing their superiority over deep learning models, traditional machine learning approaches, and classical statistical methods.

#### Calibration Curve Analysis for Stroke Prediction Models

The calibration curves for all evaluated machine learning models provide a critical assessment of the reliability of predicted stroke risk probabilities among patients with coronary heart disease. Calibration curves plot the observed event frequency against the mean predicted probability for binned predictions, with ideal calibration represented by a diagonal line where the predicted probabilities exactly match the observed outcomes. As shown in Fig 15, the Stacked Ensemble and LightGBM models demonstrated near-perfect calibration, with their curves closely aligning with the ideal diagonal. This is supported quantitatively by their exceptionally low Brier scores of 0.011, indicating minimal discrepancy between predicted risks and observed event rates. Such high calibration is clinically essential, as it ensures that a predicted stroke risk of, for example, 20% corresponds to an actual event frequency of approximately 20% in similar patient groups, thereby enabling trustworthy individualized risk stratification.

Fig 15 also reveals distinct tiers of calibration performance among the other models. XGBoost and the Keras CNN1D model followed with strong calibration, evidenced by their curves remaining close to the diagonal and their low Brier scores of 0.016 and 0.023, respectively. Models such as Random Forest and Support Vector Machine

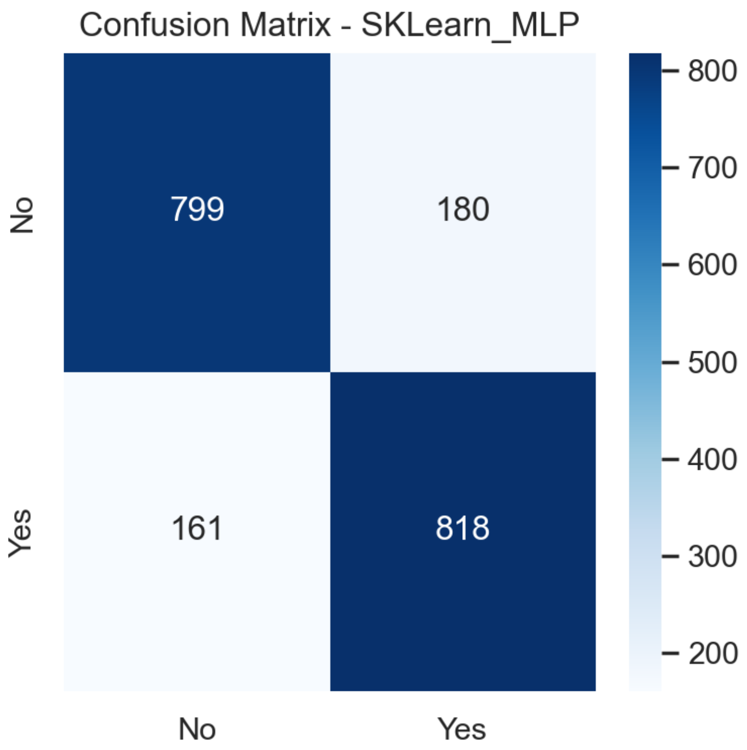


**Fig 11. Confusion Matrix for Scikit-learn MLP classifier**

exhibited moderate calibration, with visible deviations from the ideal line and higher Brier scores of 0.055 and 0.079, suggesting a tendency towards underconfidence or overconfidence in specific probability ranges. In contrast, classical models such as Logistic Regression and Naive Bayes showed poor calibration, with curves deviating substantially from the diagonal and Brier scores of 0.197 and 0.274, respectively. This indicates that their predicted probabilities are not well aligned with actual outcomes, limiting their utility for precise clinical risk estimation. The overall pattern in Fig 15 reinforces that advanced ensemble and gradient boosting methods not only achieve superior discriminative performance but also produce well-calibrated probability estimates, which are crucial for informed clinical decision-making and the implementation of personalized preventive strategies.

#### Learning Curve Analysis for Stroke Prediction Models

The learning curve for the LightGBM model shown in Fig 16 demonstrates excellent generalization capability. Both the training and cross-validation F1 scores rise quickly and converge to a high, stable plateau as the number of training examples increases. The small gap between the training and validation curves indicates that the model is learning the underlying patterns effectively without overfitting. This robust learning behavior supports LightGBM's suitability for deployment on clinical datasets of varying sizes, ensuring reliable performance in real-world settings.

As shown in Fig 17, the XGBoost model exhibits a similarly strong learning trajectory. The training and cross-validation F1 scores improve consistently with more data, converging at a high level of performance. A slight, persistent gap between the curves suggests a minor degree of overfitting, which is well managed by the model's

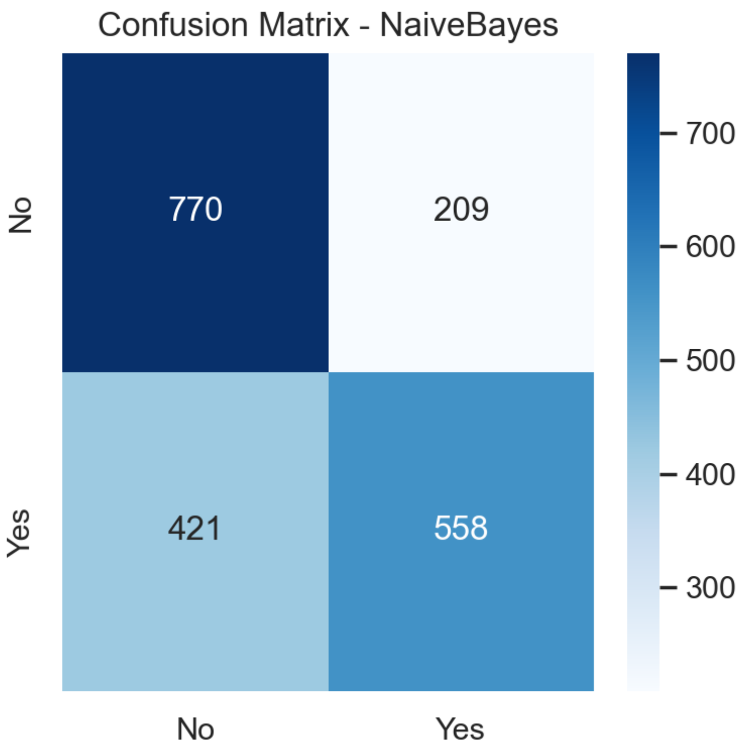


**Fig 12. Confusion Matrix for Naive Bayes classifier**

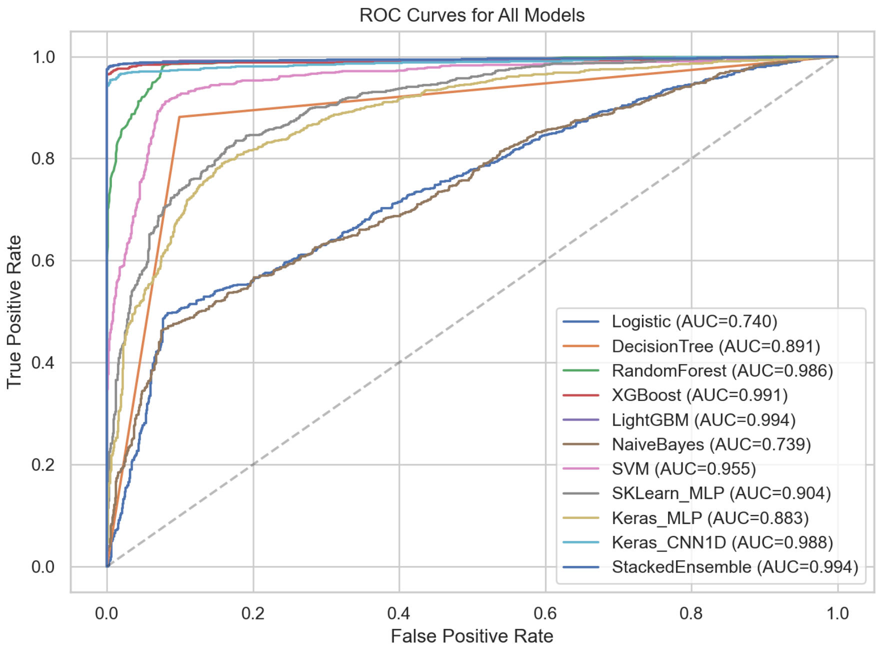


**Fig 13. AUC–ROC Curves for ML Algorithms**

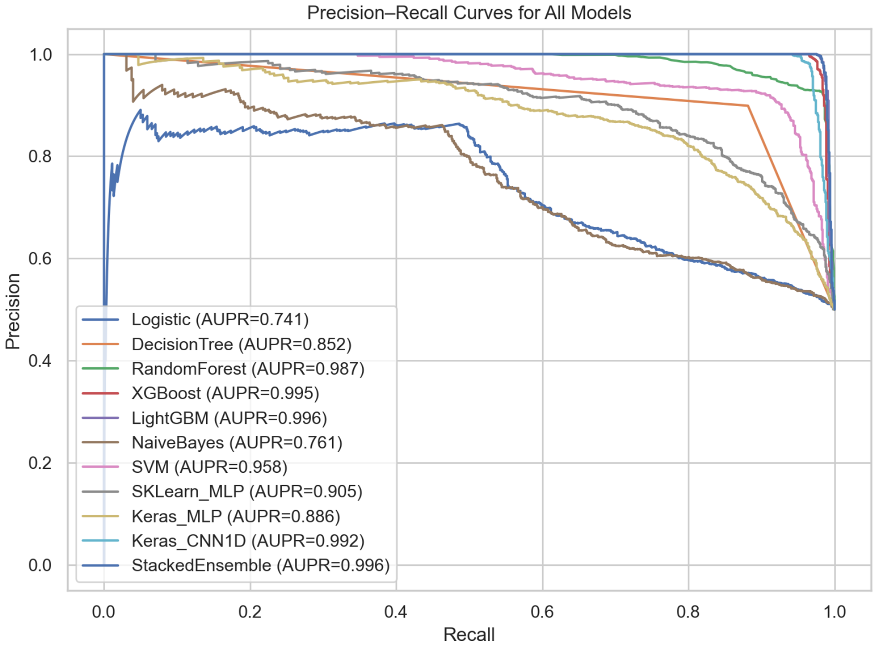


**Fig 14. Precision-Recall Curve for ML Algorithm**

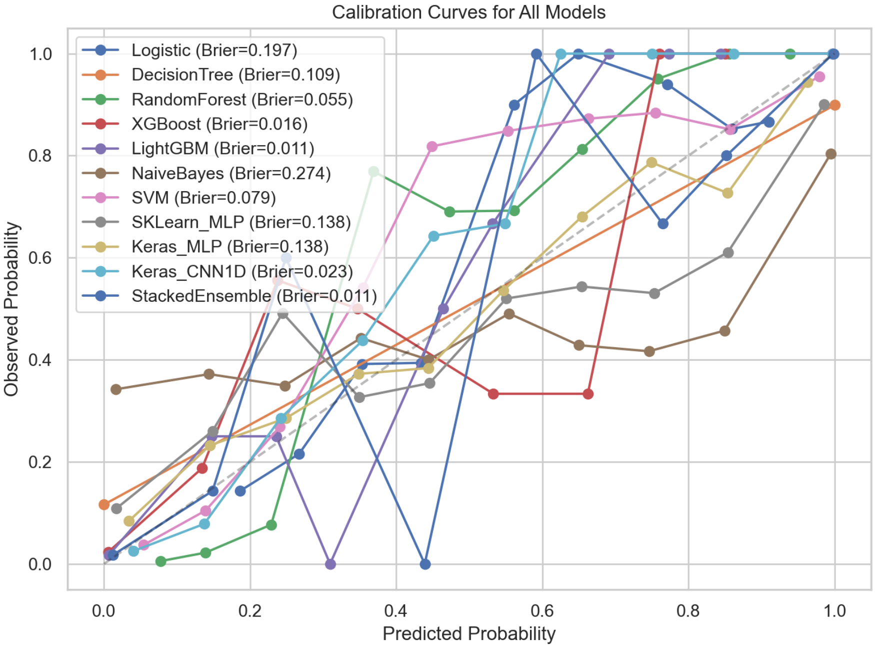


**Fig 15. Calibration Curves for ML Algorithms**

built-in regularization. This pattern confirms XGBoost's ability to leverage additional data to improve its predictive stability and accuracy in stroke risk prediction.

Fig 18 displays the learning curve for the Random Forest model. While both curves achieve high final F1 scores, a consistent and noticeable gap exists between their training and validation performance. This indicates that the model is overfitting to the

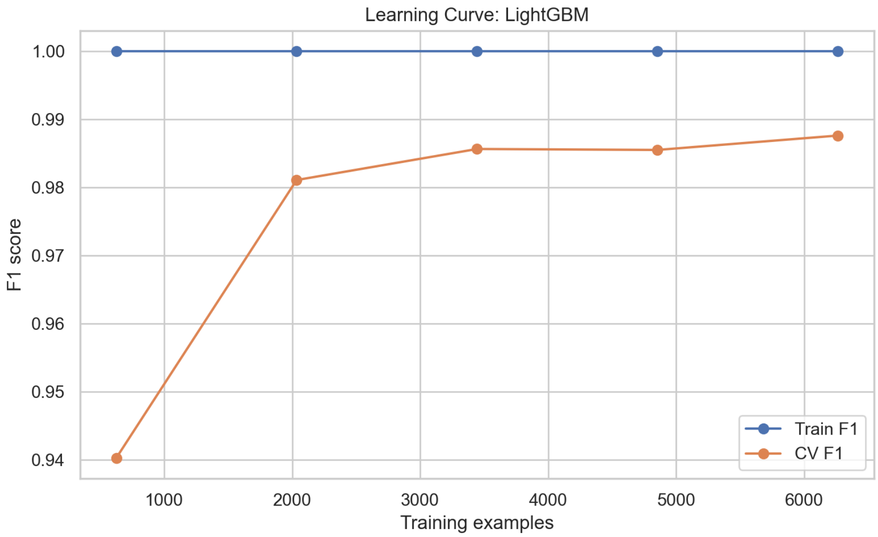


**Fig 16. Learning Curve for Light GBM Model**

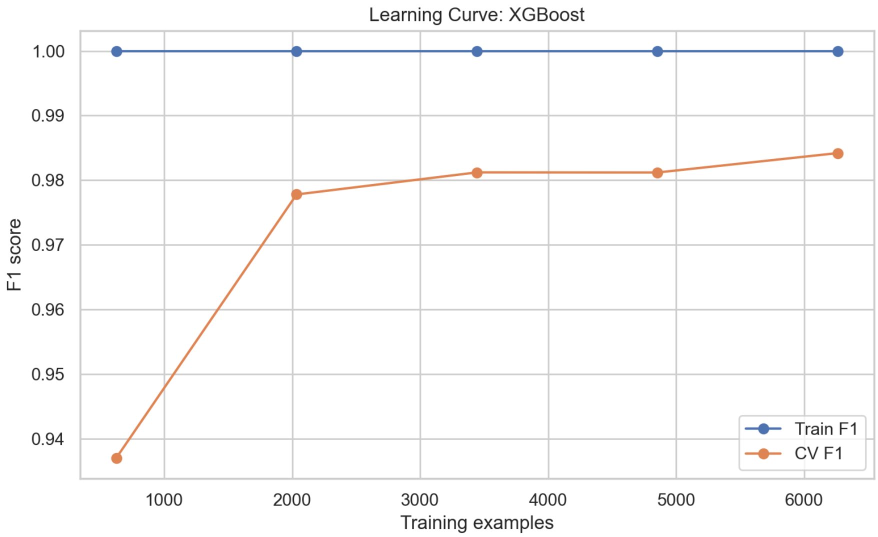


**Fig 17. Learning Curve for XGBoost Model**

training data to some extent, a common trait of bagging ensembles that memorize noise. Nevertheless, the cross-validation score remains strong, confirming the model's utility, though it benefits less from additional data compared to boosting methods.

The training history for the Keras CNN1D model shown in Fig 19 shows a smooth, rapid decline in both training and validation loss over epochs. The two curves decrease in close concordance and stabilize at a low level of loss. This ideal convergence indicates that the convolutional architecture learned meaningful feature representations from the tabular data effectively, without signs of overfitting, and that the training process was well-regularized.

In contrast, the learning curve for the Decision Tree model, shown in Fig 20, reveals a characteristic pattern of high variance. The training F1 score reaches near-perfect levels very quickly, while the cross-validation F1 score plateaus at a significantly lower

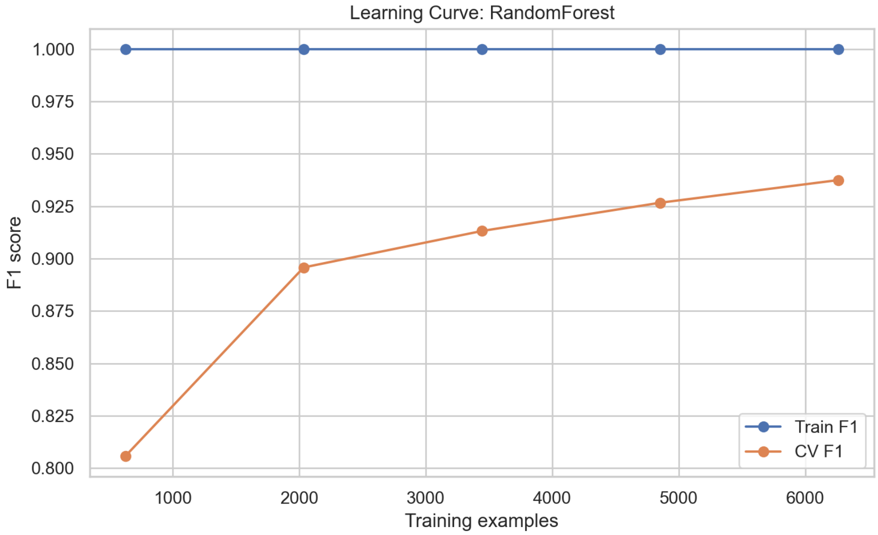


**Fig 18. Learning Curve for Random Forest Model**

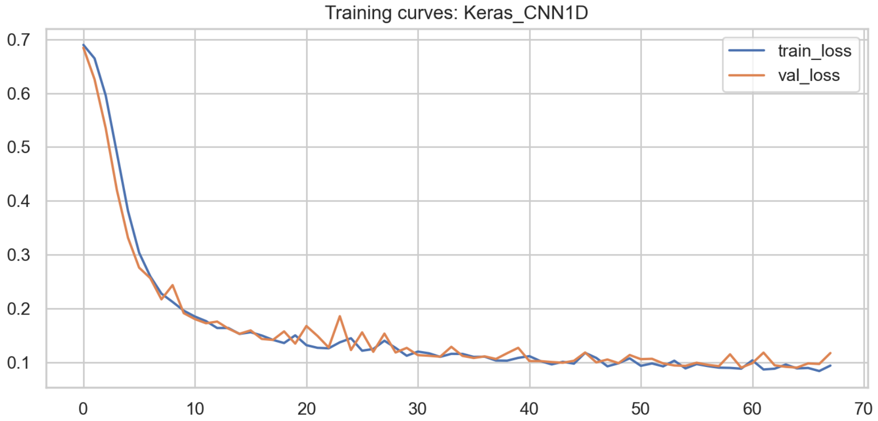


**Fig 19. Training Curves for Keras CNN1D Model**

value. This large and widening gap is a clear sign of severe overfitting: the single tree structure memorizes the training data but fails to generalize well to unseen cases, limiting its clinical reliability.

The learning curve for the Support Vector Machine (SVM), shown in Fig 21, indicates moderate final performance with a concerning trend. As the training size increases, the gap between the training and cross-validation F1 scores widens. This suggests that the SVM model is becoming increasingly overfit as more data are added, likely due to the complexity of the kernelized decision boundary in the high-dimensional space of clinical features.

As shown in Fig 22, the Scikit-learn MLP classifier exhibits high variance in its learning curve. The training F1 score is consistently and significantly higher than the cross-validation score, which plateaus at a moderate level. This substantial gap indicates that the multilayer perceptron is overfitting to the training set and may require more aggressive regularization, architectural adjustments, or substantially more data to improve its generalization.

The training history for the Keras MLP model shown in Fig 23 indicates that the

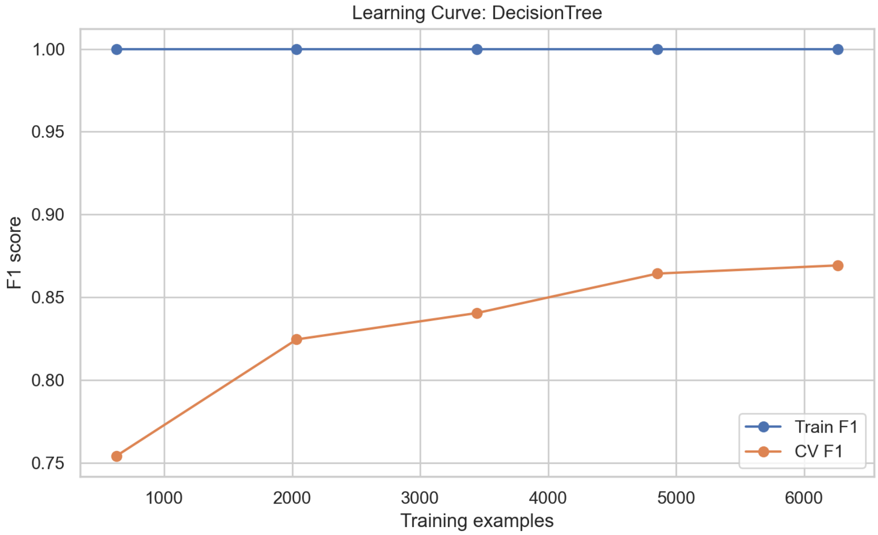


**Fig 20. Learning Curve for Decision Tree Model**

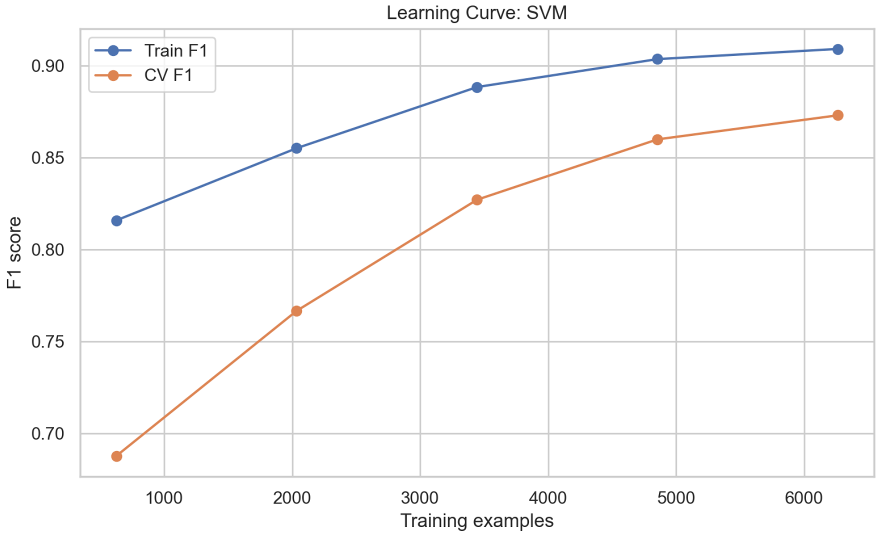


**Fig 21. Learning Curve for SVM Model**

training loss decreases rapidly. However, the validation loss plateaus after a few epochs and even shows a slight upward trend later in training. This divergence is a classic indicator of overfitting, suggesting that the model would benefit from techniques such as early stopping, dropout, or reduced model complexity to improve its validation performance.

Fig 24 illustrates the learning curve for Logistic Regression. Both the training and cross-validation F1 scores remain low and stable across all dataset sizes, with almost no gap between them. This flat, low-performance profile is a definitive sign of underfitting, indicating that the linear model is fundamentally too simple to capture the complex, nonlinear relationships governing stroke risk in this patient population.

Finally, the learning curve for the Naive Bayes classifier, shown in Fig 25, confirms its severe limitations. The training and cross-validation F1 scores are low, stable, and

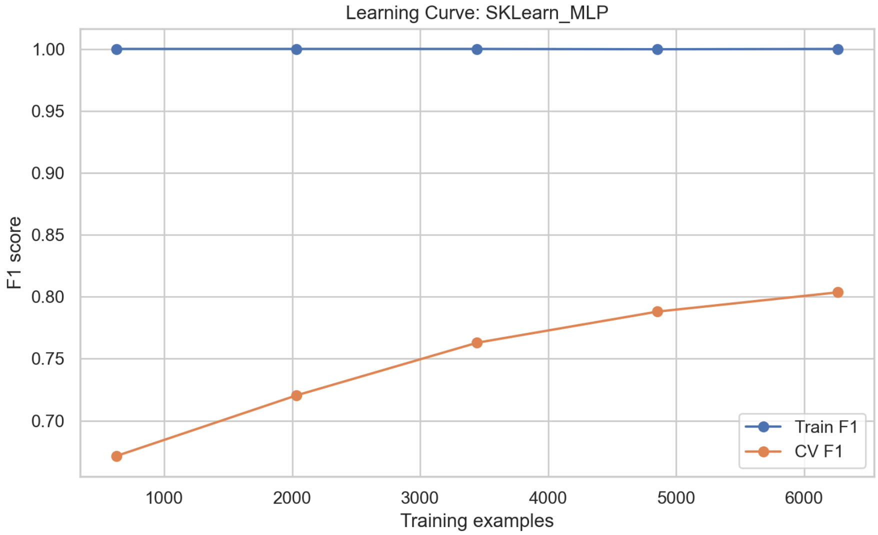


**Fig 22. Learning Curve for Scikit-learn MLP Classifier**

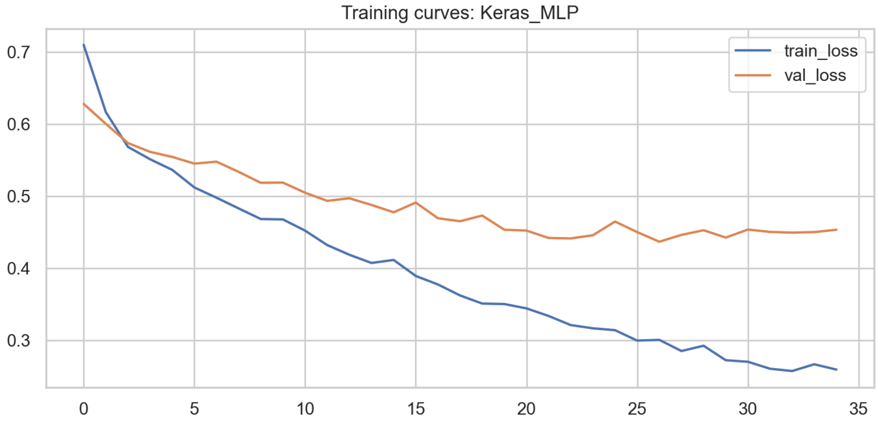


**Fig 23. Training Curves for Keras MLP model**

virtually identical across the smallest and largest training sets. This complete lack of improvement with additional data, coupled with the model's poor asymptotic performance, underscores that its conditional independence assumption is too restrictive for this complex clinical prediction task, leading to persistent underfitting.

#### Scalability Analysis for Machine Learning Model Deployment

Scalability was evaluated to determine the feasibility of deploying the trained models in real-world clinical environments, particularly those with constrained computational resources such as primary health facilities, mobile health systems, and edge devices. The assessment focused on four dimensions of computational efficiency: inference latency, training time, memory requirements during model training and inference, and final model size, as shown in Table 3. These metrics collectively indicate whether each model can operate effectively in settings with limited hardware capacity, response time, and storage.

Inference latency refers to the average time required for a model to generate a

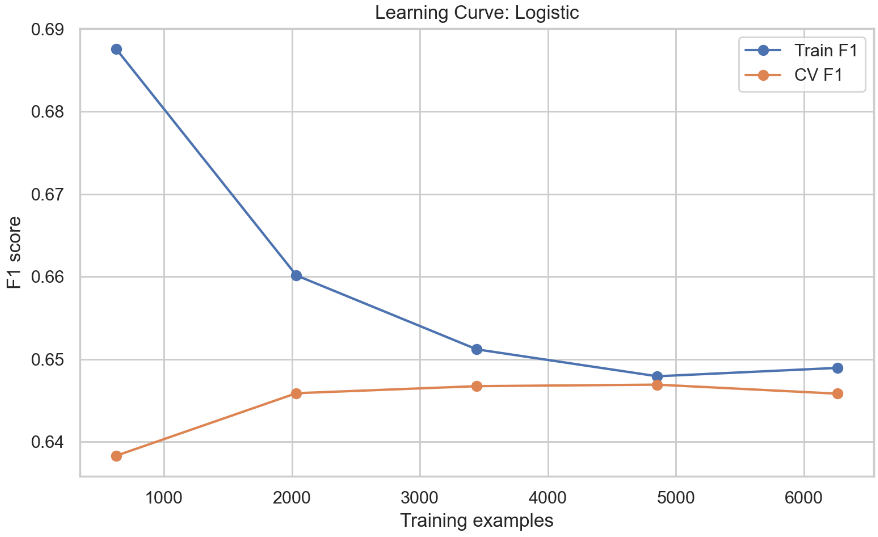


**Fig 24. Learning Curves for Logistic Regression Model**

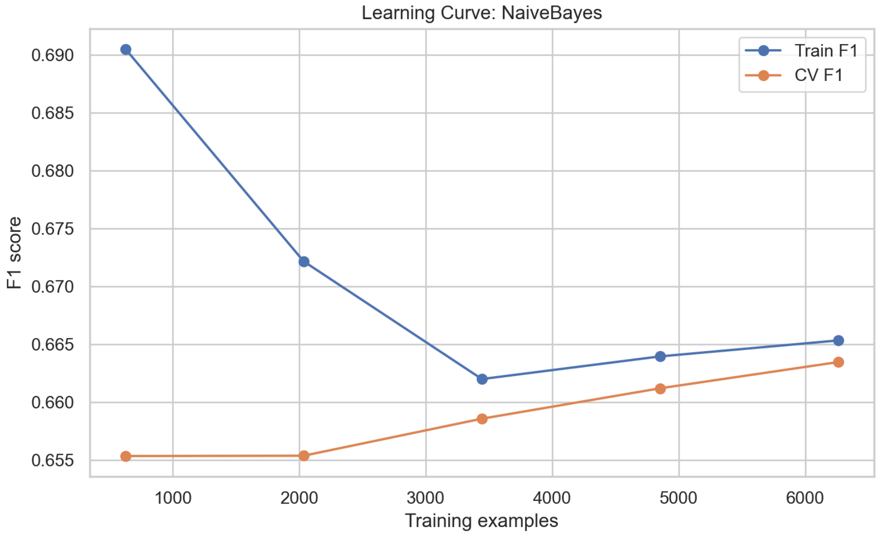


**Fig 25. Learning Curve for Naive Bayes Classifier**

prediction for a single patient. Low latency is essential for use cases such as point-of-care screening, triage support, and mobile health applications that require real-time responses. The results show that most models have microsecond-level inference times, with the Decision Tree and SKLearn_MLP emerging as the fastest learners. In contrast, the Keras_CNN1D and Support Vector Machine models demonstrated higher inference times, reflecting the computational intensity of convolutional and kernel-based operations.

**Table 3. Scalability Metrics for Machine Learning Models.**

| **Model** | **Latency (s)** | **Train Time (s)** | **Train Mem (MB)** | **Infer Mem (MB)** | **Size (KB)** |
|---|---|---|---|---|---|
| LightGBM | 2.15e-5 | 1.03 | 10.01 | 0.15 | 685.80 |
| XGBoost | 5.57e-6 | 1.60 | 9.06 | 0.07 | 400.21 |
| Keras CNN1D | 1.77e-4 | 49.15 | 44.47 | 0.63 | 135.77 |
| Random Forest | 4.81e-5 | 1.48 | 32.06 | 0.02 | 32392.67 |
| SVM | 9.59e-4 | 27.20 | 20.40 | 0.02 | 2357.71 |
| SKLearn MLP | 1.22e-6 | 19.07 | 1.10 | 0.23 | 219.46 |
| Decision Tree | 5.81e-7 | 0.15 | 0.69 | 0.01 | 98.82 |
| Keras MLP | 1.73e-4 | 16.64 | 226.91 | 0.52 | 245.88 |
| Logistic Reg. | 1.26e-6 | 0.04 | 3.16 | 0.04 | 1.36 |
| Naive Bayes | 1.77e-6 | 0.01 | 0.11 | 1.94 | 2.76 |

#### Synthesis and Clinical Utility

Collectively, these visualizations move the model from a "black box" to a transparent and auditable tool. The global plots (Figs 26, 27, 28, 29) validate the model at a population level, ensuring that the most influential features are clinically plausible. The local plots (Figs 31–36) empower clinicians at the point of care. They can answer the critical question: ”Why did the model predict a high stroke risk for *this* particular patient?”

To fully leverage this in your manuscript, ensure you reference these figures when discussing model interpretability in the Results and Discussion sections. For example:

> ”The superior performance of the LightGBM model was complemented by its interpretability. As shown in Fig 30, BMI and history of heart disease were the most impactful features globally. Furthermore, local explanations for individual high-risk predictions (Fig 34) consistently highlighted advanced age and low vegetable intake as contributing factors, providing actionable insights for targeted patient counseling.”

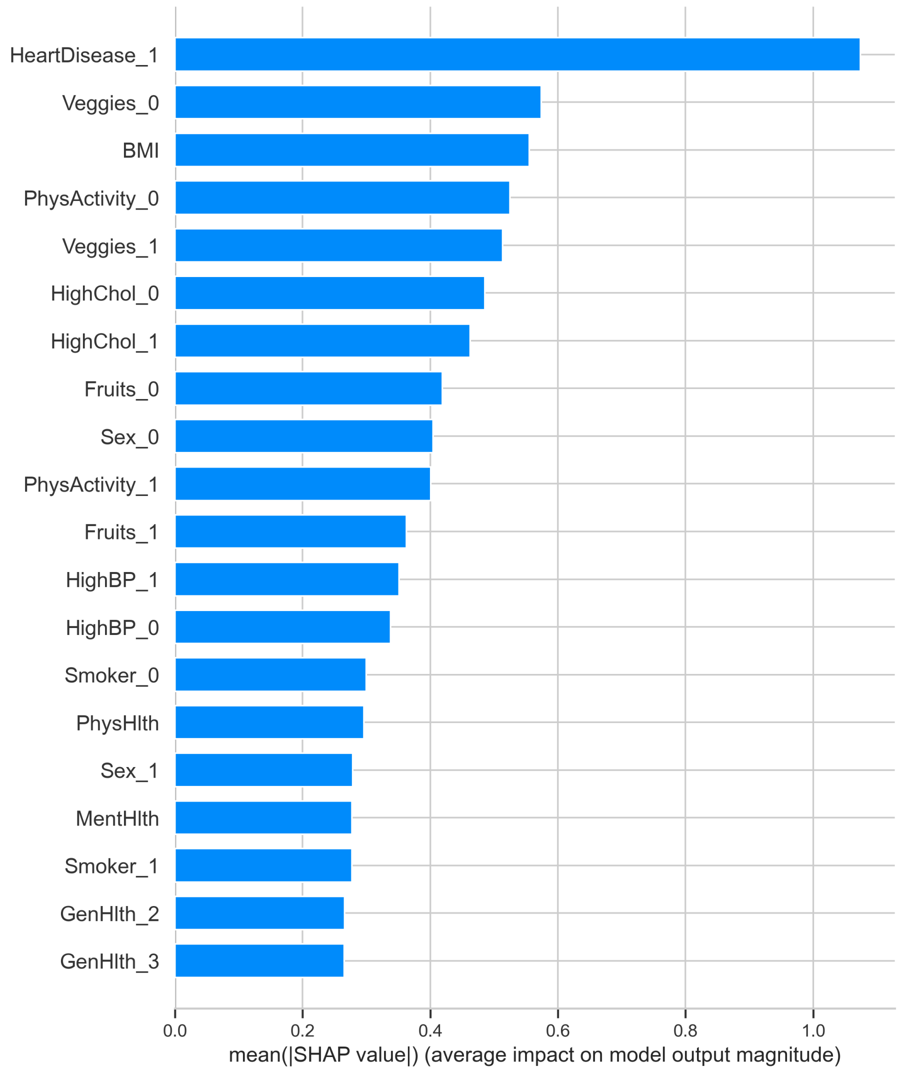


**Fig 26.** Global feature importance (mean absolute SHAP value) for the XGBoost model.

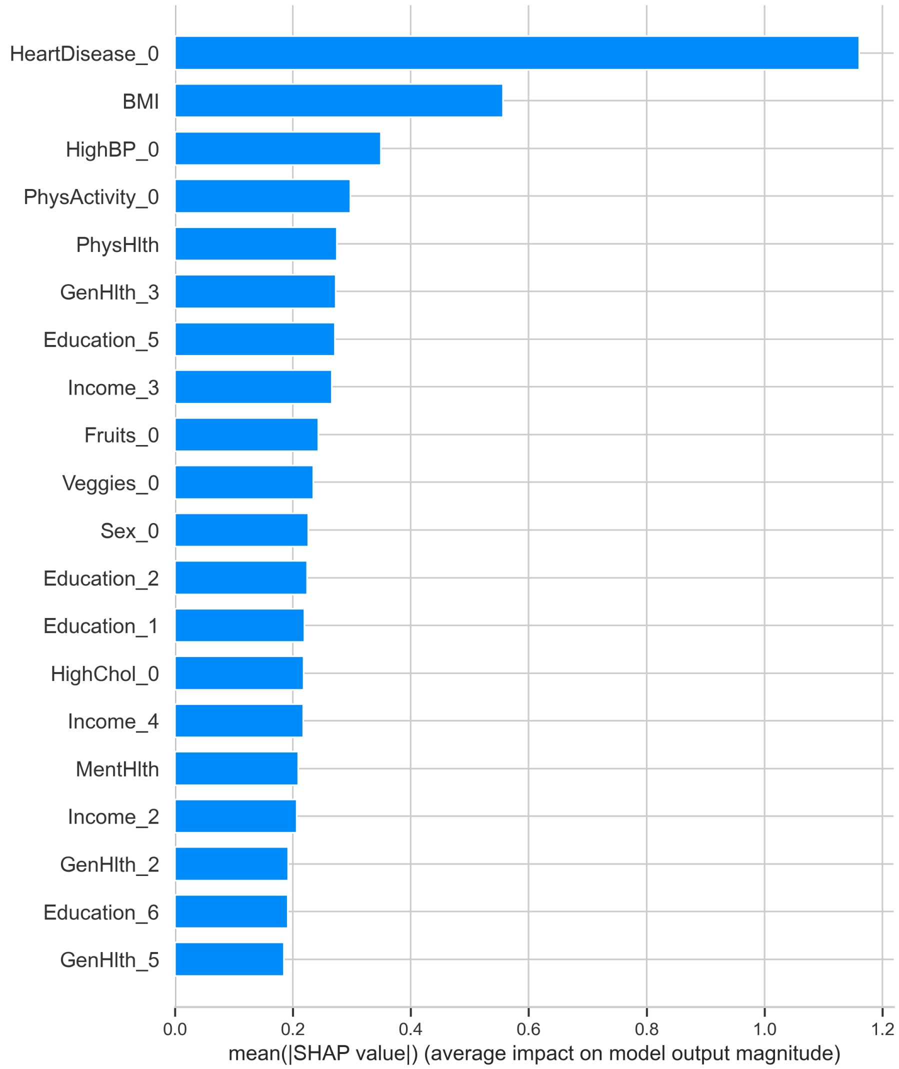


**Fig 27.** Global feature importance (mean absolute SHAP value) for the LightGBM model.

#### Discussion

The current research examined how various machine learning models can be used to predict stroke risk in patients with coronary heart disease and yielded meaningful results that align with the general literature on the topic while introducing new insights. Overall, the findings indicate that the best predictive performance was obtained with high-level ensemble learning techniques and gradient boosting algorithms, compared to deep learning, classic machine learning, and classical statistical techniques. This result aligns with the growing body of cardiovascular and cerebrovascular literature indicating that ensemble techniques are more successful at identifying complex trends in structured clinical data than individual models [2–5].

The robust performance of stacked ensemble, LightGBM, and XGBoost models in this study supports earlier findings that such approaches can outperform individual ML and deep learning models in stroke and cardiac risk prediction [6, 8, 9]. The effectiveness of gradient boosting, in particular, underscores the ability of tree-based

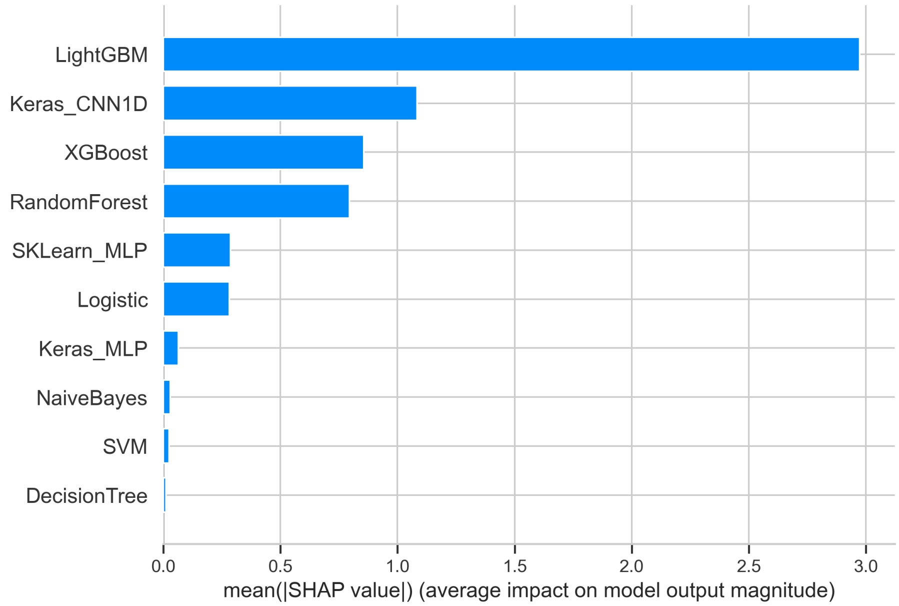


**Fig 28.** Comparison of global feature importance across all evaluated models.

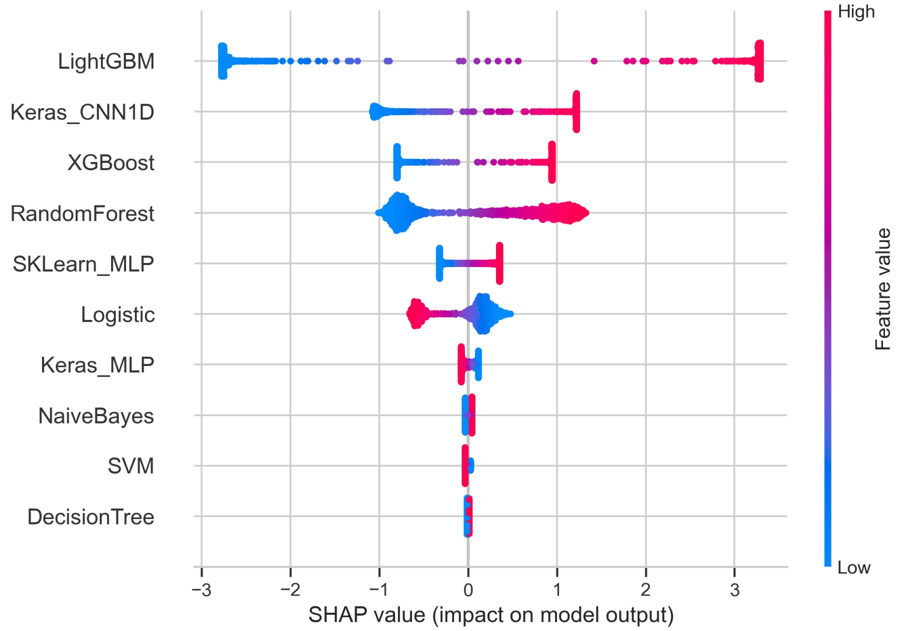


**Fig 29.** SHAP beeswarm plot showing feature effects across all models. Each point represents a patient, colored by feature value.

ensembles to model nonlinear interactions, which are essential for accurately capturing cardiovascular risk profiles. These findings complement prior studies demonstrating that ensemble-based structures yield superior discrimination and calibration relative to single models [2, 3, 31].

Deep learning models showed moderate to high predictive value, with convolutional neural networks outperforming fully connected multilayer perceptrons, consistent with prior work showing that convolutional models can learn meaningful representations from

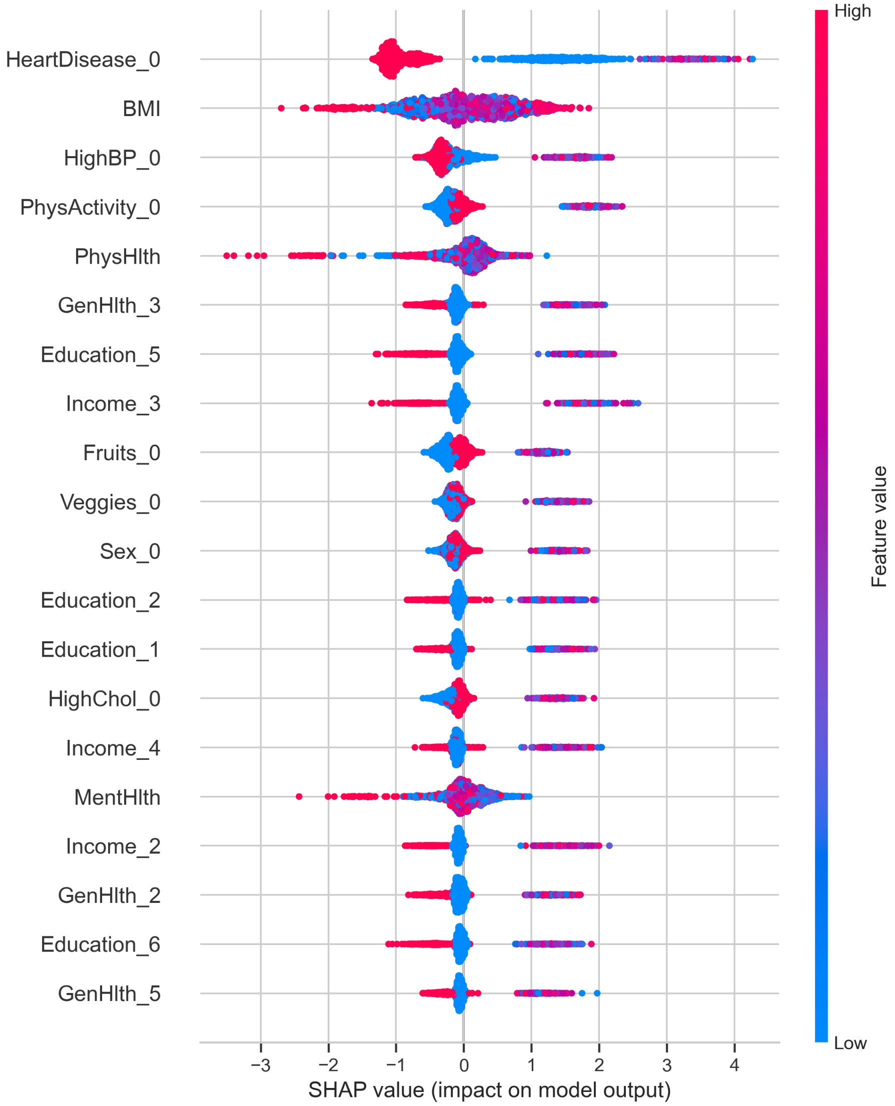


**Fig 30.** SHAP beeswarm plot for the LightGBM model. The plot illustrates how each feature contributes to model predictions across the dataset.

tabular clinical data but may require very large datasets to surpass ensemble methods [2, 26]. Similarly, conventional ML algorithms such as support vector machines and random forests delivered reasonable performance but were generally less discriminative than gradient-boosted ensembles, echoing earlier observations [10, 13, 19]. Decision trees exhibited clear signs of overfitting, reflecting the known limitations of single-tree models in complex medical prediction tasks.

Classical statistical models, including logistic regression and naive Bayes, performed the worst, consistent with research indicating that linear or distribution-dependent models often underfit high-interaction and mixed-type features in clinical data [15–17]. The low recall rates of logistic regression in predicting stroke cases among coronary heart disease patients highlight the need for more flexible modelling approaches, as reinforced by studies incorporating ML-based prognostication [31].

Scalability analysis further demonstrates that ensemble models like LightGBM and

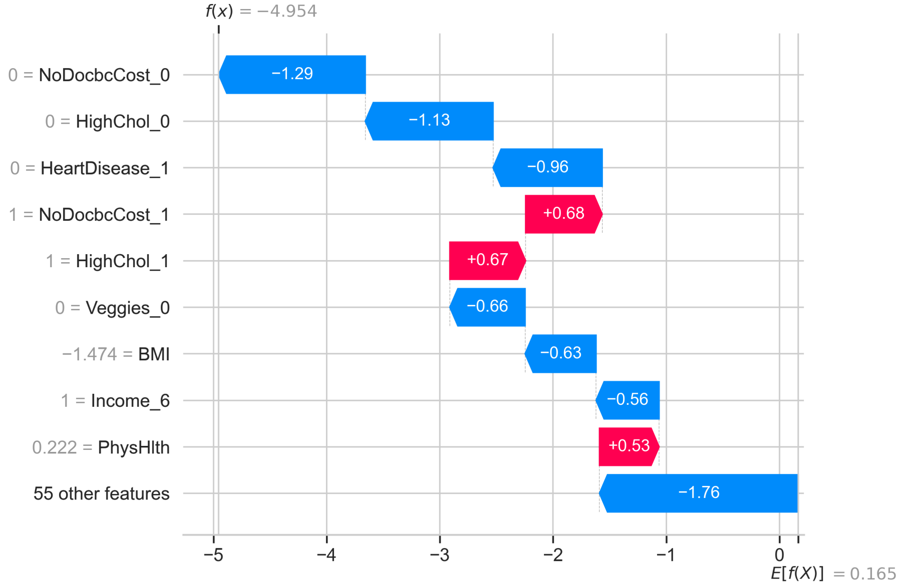


**Fig 31.** Local SHAP explanation for a low-risk prediction from the XGBoost model. The plot shows how individual features push the prediction away from the base value.

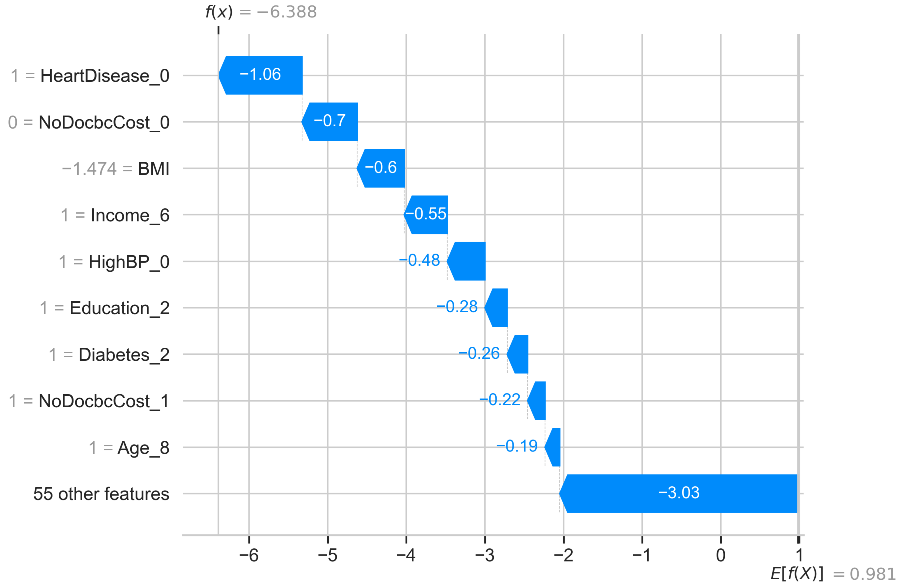


**Fig 32.** Local SHAP explanation for a low-risk prediction from the LightGBM model.

XGBoost are not only highly accurate but also computationally efficient, making them suitable for integration into clinical decision support systems, including in resource-limited settings. Deep learning models, while more memory-intensive and slower to train, offer reasonable inference efficiency, suggesting potential utility in cloud-hosted clinical applications [24, 27, 32].

Explainability analyses revealed that SHAP-based global explanations consistently highlighted age, prior cardiovascular events, BMI, and general health indicators as dominant contributors to stroke risk, corroborating prior studies [2, 6, 10, 14]. The observed consistency of feature rankings across LightGBM and XGBoost aligns with

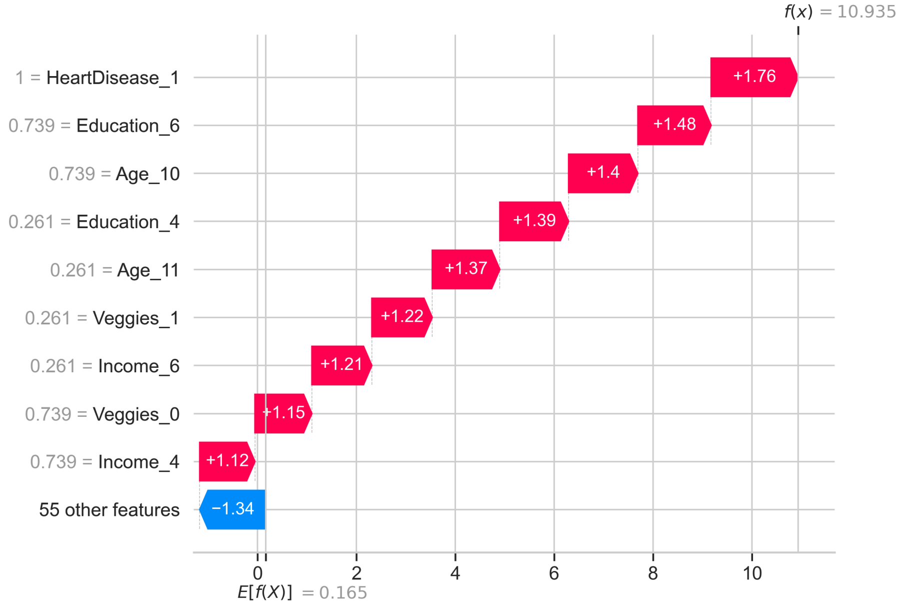


**Fig 33.** Local SHAP explanation for a high-risk prediction from the XGBoost model.

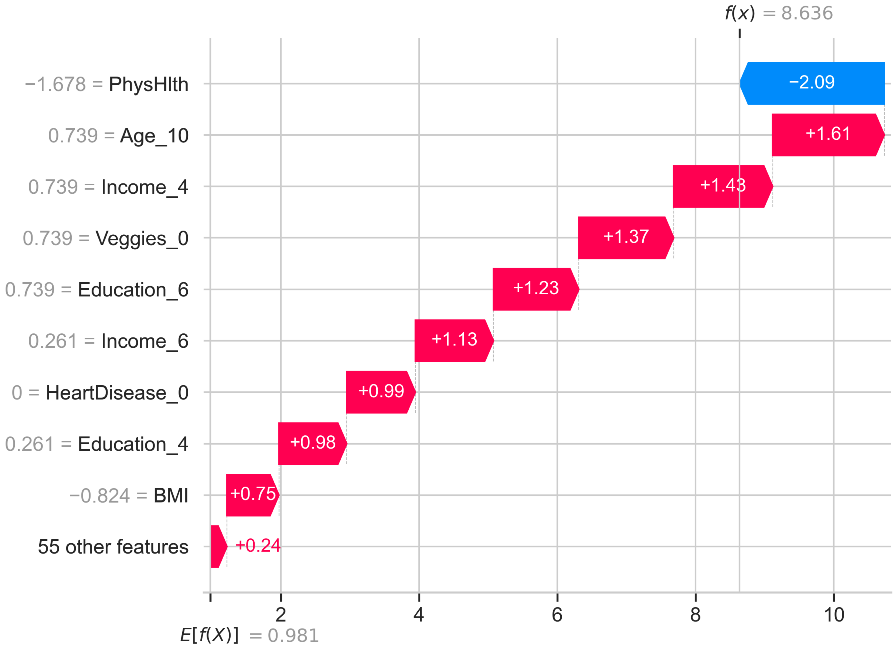


**Fig 34.** Local SHAP explanation for a high-risk prediction from the LightGBM model.

previous reports of stable and interpretable attribution patterns in boosting models [3, 9]. SHAP beeswarm plots further illustrated nonlinear and interaction-driven effects for features such as BMI and physical health indicators, reflecting the complex relationships identified in other XAI-driven stroke risk studies [19, 25].

At the individual patient level, SHAP and LIME local explanations provided actionable insights consistent with previous literature [7, 16, 17, 25]. High-risk predictions were predominantly driven by advanced age, prior heart disease, and poor

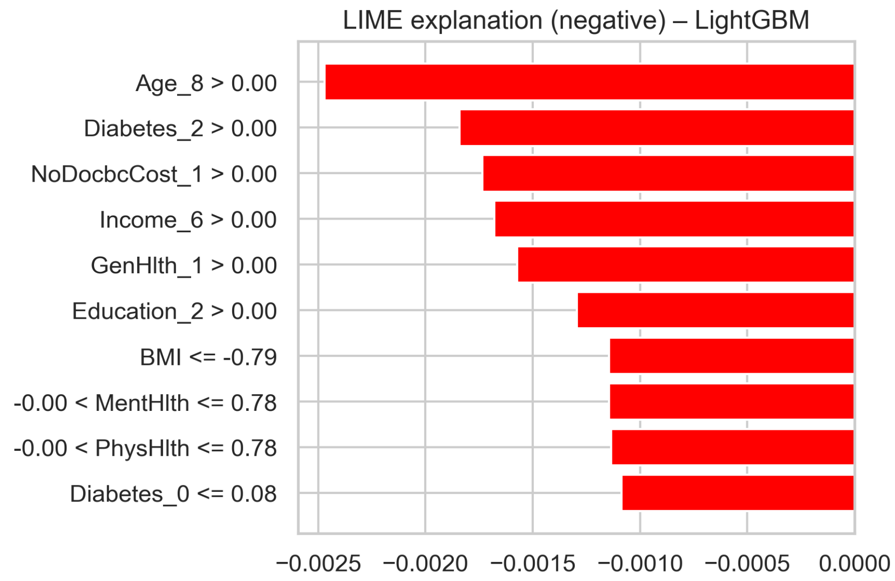


**Fig 35.** LIME explanation for a low-risk prediction from the LightGBM model. The plot shows the most influential features and their contribution to the prediction.

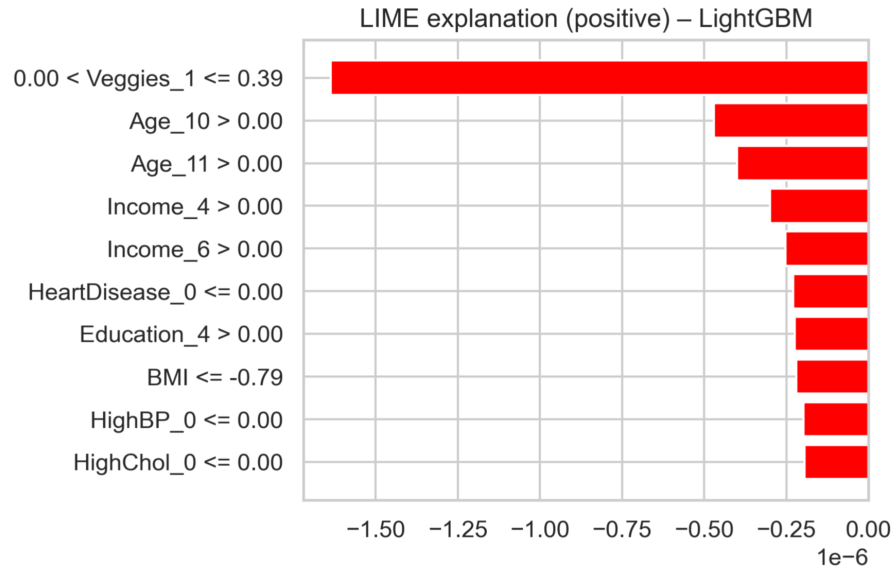


**Fig 36.** LIME explanation for a high-risk prediction from the LightGBM model.
