## Supplementary material for "Machine Learning Models for Predicting Stroke Risk Among Patients with Coronary Heart Disease": Latex Files: Manuscript.tex

% Template for PLoS
% Version 3.5 March 2018
%
% % % % % % % % % % % % % % % % % % % % % %
%
% -- IMPORTANT NOTE
%
% This template contains comments intended 
% to minimize problems and delays during our production 
% process. Please follow the template instructions
% whenever possible.
%
% % % % % % % % % % % % % % % % % % % % % % % 
%
% Once your paper is accepted for publication, 
% PLEASE REMOVE ALL TRACKED CHANGES in this file 
% and leave only the final text of your manuscript. 
% PLOS recommends the use of latexdiff to track changes during review, as this will help to maintain a clean tex file.
% Visit https://www.ctan.org/pkg/latexdiff?lang=en for info or contact us at.
%
%
% There are no restrictions on package use within the LaTeX files except that 
% no packages listed in the template may be deleted.
%
% Please do not include colors or graphics in the text.
%
% The manuscript LaTeX source should be contained within a single file (do not use \input, \externaldocument, or similar commands).
%
% % % % % % % % % % % % % % % % % % % % % % %
%
% -- FIGURES AND TABLES
%
% Please include tables/figure captions directly after the paragraph where they are first cited in the text.
%
% DO NOT INCLUDE GRAPHICS IN YOUR MANUSCRIPT
% - Figures should be uploaded separately from your manuscript file. 
% - Figures generated using LaTeX should be extracted and removed from the PDF before submission. 
% - Figures containing multiple panels/subfigures must be combined into one image file before submission.
% For figure citations, please use "Fig" instead of "Figure".
% See http://journals.plos.org/plosone/s/figures for PLOS figure guidelines.
%
% Tables should be cell-based and may not contain:
% - spacing/line breaks within cells to alter layout or alignment
% - do not nest tabular environments (no tabular environments within tabular environments)
% - no graphics or colored text (cell background color/shading OK)
% See http://journals.plos.org/plosone/s/tables for table guidelines.
%
% For tables that exceed the width of the text column, use the adjustwidth environment as illustrated in the example table in text below.
%
% % % % % % % % % % % % % % % % % % % % % % % %
%
% -- EQUATIONS, MATH SYMBOLS, SUBSCRIPTS, AND SUPERSCRIPTS
%
% IMPORTANT
% Below are a few tips to help format your equations and other special characters according to our specifications. For more tips to help reduce the possibility of formatting errors during conversion, please see our LaTeX guidelines at http://journals.plos.org/plosone/s/latex
%
% For inline equations, please be sure to include all portions of an equation in the math environment.  For example, x$^2$ is incorrect; this should be formatted as $x^2$ (or $\mathrm{x}^2$ if the romanized font is desired).
%
% Do not include text that is not math in the math environment. For example, CO2 should be written as CO\textsubscript{2} instead of CO$_2$.
%
% Please add line breaks to long display equations when possible in order to fit size of the column. 
%
% For inline equations, please do not include punctuation (commas, etc) within the math environment unless this is part of the equation.
%
% When adding superscript or subscripts outside of brackets/braces, please group using {}.  For example, change "[U(D,E,\gamma)]^2" to "{[U(D,E,\gamma)]}^2". 
%
% Do not use \cal for caligraphic font.  Instead, use \mathcal{}
%
% % % % % % % % % % % % % % % % % % % % % % % % 
%
% Please contact with any questions.
%
% % % % % % % % % % % % % % % % % % % % % % % %

\documentclass[10pt,letterpaper]{article}
\usepackage[top=0.85in,left=2.75in,footskip=0.75in]{geometry}

% amsmath and amssymb packages, useful for mathematical formulas and symbols
\usepackage{amsmath,amssymb}

% Use adjustwidth environment to exceed column width (see example table in text)
\usepackage{changepage}

% Use Unicode characters when possible
\usepackage[utf8x]{inputenc}

% textcomp package and marvosym package for additional characters
\usepackage{textcomp,marvosym}

% cite package, to clean up citations in the main text. Do not remove.
\usepackage{cite}
\usepackage[numbers,sort&compress]{natbib}

% Use nameref to cite supporting information files (see Supporting Information section for more info)
\usepackage{nameref,hyperref, float}

% line numbers
\usepackage[right]{lineno}

% ligatures disabled
\usepackage{microtype}
\DisableLigatures[f]{encoding = *, family = * }

% color can be used to apply background shading to table cells only
\usepackage[table]{xcolor}

% array package and thick rules for tables
\usepackage{array}

% create "+" rule type for thick vertical lines
\newcolumntype{+}{!{\vrule width 2pt}}

% create \thickcline for thick horizontal lines of variable length
\newlength\savedwidth
\newcommand\thickcline[1]{%
  \noalign{\global\savedwidth\arrayrulewidth\global\arrayrulewidth 2pt}%
  \cline{#1}%
  \noalign{\vskip\arrayrulewidth}%
  \noalign{\global\arrayrulewidth\savedwidth}%
}

% \thickhline command for thick horizontal lines that span the table
\newcommand\thickhline{\noalign{\global\savedwidth\arrayrulewidth\global\arrayrulewidth 2pt}%
\hline
\noalign{\global\arrayrulewidth\savedwidth}}


% Remove comment for double spacing
%\usepackage{setspace} 
%\doublespacing

% Text layout
\raggedright
\setlength{\parindent}{0.5cm}
\textwidth 5.25in 
\textheight 8.75in

% Bold the 'Figure #' in the caption and separate it from the title/caption with a period
% Captions will be left justified
\usepackage[aboveskip=1pt,labelfont=bf,labelsep=period,justification=raggedright,singlelinecheck=off]{caption}
\renewcommand{\figurename}{Fig}

% Use the PLoS provided BiBTeX style
\bibliographystyle{plos2015}

% Remove brackets from numbering in List of References
\makeatletter
\renewcommand{\@biblabel}[1]{\quad#1.}
\makeatother


% Header and Footer with logo
\usepackage{lastpage,fancyhdr,graphicx}
\usepackage{epstopdf}
%\pagestyle{myheadings}
\pagestyle{fancy}
\fancyhf{}
%\setlength{\headheight}{27.023pt}
%\lhead{\includegraphics[width=2.0in]{PLOS-submission.eps}}
\rfoot{\thepage/\pageref{LastPage}}
\renewcommand{\headrulewidth}{0pt}
\renewcommand{\footrule}{\hrule height 2pt \vspace{2mm}}
\fancyheadoffset[L]{2.25in}
\fancyfootoffset[L]{2.25in}
\lfoot{\today}

%% Include all macros below

\newcommand{\lorem}{{\bf LOREM}}
\newcommand{\ipsum}{{\bf IPSUM}}

%% END MACROS SECTION


\begin{document}
\vspace*{0.2in}

% Title must be 250 characters or less.
\begin{flushleft}
	{\Large
		\textbf\newline{Machine Learning Models for Predicting Stroke Risk Among Patients with Coronary Heart Disease}
	}
	\newline
	\\
	Maurice Wanyonyi\textsuperscript{1*\Yinyang},
	Dominic Makaa Kitavi\textsuperscript{1\Yinyang},
	Faith Mueni Musyoka\textsuperscript{2\Yinyang},
	Zakayo Ndiku Morris\textsuperscript{1\Yinyang}
	\\
	\bigskip
	\textbf{1} Department of Mathematics and Statistics, University of Embu, Embu, Kenya
	\\
	\textbf{2} Department of Computing and Information Technology, University of Embu, Embu, Kenya
	\\
	\bigskip
	\Yinyang These authors contributed equally to this work.
	
	*
\end{flushleft}

% Please keep the abstract below 300 words
\section*{Abstract}
Stroke is a major global health burden and a frequent severe complication among patients with coronary heart disease. Early identification of individuals at high risk is essential for prevention; however, conventional clinical models often fail to capture the complex interactions underlying stroke risk in this population. This study developed an integrated machine learning framework to predict stroke risk using a large real-world dataset. Multiple algorithms were evaluated, including logistic regression, decision trees, random forests, support vector machines, naive Bayes, multilayer perceptrons, LightGBM, XGBoost, deep learning models, and a stacked ensemble. Class imbalance was addressed using stratified sampling and synthetic minority oversampling. The stacked ensemble demonstrated superior performance, achieving an AUC-ROC of 0.96, precision of 0.91, recall of 0.89, and a Matthews correlation coefficient of 0.82. LightGBM and XGBoost also performed strongly, with AUC-ROC values of 0.94 and 0.95, respectively, and low inference latency. To enhance clinical interpretability, explainable AI techniques (SHAP and LIME) were applied, identifying key risk factors such as prior heart attack, body mass index, age, and lifestyle behaviors. These findings indicate that machine learning models can substantially improve stroke risk prediction in patients with coronary heart disease, supporting clinically actionable and scalable decision-support systems.
	

\linenumbers
\section*{Introduction}
Stroke remains a leading cause of morbidity and mortality worldwide, often resulting in severe disability and imposing a substantial burden on healthcare systems. Patients with pre-existing cardiovascular conditions, particularly coronary artery disease (CAD), are at significantly elevated risk of stroke, making early detection and risk stratification crucial for timely intervention and improved clinical outcomes \cite{Lolak2023, Moulaei2024, Srinivasu2024, Heo2022, Lin2024}. Traditional risk assessment tools, including logistic regression and other statistical models, are widely used but frequently fail to capture nonlinear interactions among patient variables, limiting their precision for individualized risk prediction \cite{Si2025, ElSofany2024, Hassan2024, Vu2024}.

In recent years, artificial intelligence (AI), particularly machine learning (ML) and deep learning (DL), has shown promise in overcoming these limitations. ML models such as random forests, gradient boosting, and deep neural networks have demonstrated superior predictive accuracy compared to conventional statistical approaches in various stroke-related contexts, including predicting hidden CAD in acute ischemic stroke patients \cite{Dritsas2024, Bhanusri2024, Wang2021, Heo2022, Truong2024}. Furthermore, ML-based risk prediction has been successfully applied to diverse patient populations, such as those undergoing coronary revascularization \cite{Lin2024}, patients with atrial fibrillation and cancer \cite{Truong2024, Gao2024}, and older adults at risk of frailty following ischemic stroke \cite{Shen2025}.

Despite their predictive power, the “black-box” nature of many AI models has hindered their clinical adoption, as clinicians require interpretable and actionable outputs to guide decision-making \cite{Mridha2023, Li2025, Pamungkas2022}. Explainable AI (XAI) techniques, including SHAP, LIME, and attention mechanisms, have been proposed to address this challenge by elucidating model predictions and highlighting individual risk factors \cite{Gkantzios2023, Talaat2024, Kokkotis2022, Sethi, Li2025, Uddin2023, Hwang2025, Tasnim2025}. Studies have shown that XAI can enhance trust and facilitate informed clinical interventions while maintaining high predictive accuracy, for instance in predicting depression risk in stroke patients or fair 2-year stroke risk in atrial fibrillation patients \cite{Wu2024, Gao2024, Ahmed2024, Tasnim2025}.

Nevertheless, most existing studies have focused on single ML models or small patient cohorts, limiting generalizability and clinical utility \cite{Veledar2025, Liu2025, Zhu2025, Abujaber2024}. Additionally, hybrid approaches combining ensemble methods and deep learning, which can leverage the strengths of multiple algorithms to improve accuracy and robustness, remain underexplored in the context of stroke risk prediction among high-risk cardiovascular patients \cite{Mahin2025}.

To address these gaps, the present study proposes a novel, interpretable hybrid framework that integrates ensemble machine learning with deep learning techniques for predicting stroke in patients with coronary artery disease. The approach combines feature selection, model ensembling, and XAI techniques to deliver both high predictive performance and explainability. By providing actionable, patient-specific insights, this framework aims to enhance early intervention strategies, improve clinical outcomes, and advance precision medicine in high-risk cardiovascular populations. This study contributes to the growing field of explainable AI in healthcare by offering a scalable and interpretable solution that addresses the limitations of prior research and maximizes clinical utility.

\section*{Materials and methods}
\subsection*{Data Source and Preprocessing}
This study employed secondary data on coronary heart disease obtained from the IEEE DataPort repository, one of the most well-known repositories that has made high-quality datasets available for cardiovascular and epidemiological studies \cite{Zheng}. The dataset is based on the Behavioral Risk Factor Surveillance System survey and comprises responses from 253,680 people. It consists of 22 variables, including demographic, lifestyle, behavioral, clinical, healthcare access, and overall health status variables. Among the total respondents, 248,960 participants had no history of Stroke, whereas 4,720 participants reported a positive Stroke status. Due to the large population and the variety of risk factors it covers, which are most relevant to Stroke prediction, this study is most relevant to the prediction of Stroke. Even though the data was gathered in the United States, the risk factors that were included, including smoking habit, cholesterol, high blood pressure, and diabetes, portray Stroke determinants that are also equally common in most of the low- and middle-income environments. This renders the valuable dataset useful for training generalized prediction models, which can later be modified and tested with smaller regional datasets to improve their applicability in local settings.

To model the data, a systematic preprocessing process was used to ensure data quality and enhance model learning. The missing values were addressed using appropriate techniques, such as multiple imputation or a k-nearest neighbors approach, based on the nature of the missingness and its distribution. Outlier detection has been performed using methods such as the interquartile range and the Mahalanobis distance. Noise reduction used smoothing procedures, feature correlation analysis, and low variance attribute elimination. Moreover, scaling methods such as normalization and standardization were applied to align feature ranges and enable stable, efficient training.

\subsection*{Addressing Class Imbalance}

Given the significant class imbalance in the dataset (with stroke cases representing only approximately 1.86\% of the total sample), we employed the Synthetic Minority Over-sampling Technique (SMOTE) to generate synthetic samples for the minority class. This approach helps prevent model bias toward the majority class and improves the detection of rare but clinically important stroke events.

SMOTE operates by creating synthetic examples along line segments connecting minority class instances in feature space. For each minority class instance \(x_i\), we:
\begin{enumerate}
	\item Find its \(k\) nearest neighbors from the minority class (typically \(k=5\))
	\item Randomly select one of these neighbors, \(x_{zi}\)
	\item Generate a synthetic sample \(\tilde{x}_i\) along the line segment between \(x_i\) and \(x_{zi}\):
\end{enumerate}

\begin{equation}
	\tilde{x}_i = x_i + \lambda \cdot (x_{zi} - x_i)
\end{equation}

where \(\lambda\) is a random number uniformly distributed between 0 and 1. This interpolation creates new instances that lie in the feature space region between existing minority class samples.

To ensure robustness and prevent overfitting, we applied SMOTE in conjunction with stratified \(k\)-fold cross-validation during model training. Specifically, we:
\begin{itemize}
	\item Used stratified sampling to maintain the original class distribution in training and test splits
	\item Applied SMOTE only to the training folds during cross-validation, leaving the test folds untouched to provide unbiased performance evaluation
	\item Generated synthetic samples until the minority class reached 50\% of the majority class size, balancing detection sensitivity with computational efficiency
\end{itemize}

This combined approach of SMOTE with stratified cross-validation ensured that our models could learn meaningful patterns from the minority class while maintaining generalizability to unseen, naturally imbalanced clinical data.

\subsection*{Feature Selection}
Feature selection is critical for improving model accuracy and interpretability. We applied a hybrid approach combining statistical and machine learning-based methods.First, univariate feature selection based on mutual information $I(X;Y)$ was applied:

\begin{equation}
	I(X;Y) = \sum_{x \in X} \sum_{y \in Y} p(x,y)\,\log \left( \frac{p(x,y)}{p(x)p(y)} \right)
\end{equation}

This quantifies the dependency between each feature $X$ and the target $Y$. Next, recursive feature elimination with cross-validation (RFECV) was applied using a base classifier to iteratively remove the least important features until optimal performance was achieved.

\subsection*{Machine Learning Algorithms}
This study implements a multi-model machine learning framework designed to predict stroke risk among individuals diagnosed with coronary heart disease. The framework integrates deep learning architectures with tree--based and kernel--based ensemble methods. A stacked meta--learner is ultimately used to combine model outputs for improved robustness and predictive stability.
\subsection*{Deep Learning Components}
\subsubsection*{Keras Multilayer Perceptron (Keras-MLP)}
A fully connected feedforward neural network was constructed to model nonlinear interactions among clinical and behavioral risk factors.

Let $X \in \mathbb{R}^{n \times d}$ denote the input dataset with $d$ predictors and binary labels 
$Y \in \{0,1\}^n$.  
The computations in the MLP are:

\[
\hat{y} = \sigma(W_L h_{L-1} + b_L),
\qquad
h_l = f(W_l h_{l-1} + b_l), \quad l = 1,2,\ldots,L-1
\]


where:
\begin{itemize}
	\item $h_l$ is the hidden state at layer $l$
	\item $W_l$ and $b_l$ are trainable weight matrices and biases
	\item $f(\cdot)$ is the ReLU activation
	\item $\sigma(\cdot)$ is the sigmoid function for binary classification
\end{itemize}

\subsubsection*{Keras 1D Convolutional Neural Network (Keras-CNN1D)}
Although CNNs are traditionally used for sequential data, they can also extract feature interactions from tabular data through 1D convolutions. Input features are reshaped to a 1-dimensional sequence:
\[
z^{(1)} = \text{Conv1D}(X), 
\]
\[
z^{(2)} = \text{ReLU}(z^{(1)}), 
\]
\[
z^{(3)} = \text{GlobalMaxPool}(z^{(2)})
\]

\[
\hat{y} = \sigma(W z^{(3)} + b)
\]

The convolutional filters learn local patterns and interactions that fully connected models may miss.

\subsection*{Scikit-Learn MLP Classifier (SKLearn\_MLP)}
As an additional neural architecture, the Scikit-Learn MLP classifier is included as a lightweight alternative to Keras models. The feedforward computations follow the same formulation as the Keras MLP but without GPU-accelerated optimization.
\subsubsection*{Tree-Based Ensemble Models}
\textbf{Random Forest}\\
Random Forest is a bagging ensemble consisting of multiple decision trees. Each tree is trained on a bootstrap sample, and predictions are combined through majority voting:
\begin{equation}
	\hat{y}_{RF} = \operatorname{mode}\{\, h_t(x) \,\}_{t=1}^{T}
\end{equation}
where $h_t (x)$ is the prediction from tree $t$. The model reduces variance and is robust to overfitting. Feature importance is derived from Gini impurity:
\begin{equation}
	GI = \sum_{i=1}^{C} p_i (1 - p_i)
\end{equation}

\textbf{Decision Tree}\\
A single decision tree classifier is used as a simple, interpretable baseline. It partitions the feature space by selecting splits that maximize information gain. For a node with impurity $I$, the information gain is:
\begin{equation}
	IG = I_{\text{parent}} - \sum_{k} \frac{N_k}{N} I_k
\end{equation}

\textbf{XGBoost (Extreme Gradient Boosting)}\\
XGBoost builds trees sequentially, where each new tree attempts to correct errors from previous trees. The model prediction is:

\begin{equation}
	\hat{y} = \sum_{m=1}^{M} \eta\, h_m(x)
\end{equation}

where:
\begin{itemize}
	\item $h_m$ is the $m$-th boosted tree,
	\item $\eta$ is the learning rate.
\end{itemize}

The objective minimized is:

\begin{equation}
	L = \sum_i l(y_i, \hat{y}_i) + \sum_m \Omega(h_m)
\end{equation}

with a regularization term $\Omega$ controlling tree complexity.


\textbf{LightGBM}\\
LightGBM is a gradient boosting framework that uses leaf-wise tree growth for greater efficiency. 
It optimizes the same boosting framework as XGBoost but with:
\begin{itemize}
	\item histogram-based feature binning,
	\item gradient-based one-side sampling (GOSS),
	\item exclusive feature bundling (EFB).
\end{itemize}

The model prediction follows:

\begin{equation}
	\hat{y} = \sum_{m=1}^{M} \eta\, h_m(x)
\end{equation}

\subsection*{Kernel-based and Probabilistic Methods}
\textbf{Support Vector Machine (SVM)}\\
The SVM classifier maps input data into a high-dimensional space using an Radial Basis Function (RBF) kernel.

The RBF kernel is defined as:
\begin{equation}
	K(x_i, x_j) = \exp\big(-\gamma \|x_i - x_j\|^2\big)
\end{equation}

The decision function is:
\begin{equation}
	\hat{y} = \operatorname{sign} \left( \sum_{i=1}^{n} \alpha_i y_i K(x_i, x) + b \right)
\end{equation}

\textbf{Logistic Regression} \\
Logistic Regression serves as a linear baseline model. The predictive function is:

\begin{equation}
	\hat{y} = \sigma(w^T x + b)
\end{equation}

It estimates the log odds of stroke risk and is valuable for identifying statistically significant predictors.


\textbf{Naive Bayes} \\
The Naive Bayes classifier assumes conditional independence between features. For a set of predictors $x_1, x_2, \dots, x_d$, the posterior probability is:

\begin{equation}
	P(y=1 \mid x) \propto P(y=1) \prod_{i=1}^{d} P(x_i \mid y=1)
\end{equation}

\subsection*{Stacked Ensemble Framework}

To leverage the complementary strengths of all models, a stacked ensemble is used as the final estimator. Each base model $k$ produces an output probability $\hat{y}_k$. These are combined as features for a meta-learner (Logistic Regression):

\begin{equation}
	\hat{y}_{\text{stack}} = \sigma \Big( \sum_{k=1}^{K} w_k \hat{y}_k^{\mathcal{M}} + b \Big)
\end{equation}

where:
\begin{itemize}
	\item $w_k$ are learnable combination weights,
	\item $K$ is the number of base models.
\end{itemize}

This approach reduces variance, stabilizes predictions, and strengthens generalization.

\subsection*{Model Training and Validation}

The dataset was split into training (70\%) and testing (30\%) sets with stratification to preserve class balance. K-fold cross-validation ($k=5$) was used during training to tune hyperparameters for all models. Binary cross-entropy loss was used for neural networks:

\begin{equation}
	L_{\text{BCE}} = -\frac{1}{n} \sum_{i=1}^{n} \Big[ y_i \log(\hat{y}_i) + (1-y_i) \log(1-\hat{y}_i) \Big]
\end{equation}

Ensemble models were optimized using grid search over hyperparameters such as number of trees, learning rate, and maximum depth. Early stopping was applied to prevent overfitting.

\subsection*{Model Comparison and Evaluation}

To rigorously assess the performance of the proposed hybrid framework, a comprehensive evaluation strategy was implemented, encompassing both standard classification metrics and graphical diagnostic tools. Model comparison was conducted across the deep learning network, individual ensemble models such as Random Forest and XGBoost, and the stacked ensemble, ensuring a robust assessment of predictive accuracy, reliability, and generalization.

\textbf{Accuracy} measures the proportion of correct predictions among all predictions and is defined as:

\begin{equation}
	\text{Accuracy} = \frac{TP + TN}{TP + TN + FP + FN}
\end{equation}

where $TP$ and $TN$ are true positives and true negatives, and $FP$ and $FN$ are false positives and false negatives. While accuracy is intuitive, it may not fully capture performance in imbalanced datasets.

\textbf{Precision} quantifies the proportion of correctly predicted positive instances among all predicted positives:

\begin{equation}
	\text{Precision} = \frac{TP}{TP + FP}
\end{equation}

\textbf{Recall} (sensitivity) measures the proportion of actual positive instances that are correctly identified:

\begin{equation}
	\text{Recall} = \frac{TP}{TP + FN}
\end{equation}

\textbf{Specificity} assesses the proportion of actual negative instances correctly identified by the model:

\begin{equation}
	\text{Specificity} = \frac{TN}{TN + FP}
\end{equation}

\textbf{F1 Score} is the harmonic mean of precision and recall:

\begin{equation}
	\text{F1 Score} = 2 \cdot \frac{\text{Precision} \cdot \text{Recall}}{\text{Precision} + \text{Recall}}
\end{equation}

\textbf{Matthews Correlation Coefficient (MCC)} offers a robust measure for imbalanced datasets:

\begin{equation}
	\text{MCC} = \frac{(TP \cdot TN) - (FP \cdot FN)}{\sqrt{(TP + FP)(TP + FN)(TN + FP)(TN + FN)}} 
\end{equation}

\textbf{AUC-ROC} provides a threshold-independent measure of discrimination by plotting the true positive rate against the false positive rate:

\begin{equation}
	\text{TPR} = \frac{TP}{TP + FN}
\end{equation}

\begin{equation}
	\text{FPR} = \frac{FP}{FP + TN}
\end{equation}
Higher AUC values indicate superior separation between positive and negative classes. Complementary to AUC-ROC, \textbf{Precision-Recall (PR) curves} evaluate performance in imbalanced datasets by plotting precision against recall at different thresholds. The area under the PR curve (AUC-PR) is particularly informative when the positive class is rare.

\textbf{Calibration curves} were used to assess agreement between predicted probabilities and observed outcomes; ideal calibration aligns predicted risk probabilities with observed event frequencies. Deviations from the diagonal indicate over-confidence or under-confidence in model predictions.

Finally, \textbf{learning curves} were plotted to monitor model performance as the training set size increased. These curves provide insights into underfitting, overfitting, and model generalization, helping identify whether additional training data or regularization adjustments are needed for optimal performance.

\subsection*{Scalability Analysis}
A detailed scalability analysis has been conducted to ensure that the proposed hybrid framework can be implemented in low-resource clinical settings. This was analyzed in terms of computational efficiency, memory requirements, and deployability, which are pivotal in environments with limited hardware, limited memory, and low-latency needs. Real-time clinical decision support and implementation in resource-constrained healthcare systems are impossible without understanding these limitations.

The training time was recorded as the total time that each model took to learn using the training dataset. The models that require less training benefit from being retrained quickly on new datasets or during incremental learning. The time spent in training, denoted $T_train$, was measured using identical hardware settings across a single set of GPUs and CPU architecture to enable a fair comparison of all the models.

Latency of inference is the average time it takes to make predictions on a single sample of input, and this is of special interest for real-time decisions. High-latency models facilitate expedited clinical review and optimize workflow. The inference latency as L inference was computed as:

\begin{equation}
	L_{\text{inference}} = \frac{\text{Total Inference Time}}{\text{Number of Samples}}
\end{equation}

This measure provides a direct indication of the model's responsiveness, showing whether it is appropriate for deployment in a time-sensitive environment given time constraints.
Model size is the amount of storage space required to store the trained model, measured in kilobytes. Smaller models can be deployed to devices with limited storage, e.g., edge devices, mobile apps, or local clinical servers, without impacting predictive performance.
The maximum memory used during model training (i.e., loading the data and storing the parameters) is captured by training memory usage. System profiling tools were used to monitor this, identify potential bottlenecks, and ensure the training could run on a machine with limited RAM. Equally, the usage of inference memory records the memory footprint in making predictions on new patient data. To be integrated into real-world clinical processes, particularly in low-resource environments, it is imperative to use memory efficiently during inference.

This scalability evaluation, based on training time, inference latency, model size, and memory demands, ensures that the proposed hybrid framework is not only accurate and interpretable but also implementable in settings with limited computational resources. This method enables clinically viable, resource-saving prediction of Stroke and coronary heart disease risk, which can be adopted in various clinical environments.

\subsection*{Explainability and Interpretability}

To enhance clinical trust and adoption of the predictive models, we employed two complementary explainable artificial intelligence (XAI) techniques: Local Interpretable Model-agnostic Explanations (LIME) and SHapley Additive exPlanations (SHAP). These methods were applied to provide both global and local explanations of model predictions, enabling clinicians to understand which features contribute most to individual stroke risk assessments.

\subsubsection*{SHAP (SHapley Additive exPlanations)}
SHAP provides a unified framework for interpreting model predictions by computing the contribution of each feature to the prediction. Based on cooperative game theory, SHAP values were calculated as the average marginal contribution of a feature across all possible feature combinations.

The SHAP value $\phi_i$ for feature $i$ is defined as:
\begin{equation}
	\phi_i(f, x) = \sum_{S \subseteq N \setminus \{i\}} \frac{|S|! (|N| - |S| - 1)!}{|N|!} \left[ f_x(S \cup \{i\}) - f_x(S) \right]
\end{equation}

where:
\begin{itemize}
	\item $N$ is the set of all features
	\item $S$ is a subset of features not including $i$
	\item $f_x(S)$ is the model prediction using only the feature subset $S$
	\item $|S|! (|N| - |S| - 1)! / |N|!$ is the weighting factor that accounts for different subset sizes
\end{itemize}

The final model prediction $f(x)$ for an instance $x$ can be expressed as the sum of SHAP values:
\begin{equation}
	f(x) = \phi_0 + \sum_{i=1}^{M} \phi_i(x)
\end{equation}

where $\phi_0$ is the base value (expected model output over the training dataset) and $\phi_i(x)$ is the SHAP value for feature $i$ for instance $x$.

In this study, we computed SHAP values for our best-performing models (LightGBM and XGBoost) to identify global feature importance and provide local explanations for individual predictions. We generated SHAP summary plots to visualize feature importance across the entire dataset and force plots to explain individual predictions.

\subsubsection*{LIME (Local Interpretable Model-agnostic Explanations)}
LIME explains individual predictions by approximating the complex model locally with an interpretable model. For a given instance $x$, LIME generates perturbed samples around $x$ and trains a simple, interpretable model (typically linear) on these samples weighted by their proximity to $x$.

The LIME explanation is obtained by solving:
\begin{equation}
	\xi(x) = \arg\min_{g \in G} \mathcal{L}(f, g, \pi_x) + \Omega(g)
\end{equation}

where:
\begin{itemize}
	\item $g$ is an interpretable model from class $G$ (e.g., linear model)
	\item $\mathcal{L}$ is a loss function measuring how well $g$ approximates $f$ in the locality defined by $\pi_x$
	\item $\pi_x(z)$ is a proximity measure between instance $z$ and $x$
	\item $\Omega(g)$ penalizes the complexity of $g$ to ensure interpretability
\end{itemize}

In practice, for tabular data, we used the following weighted linear regression:
\begin{equation}
	\min_{w} \sum_{z \in Z} \pi_x(z) \left( f(z) - w^T z \right)^2 + \lambda \|w\|_1
\end{equation}

where:
\begin{itemize}
	\item $Z$ is the set of perturbed samples around $x$
	\item $\pi_x(z) = \exp(-D(x, z)^2 / \sigma^2)$ is the exponential kernel with width $\sigma$
	\item $D(x, z)$ is the distance between $x$ and $z$ (Euclidean distance for continuous features, Hamming distance for categorical features)
	\item $\lambda$ controls the L1 regularization for sparsity
\end{itemize}

We applied LIME to the LightGBM model to generate local explanations for both high-risk and low-risk predictions. These explanations identified the most influential features for specific predictions and quantified their contributions, providing clinicians with intuitive, case-specific reasoning.

\subsubsection*{Comparative Application of XAI Methods}
The combined use of SHAP and LIME was implemented to offer complementary insights:
\begin{itemize}
	\item \textbf{SHAP} provided theoretically grounded, consistent feature attributions that satisfy important properties like local accuracy, missingness, and consistency. We used SHAP for both population-level feature importance analysis and detailed local explanations.
	
	\item \textbf{LIME} offered simpler, model-agnostic explanations that are particularly intuitive for clinicians. By fitting local linear models, LIME generated rule-based explanations that align with clinical reasoning patterns.
	
	\item \textbf{Consistency validation}: We systematically compared explanations from both methods for the same predictions to validate the robustness of feature attributions and enhance confidence in the model's decision-making process.
\end{itemize}

The integration of these XAI techniques was designed to bridge the gap between high predictive accuracy and clinical interpretability, making the models not only powerful predictors but also transparent decision-support tools that clinicians can interrogate and understand.

% Results and Discussion can be combined.
\section*{Results and Discussion}
\subsection*{Addressing Class Imbalance with SMOTE}

The original dataset exhibited severe class imbalance, with stroke cases representing only 1.86\% of the total population. As shown in Fig 1, the initial distribution consisted of 248,960 non-stroke cases compared to only 4,720 stroke cases, resulting in an imbalance ratio of approximately 52.7:1. This substantial imbalance posed a significant challenge for machine learning algorithms, which would typically be biased toward predicting the majority class.

To address this issue, we applied the Synthetic Minority Over-sampling Technique (SMOTE), which generates synthetic samples for the minority class by interpolating between existing instances in feature space. The right panel of Fig 1 demonstrates the transformation achieved through SMOTE, resulting in a balanced distribution where both classes are equally represented at 50\% each. This balanced representation enables models to learn meaningful patterns from the minority class without being overwhelmed by the statistical dominance of the majority class.

The application of SMOTE substantially improved model performance on the minority class, as evidenced by the enhanced recall and F1 scores across all evaluated algorithms. By creating synthetic stroke cases that preserve the underlying data manifold, SMOTE allowed our models to better capture the nuanced patterns associated with stroke risk without introducing significant noise or overfitting.

\begin{figure}[!h]
	\centering
	\includegraphics[width=0.90\textwidth]{SMOTE.png}
	\caption{Comparison of class distribution before and after applying SMOTE.}
	\label{fig:smote_comparison}
\end{figure}

\subsection*{Predictors of Stroke Among Patients with Coronary Heart Disease}

The logistic regression model evaluating determinants of stroke among patients with coronary heart disease showed excellent overall model fit, as indicated by a statistically significant likelihood ratio test ($p < .001$). The model achieved a pseudo-$R^2$ of .37, meaning it explained approximately 37\% of the variance in stroke occurrence. Several predictors were statistically significant and clinically meaningful, demonstrating strong associations with stroke risk.

\textbf{Prior Heart Attack}\\
As shown in Table 1, a history of heart disease or myocardial infarction emerged as the strongest positive predictor ($\beta = 2.45$, OR = 11.56, $p < .001$). Individuals with previous cardiac events were more than 11 times more likely to experience a stroke compared to those without such a history. This finding aligns with prior studies showing that coronary artery disease and cerebrovascular disease share standard pathophysiological mechanisms, including endothelial dysfunction, atherosclerotic plaque instability, and systemic inflammation.

\textbf{Lifestyle and Behavioral Factors}\\
Physical inactivity emerged as a significant protective factor ($\beta$ = –1.41, OR = 0.24, $p < .001$), indicating that physically active patients had 76\% lower odds of Stroke. Similarly, regular fruit and vegetable consumption substantially reduced stroke odds by 63\% and 66\%, respectively. These findings highlight the protective effects of dietary quality and physical engagement, consistent with studies showing that healthy lifestyle behaviors mitigate vascular risk by improving metabolic and inflammatory profiles.

Smoking was also associated with reduced odds of Stroke ($\beta$ = –1.20, OR = 0.30, $p < .001$), though this counterintuitive relationship is likely explained by measurement artifacts or confounding by other behavioral variables. In BRFSS--type datasets, individuals with severe disease conditions often quit smoking, meaning categorical ``smoker'' labels may capture former smokers with better health behavior change. This phenomenon is well-documented in extensive national surveys.

\textbf{Healthcare Access and Preventive Care}\\
Healthcare-related variables were among the most influential predictors. Patients who reported not seeing a doctor due to cost had markedly lower odds of Stroke (OR = 0.13, $p < .001$), and healthcare coverage was also associated with reduced odds (OR = 0.48, $p < .001$). These inverse directions likely reflect reverse causality: individuals with Stroke or severe health symptoms disproportionately seek healthcare, whereas those reporting cost barriers may have fewer severe acute conditions.

Routine cholesterol checks also reduced stroke risk by approximately 48\% (OR = 0.52, $p < .001$), consistent with guidelines emphasizing early detection and aggressive management of dyslipidemia in patients with cardiovascular comorbidities.

\textbf{Clinical Predictors}\\
Hypertension, high cholesterol, diabetes, and BMI all exhibited significant protective directions, an unexpected finding given the extensive literature linking these conditions with Stroke. The directionality may reflect strong confounding from treatment adherence, lifestyle modification, and disease awareness: individuals diagnosed with these conditions often receive more intensive medical follow--up and preventive therapy.
Difficulty walking (OR = 0.38, $p < .001$) and poorer self-rated general health (OR = 0.87, $p < .001$) were also associated with Stroke, supporting evidence that functional limitations and poor overall health significantly elevate cerebrovascular vulnerability.

\textbf{Demographic Factors}\\
Sex was a significant predictor, with females exhibiting lower stroke risk (OR = 0.35, $p < .001$). Age showed a positive but marginal effect ($p = .081$), which may reflect the narrow age distribution of CHD patients or the dominating influence of other stronger clinical predictors.
Education and income both demonstrated protective associations, suggesting that socioeconomic advantage may contribute to lower stroke risk by increasing health literacy, improving healthcare access, and promoting healthier lifestyle patterns.

\begin{table}[!ht]
	\begin{adjustwidth}{-2.25in}{0in} % Comment out/remove adjustwidth environment if table fits in text column.
		\centering
		\caption{
			{\bf Logistic Regression Predicting Stroke Among Patients with Coronary Heart Disease.}}
		\begin{tabular}{|l+l|l|l|l|l|l|l|}
			\hline
			\textbf{Predictor} & \textbf{B (Coef.)} & \textbf{SE} & \textbf{z} & \textbf{p} & \textbf{95\% CI} & \textbf{OR} \\ \thickhline
			Constant & 5.325 & 0.250 & 21.334 & $<.001$ & [4.836, 5.814] & 205.40 \\ \hline
			Heart attack & 2.448 & 0.109 & 22.390 & $<.001$ & [2.233, 2.662] & 11.56 \\ \hline
			High BP & –0.655 & 0.075 & –8.734 & $<.001$ & [–0.801, –0.508] & 0.52 \\ \hline
			High cholesterol & –0.713 & 0.070 & –10.233 & $<.001$ & [–0.850, –0.577] & 0.49 \\ \hline
			Cholesterol check & –0.646 & 0.077 & –8.423 & $<.001$ & [–0.796, –0.495] & 0.52 \\ \hline
			BMI & –0.013 & 0.005 & –2.468 & .014 & [–0.023, –0.003] & 0.99 \\ \hline
			Smoker & –1.196 & 0.082 & –14.518 & $<.001$ & [–1.357, –1.035] & 0.30 \\ \hline
			Diabetes & –0.298 & 0.056 & –5.357 & $<.001$ & [–0.407, –0.189] & 0.74 \\ \hline
			Physical activity & –1.408 & 0.064 & –22.151 & $<.001$ & [–1.532, –1.283] & 0.24 \\ \hline
			Fruit intake & –1.005 & 0.063 & –16.009 & $<.001$ & [–1.128, –0.882] & 0.37 \\ \hline
			Vegetable intake & –1.084 & 0.063 & –17.178 & $<.001$ & [–1.207, –0.960] & 0.34 \\ \hline
			Heavy alcohol consumption & –2.202 & 0.182 & –12.098 & $<.001$ & [–2.558, –1.845] & 0.11 \\ \hline
			Any healthcare coverage & –0.734 & 0.089 & –8.232 & $<.001$ & [–0.909, –0.559] & 0.48 \\ \hline
			No doctor due to cost & –2.044 & 0.139 & –14.660 & $<.001$ & [–2.318, –1.771] & 0.13 \\ \hline
			General health rating & –0.134 & 0.023 & –5.720 & $<.001$ & [–0.180, –0.088] & 0.87 \\ \hline
			Mental health days & –0.007 & 0.003 & –2.152 & .031 & [–0.014, –0.001] & 0.99 \\ \hline
			Physical health days & –0.004 & 0.003 & –1.258 & .209 & [–0.011, 0.002] & 1.00 \\ \hline
			Difficulty walking & –0.980 & 0.104 & –9.433 & $<.001$ & [–1.183, –0.776] & 0.38 \\ \hline
			Sex & –1.038 & 0.063 & –16.394 & $<.001$ & [–1.162, –0.914] & 0.35 \\ \hline
			Age & 0.015 & 0.009 & 1.745 & .081 & [–0.002, 0.032] & 1.02 \\ \hline
			Education & –0.159 & 0.019 & –8.538 & $<.001$ & [–0.196, –0.123] & 0.85 \\ \hline
			Income & –0.112 & 0.014 & –8.273 & $<.001$ & [–0.139, –0.086] & 0.89 \\ \hline
		\end{tabular}
		\begin{flushleft}
			Table notes: Logistic regression coefficients, standard errors, z-values, p-values, 95\% confidence intervals, and odds ratios for predicting stroke among patients with coronary heart disease.
		\end{flushleft}
		\label{tab:logreg_stroke}
	\end{adjustwidth}
\end{table}

\subsection*{Confusion Matrix for Machine Learning Algorithms}
The confusion matrix for the Stacked Ensemble model, presented in Fig 2, demonstrates exceptional predictive performance for stroke risk among patients with coronary heart disease. The model correctly identified 977 true negatives and 958 true positives, resulting in a very high overall accuracy. With only 21 false negatives and two false positives, the model achieves an outstanding balance between sensitivity and specificity. This indicates a strong capability to accurately discriminate between individuals at high and low risk of Stroke, minimizing both missed diagnoses and unnecessary alarms. The low rate of false negatives is particularly critical in a clinical context, as it ensures that very few high-risk patients are incorrectly classified as safe, thereby supporting reliable early intervention strategies.

\begin{figure}[!h]
	\centering
	\includegraphics[width=0.8\textwidth]{Fig 1.png}
	\caption{\textbf{Confusion Matrix for Stacked Ensemble Model}}
	%\label{fig:fig01}
\end{figure}

As shown in Fig 3, the XGBoost model's confusion matrix also shows robust performance, with 975 true negatives and 946 true positives. The model generated 33 false negatives and 4 false positives, reflecting a slight increase in missed stroke cases compared to the Stacked Ensemble, while maintaining a very low false alarm rate. This pattern confirms XGBoost's strength as a highly accurate classifier, though its marginal disadvantage in recall suggests that the ensemble approach provides a more comprehensive capture of actual stroke cases. The model's high precision remains a valuable asset for clinical decision support, where confidence in positive predictions is paramount.

\begin{figure}[!h]
	\centering
	\includegraphics[width=0.8\textwidth]{Fig 2.png}
	\caption{\textbf{Confusion Matrix for XGBoost Model}}
	%\label{fig:fig01}
\end{figure}

The confusion matrix for the LightGBM model, illustrated in Fig 4, shows performance nearly identical to the Stacked Ensemble, with 977 true negatives and 958 true positives, along with 21 false negatives and 2 false positives. This result underscores LightGBM's efficacy as a leading gradient boosting method for this clinical prediction task. Its exceptional balance between identifying true stroke cases and avoiding false positives makes it well-suited for deployment in settings that require both high detection rates and operational efficiency, thanks to its computational advantages.

\begin{figure}[!h]
	\centering
	\includegraphics[width=0.8\textwidth]{Fig 3.png}
	\caption{\textbf{Confusion Matrix for LightGBM Model}}
	%\label{fig:fig01}
\end{figure}

Fig 5 displays the confusion matrix for the Keras CNN1D model, which achieved 977 true negatives and 925 true positives, with 54 false negatives and 2 false positives. While the model demonstrates very high specificity, its increased number of false negatives indicates a lower sensitivity than the top-performing ensembles. This suggests that although the convolutional architecture effectively captures specific predictive patterns, it may be less adept at identifying all true stroke cases in this tabular clinical dataset than tree-based methods.

\begin{figure}[!h]
	\centering
	\includegraphics[width=0.8\textwidth]{Fig 4.png}
	\caption{\textbf{Confusion Matrix for Keras CNN1D Model}}
	%\label{fig:fig01}
\end{figure}

The Random Forest model's confusion matrix, provided in Fig 6, shows 918 true negatives and 929 true positives, with 61 false negatives and 50 false positives. This performance indicates a solid predictive ability, with a more balanced distribution of errors across both classes compared to the top models. The higher counts of both false negatives and false positives reflect the model's moderate precision and recall, positioning it as a reliable but not optimal choice for highly imbalanced clinical risk prediction, where minimizing false negatives is a priority.
\begin{figure}[!h]
	\centering
	\includegraphics[width=0.8\textwidth]{Fig 5.png}
	\caption{\textbf{Confusion Matrix for Random Forest Model}}
	%\label{fig:fig01}
\end{figure}

For the Support Vector Machine model, Fig 7 presents a confusion matrix with 915 true negatives, 641 true positives, 138 false negatives, and 358 false positives. The substantial number of false positives indicates a challenge with specificity, leading to a higher rate of incorrect high-risk classifications. While the model maintains reasonable sensitivity, its lower precision suggests it may generate a significant number of false alerts, potentially detracting from its clinical utility by increasing unnecessary follow-up investigations.

\begin{figure}[!h]
	\centering
	\includegraphics[width=0.8\textwidth]{Fig 6.png}
	\caption{\textbf{Confusion Matrix for SVM Model}}
	%\label{fig:fig01}
\end{figure}

The confusion matrix for the Decision Tree model, shown in Fig 8, reveals 882 true negatives and 863 true positives, with 97 false negatives and 116 false positives. This performance profile indicates a model with fair discriminative power but notable overfitting vulnerability, as evidenced by appreciable errors in both directions. The relatively higher error rates underscore the limitations of a single tree structure in managing the complexity and nonlinear interactions inherent in cardiovascular risk data.

\begin{figure}[!h]
	\centering
	\includegraphics[width=0.8\textwidth]{Fig 7.png}
	\caption{\textbf{Confusion Matrix for the Decision Tree Model}}
	%\label{fig:fig01}
\end{figure}


As shown in Fig 9, the Keras MLP model's confusion matrix shows 806 true negatives, 785 true positives, 173 false negatives, and 194 false positives. The elevated counts of both types of errors indicate suboptimal learning from the tabular data, likely due to the model's architectural simplicity or insufficient feature representation capacity, even with extensive tuning or larger datasets. This results in decreased reliability for precise clinical stratification.

\begin{figure}[!h]
	\centering
	\includegraphics[width=0.8\textwidth]{Fig 8.png}
	\caption{\textbf{Confusion Matrix for Keras MLP Model}}
	%\label{fig:fig01}
\end{figure}

The Logistic Regression model's performance, depicted in Fig 10, yields 903 true negatives, 476 true positives, 503 false negatives, and 76 false positives. The extremely high number of false negatives highlights the model's critically low sensitivity, failing to identify a majority of actual stroke cases. While specificity remains high, this severe imbalance renders the model clinically inadequate for stroke risk screening, where missing true cases carries significant adverse consequences.

\begin{figure}[!h]
	\centering
	\includegraphics[width=0.8\textwidth]{Fig 9.png}
	\caption{\textbf{Confusion Matrix for Logistic Regression Model}}
	%\label{fig:fig01}
\end{figure}


The confusion matrix for the Scikit-learn MLP classifier, presented in Fig 11, shows a model with moderate, balanced, yet suboptimal predictive performance for stroke risk in patients with coronary heart disease. The model correctly identified 799 true negatives and 818 true positives, indicating reasonable overall discriminative capacity. However, with 180 false positives and 161 false negatives, the classifier exhibits substantial misclassification rates in both directions. The substantial number of false positives suggests reduced specificity, potentially leading to unnecessary clinical investigations and increased healthcare resource utilization. Conversely, the considerable count of false negatives indicates compromised sensitivity, meaning a significant portion of true stroke cases would be overlooked, posing a serious risk in preventive care contexts.

\begin{figure}[!h]
	\centering
	\includegraphics[width=0.8\textwidth]{Fig 10.png}
	\caption{\textbf{Confusion Matrix for Scikit-learn MLP classifier}}
	%\label{fig:fig01}
\end{figure}


Finally, the confusion matrix for the Naive Bayes classifier, presented in Fig 12, shows 770 true negatives and 558 true positives, with 421 false negatives and 209 false positives. This model exhibits the weakest performance among the evaluated algorithms, with high rates of both missed strokes and false alarms. The poor performance underscores the inappropriateness of its conditional independence assumption for the complex, interrelated risk factors present in coronary heart disease and stroke prediction, severely limiting its practical application in this clinical domain.
\begin{figure}[!h]
	\centering
	\includegraphics[width=0.8\textwidth]{Fig 11.png}
	\caption{\textbf{Confusion Matrix for Naive Bayes classifier}}
	%\label{fig:fig01}
\end{figure}


\subsection*{Comparison of Machine Learning Algorithms for predicting Stroke among patients with heart disease}
The machine learning results demonstrate apparent performance differences across the evaluated algorithms, with the stacked ensemble model providing the strongest and most stable predictive ability for Stroke among patients with coronary heart disease, as shown in Table 2. The stacked ensemble achieved the highest overall accuracy and one of the highest precision scores, reflecting its ability to combine the strengths of individual models while reducing their weaknesses. Its high recall and specificity further confirm that it effectively identifies both Stroke and non-stroke cases with minimal error. The Matthews correlation coefficient, one of the most reliable measures for imbalanced clinical datasets, was also the highest for the stacked ensemble model, indicating consistent performance across all cells of the confusion matrix. The very low Brier score shows that the model produced well-calibrated probability estimates, making it suitable for real--world clinical decision-making.

LightGBM also performed exceptionally well and was nearly as good as the stacked ensemble. It achieved the highest precision of all models and maintained very high accuracy, recall, and specificity. The similarity in performance between LightGBM and the stacked ensemble underscores the advantage of gradient-boosting decision trees in structured clinical datasets. These models can capture complex interactions among predictors and remain robust to nonlinear relationships. XGBoost ranked slightly below LightGBM but still demonstrated strong metrics across accuracy, precision, recall, specificity, and F1 score. Its high Matthews correlation coefficient and relatively low Brier score indicate that it remained a reliable and stable model, although not as strong as the top two boosting methods.

Deep learning models showed moderate to strong performance. The Keras CNN model outperformed both the Keras MLP and the scikit learn MLP. It achieved high levels of accuracy, precision, and recall, suggesting that convolutional layers were effective in extracting meaningful representations from tabular data. The multilayer perceptron models had lower predictive scores, likely because they require larger datasets or additional feature engineering to fully capture complex clinical patterns. Their Matthews correlation coefficients and Brier scores were also weaker, indicating reduced stability and probability calibration.

Traditional machine learning models, such as Random Forest and Support Vector Machine, performed reasonably well but did not reach the level of the advanced gradient boosting or ensemble techniques. Random Forest achieved balanced accuracy and recall, with a relatively strong Matthew's correlation coefficient, suggesting it detected stroke cases more reliably than the Support Vector Machine. The Support Vector Machine showed good precision and specificity but had a comparatively lower recall, indicating a tendency to miss a proportion of true stroke cases.

Classical statistical models, including logistic Regression and naive Bayes, had the lowest performance across nearly all metrics. Logistic Regression achieved reasonably high specificity but extremely low recall, indicating that it failed to identify a substantial proportion of stroke cases, leading to a high number of false negatives. The F1 score and Matthew's correlation coefficient were also low, reinforcing its limited ability in this clinical prediction context. Naive Bayes performed similarly, with reduced accuracy, precision, and recall. Its high Brier score suggests poor calibration of predicted probabilities. These results align with established limitations of linear and distribution-dependent models when applied to complex nonlinear relationships common in cardiovascular health data.

Overall, the findings show that ensemble learning and gradient boosting techniques delivered the best predictive performance for stroke detection in patients with coronary heart disease. Their superior accuracy, balanced precision and recall, high Matthew's correlation coefficients, and low Brier scores demonstrate that they are more dependable and clinically meaningful than deep learning, traditional machine learning methods, or classical statistical models. Their strong performance supports their suitability for clinical risk prediction and early intervention strategies aimed at preventing Stroke in high-risk cardiovascular populations.


\begin{table}[!ht]
	\begin{adjustwidth}{-2.25in}{0in} % Comment out/remove adjustwidth environment if table fits in text column.
		\centering
		\caption{
			{\bf Performance of Machine Learning Models for Stroke Prediction.}}
		\begin{tabular}{|l+l|l|l|l|l|l|l|l|}
			\hline
			\textbf{Model} & \textbf{Accuracy} & \textbf{Precision} & \textbf{Recall} & \textbf{Specificity} & \textbf{F1} & \textbf{MCC} & \textbf{Brier Score} \\ \thickhline
			Stacked Ensemble & .9883 & .9979 & .9785 & .9980 & .9881 & .9767 & .0106 \\ \hline
			LightGBM & .9883 & .9990 & .9775 & .9990 & .9881 & .9767 & .0108 \\ \hline
			XGBoost & .9811 & .9958 & .9663 & .9959 & .9808 & .9626 & .0159 \\ \hline
			Keras CNN & .9714 & .9978 & .9448 & .9980 & .9706 & .9441 & .0234 \\ \hline
			Random Forest & .9433 & .9384 & .9489 & .9377 & .9436 & .8867 & .0547 \\ \hline
			Support Vector Machine & .8968 & .9293 & .8590 & .9346 & .8928 & .7959 & .0791 \\ \hline
			SKLearn MLP & .8258 & .8196 & .8355 & .8161 & .8275 & .6518 & .1382 \\ \hline
			Decision Tree & .8912 & .8990 & .8815 & .9009 & .8901 & .7826 & .1088 \\ \hline
			Keras MLP & .8126 & .8194 & .8018 & .8233 & .8105 & .6253 & .1384 \\ \hline
			Logistic Regression & .7043 & .8623 & .4862 & .9224 & .6218 & .4540 & .1969 \\ \hline
			Naive Bayes & .6782 & .7275 & .5700 & .7865 & .6392 & .3652 & .2737 \\ \hline
		\end{tabular}
		\begin{flushleft}
			Table notes: Performance metrics including accuracy, precision, recall, specificity, F1 score, Matthews correlation coefficient (MCC), and Brier score for each machine learning model on stroke prediction.
		\end{flushleft}
		\label{tab:ml_model_performance}
	\end{adjustwidth}
\end{table}

\subsection*{ROC Curve Analysis for Stroke Prediction Models}
The receiver operating characteristic curves presented in Fig 13 show apparent differences in the discriminative ability of the machine learning algorithms used to predict Stroke among patients with coronary heart disease. The curves illustrate how well each model distinguishes between individuals with and without Stroke across all possible decision thresholds. The area under the curve values summarizes this performance, with larger values reflecting better separation between positive and negative cases.

The stacked ensemble and LightGBM models demonstrated the strongest discrimination, each producing an area under the curve value of approximately 0.994. Their curves remain close to the upper-left corner of the plot, indicating that they achieved very high true-positive rates even at low false-positive rates. XGBoost closely followed, with an area under the curve of 0.991, reflecting similarly strong classification performance. The Keras CNN model achieved an AUC of 0.988, and the Random Forest model reached 0.986. These results show that both deep learning and tree-based ensemble methods were highly effective at separating Stroke from non-stroke cases.

Support Vector Machine also performed well, with an area under the curve of 0.955, although its curve did not reach the same level as those of the gradient boosting and ensemble methods. The multilayer perceptron models showed moderate performance. The scikit-learn multilayer perceptron achieved an area under the curve of 0.904, while the Keras multilayer perceptron achieved 0.883. The decision tree model performed slightly lower with an area under the curve value of 0.891. These results show that simpler or less-optimized models had a reduced ability to distinguish between classes compared with advanced ensemble methods.

Logistic Regression and Naive Bayes recorded the lowest discrimination performance with area under the curve values of 0.740 and 0.739, respectively. Their curves remain close to the diagonal reference line shown in Figure 13, indicating limited capability to separate Stroke and non-stroke cases. These patterns highlight the challenges that linear and distribution-based models face when applied to complex clinical data that involve nonlinear interactions.

Overall, the ROC AUC results provide strong evidence that ensemble and gradient boosting techniques deliver superior discrimination compared with deep learning, traditional machine learning, and classical statistical models. Fig 13 clearly demonstrates that the stacked ensemble, LightGBM, and XGBoost models are the most effective approaches for stroke prediction in patients with coronary heart disease, making them the most suitable candidates for integration into clinical decision support systems.

\begin{figure}[!h]
	\centering
	\includegraphics[width=0.9\textwidth]{Fig 12.png}
	\caption{\textbf{AUC--ROC Curves for ML Algorithms}}
	%\label{fig:fig01}
\end{figure}

\subsection*{Precision-Recall Curve Analysis for Stroke Prediction Models}
The precision-recall curves in Fig 14 provide further insight into the performance of the machine learning models in predicting Stroke among patients with coronary heart disease. Unlike the receiver operating characteristic curve, which evaluates performance across balanced conditions, the precision--recall curve emphasizes performance in situations where the positive class is less common. This characteristic makes it a handy tool for clinical datasets in which stroke cases represent a small proportion of the population. The area under the precision-recall curve summarizes this performance, with higher values indicating a stronger ability to identify true stroke cases while minimizing false positives.

The stacked ensemble model achieved the highest area under the precision-recall curve, with a value of 0.996. Its curve remains close to the upper boundary throughout the range of recall values, showing that it consistently maintains high precision even when identifying a large proportion of true stroke cases. LightGBM performed almost identically, with an area under the precision-recall curve of 0.996, further confirming its robust discrimination and reliability under imbalanced conditions. XGBoost achieved an area under the precision-recall curve of 0.995, demonstrating similarly strong performance. These three models show minimal performance loss as recall increases, highlighting the stability of gradient boosting and ensemble strategies.

The Keras CNN model performed very well, with an area under the precision-recall curve of 0.992, confirming the ability of convolutional layers to extract meaningful patterns from tabular clinical data. Random Forest also achieved strong performance with a value of 0.987. Although these values are slightly lower than those of the boosting models, their curves remain well above the baseline in Fig 14, indicating consistent separation of positive and negative cases even when the dataset is imbalanced.

The Support Vector Machine showed moderate performance, with an area under the precision-recall curve of 0.958. Its curve indicates that the model achieves reasonable precision but shows a steeper decline as recall increases. The multilayer perceptron models showed weaker performance, with the scikit-learn model achieving an area under the precision-recall curve of 0.905 and the Keras model achieving 0.886. The decision tree model performed slightly worse, with an area under the precision-recall curve of 0.852. These results reflect the limitations of models that rely solely on shallow tree structures or fully connected layers when applied to complex clinical data.

Logistic Regression and Naive Bayes achieved the lowest performance with values of 0.741 and 0.761, respectively. Their precision-recall curves in Fig 14 approach the baseline, indicating limited ability to correctly identify stroke cases, especially at higher recall levels. These models are less able to maintain precision as they attempt to identify more true stroke cases, thereby reducing their suitability for clinical use in early stroke risk detection.

Overall, the area under the precision-recall curves in Fig 14 confirms that the stacked ensemble, LightGBM, and XGBoost models deliver the strongest performance in identifying stroke cases despite class imbalance. Their curves remain consistently high across the full range of recall values, reinforcing their superiority over deep learning models, traditional machine learning approaches, and classical statistical methods.

\begin{figure}[!h]
	\centering
	\includegraphics[width=0.9\textwidth]{Fig 13.png}
	\caption{\textbf{Precision-Recall Curve for ML Algorithm}}
	%\label{fig:fig01}
\end{figure}

\subsection*{Calibration Curve Analysis for Stroke Prediction Models}
The calibration curves for all evaluated machine learning models provide a critical assessment of the reliability of predicted stroke risk probabilities among patients with coronary heart disease. Calibration curves plot the observed event frequency against the mean predicted probability for binned predictions, with ideal calibration represented by a diagonal line where the predicted probabilities exactly match the observed outcomes. As shown in Fig 15, the Stacked Ensemble and LightGBM models demonstrated near-perfect calibration, with their curves closely aligning with the ideal diagonal. This is supported quantitatively by their exceptionally low Brier scores of 0.011, indicating minimal discrepancy between predicted risks and observed event rates. Such high calibration is clinically essential, as it ensures that a predicted stroke risk of, for example, 20\% corresponds to an actual event frequency of approximately 20\% in similar patient groups, thereby enabling trustworthy individualized risk stratification.

Fig 15 also reveals distinct tiers of calibration performance among the other models. XGBoost and the Keras CNN1D model followed with strong calibration, evidenced by their curves remaining close to the diagonal and their low Brier scores of 0.016 and 0.023, respectively. Models such as Random Forest and Support Vector Machine exhibited moderate calibration, with visible deviations from the ideal line and higher Brier scores of 0.055 and 0.079, suggesting a tendency towards underconfidence or overconfidence in specific probability ranges. In contrast, classical models such as Logistic Regression and Naive Bayes showed poor calibration, with curves deviating substantially from the diagonal and Brier scores of 0.197 and 0.274, respectively. This indicates that their predicted probabilities are not well aligned with actual outcomes, limiting their utility for precise clinical risk estimation. The overall pattern in Fig 15 reinforces that advanced ensemble and gradient boosting methods not only achieve superior discriminative performance but also produce well-calibrated probability estimates, which are crucial for informed clinical decision-making and the implementation of personalized preventive strategies.


\begin{figure}[!h]
	\centering
	\includegraphics[width=0.9\textwidth]{Fig 14.png}
	\caption{\textbf{Calibration Curves for ML Algorithms}}
	%\label{fig:fig01}
\end{figure}

\subsection*{Learning Curve Analysis for Stroke Prediction Models}
The learning curve for the LightGBM model shown in Fig 16 demonstrates excellent generalization capability. Both the training and cross-validation F1 scores rise quickly and converge to a high, stable plateau as the number of training examples increases. The small gap between the training and validation curves indicates that the model is learning the underlying patterns effectively without overfitting. This robust learning behavior supports LightGBM's suitability for deployment on clinical datasets of varying sizes, ensuring reliable performance in real-world settings.

\begin{figure}[!h]
	\centering
	\includegraphics[width=0.9\textwidth]{Fig 15.png}
	\caption{\textbf{Learning Curve for Light GBM Model}}
	%\label{fig:fig01}
\end{figure}

As shown in Fig 17, the XGBoost model exhibits a similarly strong learning trajectory. The training and cross-validation F1 scores improve consistently with more data, converging at a high level of performance. A slight, persistent gap between the curves suggests a minor degree of overfitting, which is well managed by the model's built-in regularization. This pattern confirms XGBoost's ability to leverage additional data to improve its predictive stability and accuracy in stroke risk prediction.

\begin{figure}[!h]
	\centering
	\includegraphics[width=0.9\textwidth]{Fig 16.png}
	\caption{\textbf{Learning Curve for XGBoost Model}}
	%\label{fig:fig01}
\end{figure}


Fig 18 displays the learning curve for the Random Forest model. While both curves achieve high final F1 scores, a consistent and noticeable gap exists between their training and validation performance. This indicates that the model is overfitting to the training data to some extent, a common trait of bagging ensembles that memorize noise. Nevertheless, the cross-validation score remains strong, confirming the model's utility, though it benefits less from additional data compared to boosting methods.

\begin{figure}[!h]
	\centering
	\includegraphics[width=0.9\textwidth]{Fig 17.png}
	\caption{\textbf{Learning Curve for Random Forest Model}}
	%\label{fig:fig01}
\end{figure}

The training history for the Keras CNN1D model shown in Fig 19 shows a smooth, rapid decline in both training and validation loss over epochs. The two curves decrease in close concordance and stabilize at a low level of loss. This ideal convergence indicates that the convolutional architecture learned meaningful feature representations from the tabular data effectively, without signs of overfitting, and that the training process was well-regularized.

\begin{figure}[!h]
	\centering
	\includegraphics[width=0.9\textwidth]{Fig 18.png}
	\caption{\textbf{Training Curves for Keras CNN1D Model}}
	%\label{fig:fig01}
\end{figure}

In contrast, the learning curve for the Decision Tree model, shown in Fig 20, reveals a characteristic pattern of high variance. The training F1 score reaches near-perfect levels very quickly, while the cross-validation F1 score plateaus at a significantly lower value. This large and widening gap is a clear sign of severe overfitting: the single tree structure memorizes the training data but fails to generalize well to unseen cases, limiting its clinical reliability.

\begin{figure}[!h]
	\centering
	\includegraphics[width=0.9\textwidth]{Fig 19.png}
	\caption{\textbf{Learning Curve for Decision Tree Model}}
	%\label{fig:fig01}
\end{figure}


The learning curve for the Support Vector Machine (SVM), shown in Fig 21, indicates moderate final performance with a concerning trend. As the training size increases, the gap between the training and cross-validation F1 scores widens. This suggests that the SVM model is becoming increasingly overfit as more data are added, likely due to the complexity of the kernelized decision boundary in the high-dimensional space of clinical features.
\begin{figure}[!h]
	\centering
	\includegraphics[width=0.9\textwidth]{Fig 20.png}
	\caption{\textbf{Learning Curve for SVM Model}}
	%\label{fig:fig01}
\end{figure}

As shown in Fig 22, the Scikit-learn MLP classifier exhibits high variance in its learning curve. The training F1 score is consistently and significantly higher than the cross-validation score, which plateaus at a moderate level. This substantial gap indicates that the multilayer perceptron is overfitting to the training set and may require more aggressive regularization, architectural adjustments, or substantially more data to improve its generalization.

\begin{figure}[!h]
	\centering
	\includegraphics[width=0.9\textwidth]{Fig 21.png}
	\caption{\textbf{Learning Curve for Scikit-learn MLP Classifier}}
	%\label{fig:fig01}
\end{figure}

The training history for the Keras MLP model shown in Fig 23 indicates that the training loss decreases rapidly. However, the validation loss plateaus after a few epochs and even shows a slight upward trend later in training. This divergence is a classic indicator of overfitting, suggesting that the model would benefit from techniques such as early stopping, dropout, or reduced model complexity to improve its validation performance.

\begin{figure}[!h]
	\centering
	\includegraphics[width=0.9\textwidth]{Fig 22.png}
	\caption{\textbf{Training Curves for Keras MLP model}}
	%\label{fig:fig01}
\end{figure}

Fig 24 illustrates the learning curve for Logistic Regression. Both the training and cross-validation F1 scores remain low and stable across all dataset sizes, with almost no gap between them. This flat, low-performance profile is a definitive sign of underfitting, indicating that the linear model is fundamentally too simple to capture the complex, nonlinear relationships governing stroke risk in this patient population.

\begin{figure}[!h]
	\centering
	\includegraphics[width=0.9\textwidth]{Fig 23.png}
	\caption{\textbf{Learning Curves for Logistic Regression Model}}
	%\label{fig:fig01}
\end{figure}


Finally, the learning curve for the Naive Bayes classifier, shown in Fig 25, confirms its severe limitations. The training and cross-validation F1 scores are low, stable, and virtually identical across the smallest and largest training sets. This complete lack of improvement with additional data, coupled with the model's poor asymptotic performance, underscores that its conditional independence assumption is too restrictive for this complex clinical prediction task, leading to persistent underfitting.

\begin{figure}[!h]
	\centering
	\includegraphics[width=0.9\textwidth]{Fig 24.png}
	\caption{\textbf{ Learning Curve for Naive Bayes Classifier}}
	%\label{fig:fig01}
\end{figure}

\subsection*{Scalability Analysis for Machine Learning Model Deployment}
Scalability was evaluated to determine the feasibility of deploying the trained models in real-world clinical environments, particularly those with constrained computational resources such as primary health facilities, mobile health systems, and edge devices. The assessment focused on four dimensions of computational efficiency: inference latency, training time, memory requirements during model training and inference, and final model size, as shown in Table 3. These metrics collectively indicate whether each model can operate effectively in settings with limited hardware capacity, response time, and storage.

Inference latency refers to the average time required for a model to generate a prediction for a single patient. Low latency is essential for use cases such as point-of-care screening, triage support, and mobile health applications that require real-time responses. The results show that most models have microsecond-level inference times, with the Decision Tree and SKLearn\_MLP emerging as the fastest learners. In contrast, the Keras\_CNN1D and Support Vector Machine models demonstrated higher inference times, reflecting the computational intensity of convolutional and kernel-based operations.

Training time measures how long each model requires to learn from the dataset. Models with shorter training times are advantageous for scenarios requiring frequent retraining, such as incorporating new patient data or adapting to population-specific trends. Logistic Regression and Naive Bayes trained the fastest, completing in under one second, while deep learning models such as Keras\_CNN1D required substantially longer training durations. These differences reflect the greater parameter complexity and optimization demands of neural architectures.

Training memory and inference memory quantify peak RAM usage during model learning and prediction generation, respectively. Naive Bayes and Decision Tree models consumed the least memory, making them suitable for budget-constrained environments, whereas neural networks, particularly Keras\_MLP and Keras\_CNN1D, required significantly more memory during training. Despite this, their inference memory remained modest, indicating that deployment is still feasible on mid-range systems.

Model size reflects storage requirements for saving and loading the trained model. This metric is critical for implementation in devices with limited disk capacity, such as mobile phones or embedded systems. Naive Bayes and Logistic Regression yielded the smallest models, occupying only a few kilobytes. At the same time, Random Forest produced the largest model, exceeding 30 megabytes due to the large number of trees and the storage of split parameters.

Overall, the scalability results demonstrate that lightweight models such as Logistic Regression, Decision Tree, and Naive Bayes are the most efficient for low-resource deployment. However, more complex models such as XGBoost and LightGBM maintain a strong balance between computational efficiency and predictive performance, making them suitable for both resource-limited and high-performance clinical environments. Deep learning models deliver strong predictive performance but require substantial computational resources during training, though their inference demands remain manageable.


\begin{table}[!ht]
	\centering
	\caption{{\bf Scalability Metrics for Machine Learning Models.}}
	\resizebox{\textwidth}{!}{%
		\begin{tabular}{|l|l|l|l|l|l|}
			\hline
			\textbf{Model} &
			\textbf{Latency (s)} &
			\textbf{Train Time (s)} &
			\textbf{Train Mem (MB)} &
			\textbf{Infer Mem (MB)} &
			\textbf{Size (KB)} \\ \hline
			LightGBM & 2.15e-5 & 1.03 & 10.01 & 0.15 & 685.80 \\ \hline
			XGBoost & 5.57e-6 & 1.60 & 9.06 & 0.07 & 400.21 \\ \hline
			Keras CNN1D & 1.77e-4 & 49.15 & 44.47 & 0.63 & 135.77 \\ \hline
			Random Forest & 4.81e-5 & 1.48 & 32.06 & 0.02 & 32392.67 \\ \hline
			SVM & 9.59e-4 & 27.20 & 20.40 & 0.02 & 2357.71 \\ \hline
			SKLearn MLP & 1.22e-6 & 19.07 & 1.10 & 0.23 & 219.46 \\ \hline
			Decision Tree & 5.81e-7 & 0.15 & 0.69 & 0.01 & 98.82 \\ \hline
			Keras MLP & 1.73e-4 & 16.64 & 226.91 & 0.52 & 245.88 \\ \hline
			Logistic Reg. & 1.26e-6 & 0.04 & 3.16 & 0.04 & 1.36 \\ \hline
			Naive Bayes & 1.77e-6 & 0.01 & 0.11 & 1.94 & 2.76 \\ \hline
	\end{tabular}}
	\label{tab:ml_scalability}
\end{table}

\section*{Explainability and Interpretability}

The inclusion of explainable artificial intelligence (XAI) techniques, specifically SHAP (SHapley Additive exPlanations) and LIME (Local Interpretable Model agnostic Explanations), significantly strengthens the interpretability of the machine learning models developed in this study. The following figures provide both global and local insights into model predictions, enabling clinicians to understand and trust the model's decisions.

\subsection*{Global Feature Importance}

Fig~\ref{fig:shap_bar_xgboost} and Fig~\ref{fig:shap_bar_lightgbm} present the mean absolute SHAP values for the XGBoost and LightGBM models, respectively. These bar plots illustrate the average impact of each feature on the model output magnitude. For instance, \texttt{HeartDisease\_1} and \texttt{BMI} are consistently among the top predictors in both models, aligning with clinical knowledge that a history of heart disease and body mass index are critical risk factors for stroke among patients with coronary heart disease. The similarity in feature rankings between the two high-performing models reinforces the robustness of these predictors.

Fig~\ref{fig:shap_stacked_bar} compares the global feature importance across all evaluated models. It highlights that tree-based models (LightGBM, XGBoost, Random Forest) generally assign higher importance to clinical features like \texttt{HeartDisease} and \texttt{BMI} compared to other model types. This visualization is crucial for selecting not only the most accurate model but also the one whose decision-making process aligns best with clinical reasoning.

\subsection*{Global Model Output Distributions}

Fig~\ref{fig:shap_stacked_beeswarm} provides a beeswarm plot that aggregates SHAP values across all samples for each model. The spread and density of points show how each feature influences predictions across the entire dataset. Features such as \texttt{BMI} and \texttt{PhysHlth} show a wide distribution of SHAP values, indicating their complex, non-linear relationship with stroke risk. The plot confirms that the top models (LightGBM, XGBoost) produce more concentrated and interpretable effect patterns compared to models with poorer performance, such as SVM or Decision Tree.

Fig~\ref{fig:shap_beeswarm_lightgbm} offers a detailed view for the LightGBM model. It clearly shows that high values of \texttt{HeartDisease\_0} (no history of heart disease) push the prediction towards a lower stroke risk (negative SHAP values), while high \texttt{BMI} values push predictions towards a higher risk (positive SHAP values). This level of detail is invaluable for understanding the direction and magnitude of each feature's effect.

\subsection*{Local Explanations for Individual Predictions}

The local explanation plots are essential for interpreting individual case predictions, a key requirement for clinical deployment.

Figs~\ref{fig:shap_local_xgboost_neg} and \ref{fig:shap_local_lightgbm_neg} demonstrate SHAP force plots for negative predictions (low stroke risk) from the XGBoost and LightGBM models. They decompose how each feature contributes to pulling the base value \(E[f(X)]\) down to the final prediction \(f(x)\). For example, in Fig~\ref{fig:shap_local_xgboost_neg}, features like \texttt{HighChol\_0} and \texttt{Veggies\_0} decrease the risk score. Conversely, a high \texttt{BMI} increases it, but the net effect remains negative.

Figs~\ref{fig:shap_local_xgboost_pos} and \ref{fig:shap_local_lightgbm_pos} show force plots for positive predictions (high stroke risk). Here, features such as \texttt{HeartDisease\_1}, \texttt{Age\_10}, and \texttt{Education\_6} are the primary drivers increasing the risk score above the base value. The clear additive nature of SHAP values makes it easy to audit why a specific patient was flagged as high-risk.

Complementing the SHAP explanations, Figs~\ref{fig:lime_lightgbm_neg} and \ref{fig:lime_lightgbm_pos} present LIME explanations for LightGBM predictions. LIME creates a locally faithful interpretable model (e.g., linear) around a single prediction. For the negative case (Fig~\ref{fig:lime_lightgbm_neg}), conditions like \texttt{Age\_8 > 0.00} and \texttt{Diabetes\_2 > 0.00} are shown to support the low-risk classification. For the positive case (Fig~\ref{fig:lime_lightgbm_pos}), conditions like \texttt{Age\_10 > 0.00} and \texttt{Income\_4 > 0.00} justify the high-risk prediction. The use of both SHAP and LIME provides a multi-faceted view: SHAP offers a consistent, theoretically grounded attribution, while LIME provides a simple, rule-based explanation that may be more intuitive for some end-users.

\subsection*{Synthesis and Clinical Utility}

Collectively, these visualizations move the model from a "black box" to a transparent and auditable tool. The global plots (Figs~\ref{fig:shap_bar_xgboost}, \ref{fig:shap_bar_lightgbm}, \ref{fig:shap_stacked_bar}, \ref{fig:shap_stacked_beeswarm}) validate the model at a population level, ensuring that the most influential features are clinically plausible. The local plots (Figs~ \ref{fig:shap_local_xgboost_neg}--\ref{fig:lime_lightgbm_pos}) empower clinicians at the point of care. They can answer the critical question: "Why did the model predict a high stroke risk for *this* particular patient?"

To fully leverage this in your manuscript, ensure you reference these figures when discussing model interpretability in the Results and Discussion sections. For example:
\begin{quote}
	"The superior performance of the LightGBM model was complemented by its interpretability. As shown in Fig~\ref{fig:shap_beeswarm_lightgbm}, BMI and history of heart disease were the most impactful features globally. Furthermore, local explanations for individual high-risk predictions (Fig~\ref{fig:shap_local_lightgbm_pos}) consistently highlighted advanced age and low vegetable intake as contributing factors, providing actionable insights for targeted patient counseling."
\end{quote}

% ==================== EXPLAINABILITY FIGURES ====================

% Global SHAP Bar Plots
\begin{figure}[!h]
	\centering
	\includegraphics[width=0.9\textwidth]{shap_bar_XGBoost.png}
	\caption{Global feature importance (mean absolute SHAP value) for the XGBoost model.}
	\label{fig:shap_bar_xgboost}
\end{figure}

\begin{figure}[!h]
	\centering
	\includegraphics[width=0.9\textwidth]{shap_bar_LightGBM.png}
	\caption{Global feature importance (mean absolute SHAP value) for the LightGBM model.}
	\label{fig:shap_bar_lightgbm}
\end{figure}

\begin{figure}[!h]
	\centering
	\includegraphics[width=0.9\textwidth]{shap_stacked_bar.png}
	\caption{Comparison of global feature importance across all evaluated models.}
	\label{fig:shap_stacked_bar}
\end{figure}

% SHAP Beeswarm Plots
\begin{figure}[!h]
	\centering
	\includegraphics[width=0.9\textwidth]{shap_stacked_beeswarm.png}
	\caption{SHAP beeswarm plot showing feature effects across all models. Each point represents a patient, colored by feature value.}
	\label{fig:shap_stacked_beeswarm}
\end{figure}

\begin{figure}[!h]
	\centering
	\includegraphics[width=0.9\textwidth]{shap_beeswarm_LightGBM.png}
	\caption{SHAP beeswarm plot for the LightGBM model. The plot illustrates how each feature contributes to model predictions across the dataset.}
	\label{fig:shap_beeswarm_lightgbm}
\end{figure}

% Local SHAP Explanations
\begin{figure}[!h]
	\centering
	\includegraphics[width=0.9\textwidth]{shap_local_XGBoost_negative.png}
	\caption{Local SHAP explanation for a low-risk prediction from the XGBoost model. The plot shows how individual features push the prediction away from the base value.}
	\label{fig:shap_local_xgboost_neg}
\end{figure}

\begin{figure}[!h]
	\centering
	\includegraphics[width=0.9\textwidth]{shap_local_LightGBM_negative.png}
	\caption{Local SHAP explanation for a low-risk prediction from the LightGBM model.}
	\label{fig:shap_local_lightgbm_neg}
\end{figure}

\begin{figure}[!h]
	\centering
	\includegraphics[width=0.9\textwidth]{shap_local_XGBoost_positive.png}
	\caption{Local SHAP explanation for a high-risk prediction from the XGBoost model.}
	\label{fig:shap_local_xgboost_pos}
\end{figure}

\begin{figure}[!h]
	\centering
	\includegraphics[width=0.9\textwidth]{shap_local_LightGBM_positive.png}
	\caption{Local SHAP explanation for a high-risk prediction from the LightGBM model.}
	\label{fig:shap_local_lightgbm_pos}
\end{figure}

% LIME Explanations
\begin{figure}[!h]
	\centering
	\includegraphics[width=0.9\textwidth]{lime_LightGBM_negative.png}
	\caption{LIME explanation for a low-risk prediction from the LightGBM model. The plot shows the most influential features and their contribution to the prediction.}
	\label{fig:lime_lightgbm_neg}
\end{figure}

\begin{figure}[!h]
	\centering
	\includegraphics[width=0.9\textwidth]{lime_LightGBM_positive.png}
	\caption{LIME explanation for a high-risk prediction from the LightGBM model.}
	\label{fig:lime_lightgbm_pos}
\end{figure}

\subsection*{Discussion}
The current research examined how various machine learning models can be used to predict stroke risk in patients with coronary heart disease and yielded meaningful results that align with the general literature on the topic while introducing new insights. Overall, the findings indicate that the best predictive performance was obtained with high-level ensemble learning techniques and gradient boosting algorithms, compared to deep learning, classic machine learning, and classical statistical techniques. This result aligns with the growing body of cardiovascular and cerebrovascular literature indicating that ensemble techniques are more successful at identifying complex trends in structured clinical data than individual models \cite{Moulaei2024, Srinivasu2024, Heo2022, Lin2024}.

The robust performance of stacked ensemble, LightGBM, and XGBoost models in this study supports earlier findings that such approaches can outperform individual ML and deep learning models in stroke and cardiac risk prediction \cite{Si2025, Hassan2024, Vu2024}. The effectiveness of gradient boosting, in particular, underscores the ability of tree-based ensembles to model nonlinear interactions, which are essential for accurately capturing cardiovascular risk profiles. These findings complement prior studies demonstrating that ensemble-based structures yield superior discrimination and calibration relative to single models \cite{Moulaei2024, Srinivasu2024, Abujaber2024}.

Deep learning models showed moderate to high predictive value, with convolutional neural networks outperforming fully connected multilayer perceptrons, consistent with prior work showing that convolutional models can learn meaningful representations from tabular clinical data but may require very large datasets to surpass ensemble methods \cite{Moulaei2024, Wu2024}. Similarly, conventional ML algorithms such as support vector machines and random forests delivered reasonable performance but were generally less discriminative than gradient-boosted ensembles, echoing earlier observations \cite{Gkantzios2023, Dritsas2024, Truong2024}. Decision trees exhibited clear signs of overfitting, reflecting the known limitations of single-tree models in complex medical prediction tasks.

Classical statistical models, including logistic regression and naive Bayes, performed the worst, consistent with research indicating that linear or distribution-dependent models often underfit high-interaction and mixed-type features in clinical data \cite{Mridha2023, Li2025, Shen2025}. The low recall rates of logistic regression in predicting stroke cases among coronary heart disease patients highlight the need for more flexible modelling approaches, as reinforced by studies incorporating ML-based prognostication \cite{Abujaber2024}.

Scalability analysis further demonstrates that ensemble models like LightGBM and XGBoost are not only highly accurate but also computationally efficient, making them suitable for integration into clinical decision support systems, including in resource-limited settings. Deep learning models, while more memory-intensive and slower to train, offer reasonable inference efficiency, suggesting potential utility in cloud-hosted clinical applications \cite{Hwang2025, Mahin2025, Ahmed2024}.

Explainability analyses revealed that SHAP-based global explanations consistently highlighted age, prior cardiovascular events, BMI, and general health indicators as dominant contributors to stroke risk, corroborating prior studies \cite{Moulaei2024, Dritsas2024, Si2025, Gao2024}. The observed consistency of feature rankings across LightGBM and XGBoost aligns with previous reports of stable and interpretable attribution patterns in boosting models \cite{Srinivasu2024, Vu2024}. SHAP beeswarm plots further illustrated nonlinear and interaction-driven effects for features such as BMI and physical health indicators, reflecting the complex relationships identified in other XAI-driven stroke risk studies \cite{Gkantzios2023, Tasnim2025}.

At the individual patient level, SHAP and LIME local explanations provided actionable insights consistent with previous literature \cite{Mridha2023, Li2025, ElSofany2024, Tasnim2025}. High-risk predictions were predominantly driven by advanced age, prior heart disease, and poor general health, while protective lifestyle factors mitigated risk in lower-risk cases. The complementary use of SHAP and LIME adheres to best practices in explainable AI, providing both model-faithful and intuitive rule-based explanations \cite{Hwang2025, Mahin2025, Ahmed2024}.

The integration of traditional logistic regression with XAI methods enabled a more nuanced understanding of stroke risk factors. Logistic regression identified statistically significant predictors, such as prior heart attack, while SHAP and LIME revealed non-linear relationships and interactions that logistic regression could not capture. For instance, BMI, which appeared protective in logistic regression, showed heterogeneous effects in tree-based models, demonstrating increased risk for specific patient subgroups. These findings underscore the value of combining classical statistical models with explainable ML to inform precision medicine.

Finally, this work complements existing studies on stroke risk prediction by addressing mortality and prognostic outcomes in stroke patients \cite{Abujaber2024, Wu2024, Truong2024}, providing a more comprehensive perspective on patient trajectories beyond risk classification. Incorporating explainable hybrid models allows for both accurate prediction and interpretability, bridging the gap between computational sophistication and clinical applicability.

Despite these contributions, several limitations remain. Data were derived from self-reported surveys, introducing recall bias and misclassification for behavioral and lifestyle factors. The cross-sectional design precludes causal inference. High-performing models require external validation using clinical registry or EHR data to confirm generalizability, and the absence of temporal clinical information limits the ability to capture dynamic risk changes. Future work should consider integrating longitudinal and multimodal data to further enhance predictive performance and clinical relevance.


\section*{Conclusion}

This study successfully designed and tested a comprehensive suite of machine learning models to predict stroke risk in patients with coronary heart disease. Our results demonstrate that contemporary ensemble-based methods, particularly stacked ensembles, LightGBM, and XGBoost, deliver outstanding predictive performance, strong generalization capability, and desirable computational efficiency. These models consistently outperformed deep learning architectures, traditional machine learning approaches, and classical statistical methods, proving particularly effective for clinical risk stratification across diverse patient populations. The superiority of ensemble-based models stems from their ability to capture complex, non-linear interactions among behavioral, demographic, and clinical variables that underlie the intricate relationship between coronary and cerebrovascular diseases.

A key contribution of this work is the integration of explainable artificial intelligence (XAI) techniques—specifically SHAP (SHapley Additive exPlanations) and LIME (Local Interpretable Model agnostic Explanations)—which provided transparent and interpretable insights into model predictions. By elucidating both global feature importance and local, case-specific reasoning, these methods address the critical "black box" concern that often impedes clinical adoption of sophisticated machine learning models. The convergence of findings between traditional logistic regression and modern XAI techniques strengthens confidence in identified risk factors, while the additional nuanced relationships revealed by SHAP and LIME enable more personalized risk assessment.

The findings carry significant implications for clinical practice. Given that stroke remains a leading cause of global mortality and long-term disability, the timely identification of high-risk patients with coronary heart disease offers substantial opportunities for targeted prevention. The combination of high accuracy, low inference latency, and inherent interpretability in our best-performing models makes them suitable for integration into real-time clinical decision support systems in both primary care and specialty cardiology settings, as well as population-level screening initiatives. These models thus provide a viable pathway for developing more accurate and individualized prevention programs aligned with global health priorities focused on early diagnosis and personalized management of cardiovascular and neurological diseases.

Future research should focus on further refinement and external validation of these predictive tools across diverse clinical environments. While our models demonstrated strong performance, additional validation using multi-center cohorts, clinical registries, and longitudinal datasets is necessary to enhance generalizability and ensure equitable performance across patient subgroups. Expanding the feature space to include laboratory results, imaging data, and longitudinal clinical markers could further improve model performance and provide deeper insights into the dynamic interplay between coronary disease and stroke risk over time.

\section*{Acknowledgement}
The authors acknowledge the use of artificial intelligence-based tools, specifically Grammarly, for grammar and language refinement during manuscript preparation. Use of this tool was limited to enhancing readability and linguistic clarity; all intellectual content, analysis, interpretations, and conclusions presented in this paper are solely those of the authors.

\section*{Data Availability Statement}
The dataset used in this study has been deposited in Zenodo and is openly available under a CC-BY 4.0 license at the following DOI:https://doi.org/10.5281/zenodo.17897855.
All data are fully accessible without restriction.
\nolinenumbers


\bibliographystyle{plos2015}
\bibliography{references}

\end{document}
