## Supplementary material for "Machine Learning Models for Predicting Stroke Risk Among Patients with Coronary Heart Disease": Latex Files: references.bib

@article{Lolak2023,
  author = {Lolak, S. and Attia, J. and McKay, G. J. and Thakkinstian, A.},
  title = {Comparing explainable machine learning approaches with traditional statistical methods for evaluating stroke risk models: retrospective cohort study},
  journal = {JMIR Cardio},
  year = {2023},
  volume = {7},
  pages = {e47736}
}

@article{Moulaei2024,
  author = {Moulaei, K. and Afshari, L. and Moulaei, R. and Sabet, B. and Mousavi, S. M. and Afrash, M. R.},
  title = {Explainable artificial intelligence for stroke prediction through comparison of deep learning and machine learning models},
  journal = {Scientific Reports},
  year = {2024},
  volume = {14},
  number = {1},
  pages = {31392}
}

@article{Srinivasu2024,
  author = {Srinivasu, P. N. and Sirisha, U. and Sandeep, K. and Praveen, S. P. and Maguluri, L. P. and Bikku, T.},
  title = {An interpretable approach with explainable AI for heart stroke prediction},
  journal = {Diagnostics},
  year = {2024},
  volume = {14},
  number = {2},
  pages = {128}
}

@article{Heo2022,
  author = {Heo, J. and Yoo, J. and Lee, H. and Lee, I. H. and Kim, J. S. and Park, E. and Nam, H. S.},
  title = {Prediction of hidden coronary artery disease using machine learning in patients with acute ischemic stroke},
  journal = {Neurology},
  year = {2022},
  volume = {99},
  pages = {e55--e65}
}

@article{Lin2024,
  author = {Lin, L. and Ding, L. and Fu, Z. and Zhang, L.},
  title = {Machine learning-based models for prediction of the risk of stroke in coronary artery disease patients receiving coronary revascularization},
  journal = {PLOS ONE},
  year = {2024},
  volume = {19},
  pages = {e0296402}
}

@article{Si2025,
  author = {Si, Y. and Abdollahi, A. and Ashrafi, N. and Placencia, G. and Pishgar, E. and Alaei, K. and Pishgar, M.},
  title = {Optimized feature selection and advanced machine learning for stroke risk prediction in revascularized coronary artery disease patients},
  journal = {BMC Medical Informatics and Decision Making},
  year = {2025},
  volume = {25},
  number = {1},
  pages = {276}
}

@article{ElSofany2024,
  author = {El-Sofany, H. and Bouallegue, B. and El-Latif, Y. M. A.},
  title = {A proposed technique for predicting heart disease using machine learning algorithms and an explainable AI method},
  journal = {Scientific Reports},
  year = {2024},
  volume = {14},
  number = {1},
  pages = {23277}
}


@article{Hassan2024,
  author = {Hassan, A. and Gulzar Ahmad, S. and Ullah Munir, E. and Ali Khan, I. and Ramzan, N.},
  title = {Predictive modelling and identification of key risk factors for stroke using machine learning},
  journal = {Scientific Reports},
  year = {2024},
  volume = {14},
  pages = {11498}
}

@article{Vu2024,
  author = {Vu, T. and Kokubo, Y. and Inoue, M. and Yamamoto, M. and Mohsen, A. and Martin-Morales, A. and Araki, M.},
  title = {Machine learning approaches for stroke risk prediction: findings from the Suita Study},
  journal = {Journal of Cardiovascular Development and Disease},
  year = {2024},
  volume = {11},
  pages = {207}
}

@article{Dritsas2024,
  author = {Dritsas, E. and Trigka, M.},
  title = {Application of Deep Learning for Heart Attack Prediction with Explainable Artificial Intelligence},
  journal = {Computers},
  year = {2024},
  volume = {13},
  number = {10},
  pages = {244}
}

@inproceedings{Bhanusri2024,
  author = {Bhanusri, K. and Reddy, K. N. and Gnaneswarachary, P. and Nair, P. C. and Sampath, N.},
  title = {Heart Stroke Prediction Using Machine Learning and Explainable AI Insights},
  booktitle = {2024 IEEE International Conference on Information Technology, Electronics and Intelligent Communication Systems (ICITEICS)},
  year = {2024},
  pages = {1--7},
  publisher = {IEEE}
}

@article{Wang2021,
  author = {Wang, K. and Tian, J. and Zheng, C. and Yang, H. and Ren, J. and Liu, Y. and Zhang, Y.},
  title = {Interpretable prediction of 3-year all-cause mortality in patients with heart failure caused by coronary heart disease based on machine learning and SHAP},
  journal = {Computers in Biology and Medicine},
  year = {2021},
  volume = {137},
  pages = {104813}
}
@article{Truong2024,
  author = {Truong, B. and Zheng, J. and Hornsby, L. and Fox, B. and Chou, C. and Qian, J.},
  title = {Development and validation of machine learning algorithms to predict 1-Year ischemic stroke and bleeding events in patients with atrial fibrillation and Cancer},
  journal = {Cardiovascular Toxicology},
  year = {2024},
  volume = {24},
  pages = {365--374}
}
@article{Gao2024,
  author = {Gao, J. and Mar, P. and Tang, Z. Z. and Chen, G.},
  title = {Fair prediction of 2-year stroke risk in patients with atrial fibrillation},
  journal = {Journal of the American Medical Informatics Association},
  year = {2024},
  volume = {31},
  pages = {2820--2828}
}
@article{Shen2025,
  author = {Shen, D. and Li, J. and Teng, S. and Li, M. and Tang, X.},
  title = {Development and validation of a nomogram for predicting frailty risk among older patients with ischaemic stroke},
  journal = {Journal of Clinical Nursing},
  year = {2025},
  volume = {34},
  pages = {4186--4201}
}
@article{Mridha2023,
  author = {Mridha, K. and Ghimire, S. and Shin, J. and Aran, A. and Uddin, M. M. and Mridha, M. F.},
  title = {Automated stroke prediction using machine learning: an explainable and exploratory study with a web application for early intervention},
  journal = {IEEE Access},
  year = {2023},
  volume = {11},
  pages = {52288--52308}
}

@article{Li2025,
  author = {Li, Q. and Wang, X. and Ye, Q. and Li, L. and He, X. and Liu, L.},
  title = {A prediction model for stroke risk based on explainable machine learning},
  journal = {Medicine},
  year = {2025},
  volume = {104},
  number = {40},
  pages = {e44718}
}

@article{Pamungkas2022,
  author = {Pamungkas, Y. and Wibawa, A. D. and Cahya, M. D.},
  title = {Electronic medical record data analysis and prediction of stroke disease using Explainable Artificial Intelligence (XAI)},
  journal = {Kinetik: Game Technology, Information System, Computer Network, Computing, Electronics, and Control},
  year = {2022}
}

@article{Gkantzios2023,
  author = {Gkantzios, A. and Kokkotis, C. and Tsiptsios, D. and Moustakidis, S. and Gkartzonika, E. and Avramidis, T. and Vadikolias, K.},
  title = {From admission to discharge: predicting national institutes of health stroke scale progression in stroke patients using biomarkers and explainable machine learning},
  journal = {Journal of Personalized Medicine},
  year = {2023},
  volume = {13},
  number = {9},
  pages = {1375}
}

@article{Talaat2024,
  author = {Talaat, F. M. and Elnaggar, A. R. and Shaban, W. M. and Shehata, M. and Elhosseini, M.},
  title = {CardioRiskNet: A hybrid AI-based model for explainable risk prediction and prognosis in cardiovascular disease},
  journal = {Bioengineering},
  year = {2024},
  volume = {11},
  number = {8},
  pages = {822}
}

@article{Kokkotis2022,
  author = {Kokkotis, C. and Giarmatzis, G. and Giannakou, E. and Moustakidis, S. and Tsatalas, T. and Tsiptsios, D. and Aggelousis, N.},
  title = {An explainable machine learning pipeline for stroke prediction on imbalanced data},
  journal = {Diagnostics},
  year = {2022},
  volume = {12},
  number = {10},
  pages = {2392}
}


@inproceedings{Sethi,
  author = {Sethi, A. and Dharmavaram, S. and Somasundaram, S.K.},
  title = {Explainable artificial intelligence (XAI) approach to heart disease prediction},
  booktitle = {3rd International Conference on Artificial Intelligence for Internet of Things},
  year = {2024},
  pages = {1--6}
}

@article{LiJ2025,
  author = {Li, J. and Wu, S. and Gu, J.},
  title = {Explainable machine learning model for assessing health status in patients with comorbid coronary heart disease and depression: Development and validation study},
  journal = {International Journal of Medical Informatics},
  year = {2025},
  volume = {196},
  pages = {105808}
}

@article{Uddin2023,
  author = {Uddin, M. J. and Kumar Sarker, P. and Hoque, N. and Aktar, S. and Ahmad, M. M.},
  title = {An Explainable machine learning framework for early detection of Stroke},
  journal = {International Multilingual Journal of Science and Technology},
  year = {2023},
  volume = {8}
}

@article{Hwang2025,
  author = {Hwang, Y. S. and Kim, S. and Yim, I. and Park, Y. and Kang, S. and Jo, H. S.},
  title = {Predicting the likelihood of readmission in patients with ischemic Stroke: An explainable machine learning approach using common data model data},
  journal = {International Journal of Medical Informatics},
  year = {2025},
  volume = {195},
  pages = {105754}
}
@article{Tasnim2025,
  author = {Tasnim, A. F. and Rahman, R. and Prabha, M. and Hossain, M. A. and Nilima, S. I. and Al Mahmud, M. A. and Erdei, T. I.},
  title = {Explainable machine learning algorithms to predict cardiovascular strokes},
  journal = {Engineering, Technology \& Applied Science Research},
  year = {2025},
  volume = {15},
  pages = {20131--20137}
}
@article{Wu2024,
  author = {Wu, C. and Zhu, S. and Wang, Q. and Xu, Y. and Mo, X. and Xu, W. and Xu, Z.},
  title = {Development, validation, and visualization of a novel nomogram to predict depression risk in patients with stroke},
  journal = {Journal of Affective Disorders},
  year = {2024},
  volume = {365},
  pages = {351--358}
}

@article{Ahmed2024,
  author = {Ahmed, R. and Varshney, A. and Ashraf, Z. and Farooqui, N. A. and Pathak, R. S.},
  title = {Enhanced stroke risk prediction: a fusion of machine learning models for improved healthcare strategies},
  journal = {SN Computer Science},
  year = {2024},
  volume = {5},
  pages = {1078}
}
@article{Veledar2025,
  author = {Veledar, E. and Zhou, L. and Veledar, O. and Gardener, H. and Gutierrez, C. M. and Brown, S. C. and Rundek, T.},
  title = {Identifying determinants of readmission and death post-stroke using explainable machine learning},
  journal = {PloS One},
  year = {2025},
  volume = {20},
  number = {9},
  pages = {e0332371}
}

@article{Liu2025,
  author = {Liu, J. and Gu, Y. and Wang, D. and Xia, X. and Li, X.},
  title = {Prediction of the short-term prognosis of acute ischaemic Stroke in patients with high treatment platelet reactivity using explainable machine learning},
  journal = {Scientific Reports},
  year = {2025},
  volume = {15},
  number = {1},
  pages = {36696}
}

@article{Zhu2025,
  author = {Zhu, H. and Ye, Q. and Wang, S. and Cai, H. and Maimaiti, M. and Lai, J. and Zeng, H.},
  title = {An Explainable Two-Stage Machine Learning Model for Predicting the Post-Thrombolysis Complications in Stroke Patients: A Multi-Center Study},
  journal = {Research},
  year = {2025},
  volume = {8},
  pages = {0817}
}
@article{Abujaber2024,
  author = {Abujaber, A. A. and Albalkhi, I. and Imam, Y. and Nashwan, A. and Akhtar, N. and Alkhawaldeh, I. M.},
  title = {Machine learning-based prognostication of mortality in stroke patients},
  journal = {Heliyon},
  year = {2024},
  volume = {10},
  pages = {7}
}
@article{Mahin2025,
  author = {Mahin, M. A. and Islam, M. M. and Alam, M. Z. and Pollob, A. D. and Zaman, O.},
  title = {Enhancing Stroke Risk Prediction with Explainable AI: Leveraging Resampling and Machine Learning for Improved Accuracy},
  journal = {Journal of Neurointervention and Stroke},
  year = {2025},
  volume = {1},
  number = {2},
  pages = {63--71}
}
@misc{Zheng,
  author = {Zheng, K.},
  title = {Heart disease dataset},
  year = {2025},
  howpublished = {IEEE Dataport},
  url = {https://dx.doi.org/10.21227/49m4-zh81}
}

@inproceedings{Ribeiro2016,
  author    = {Ribeiro, Marco Tulio and Singh, Sameer and Guestrin, Carlos},
  title     = {Why Should I Trust You? Explaining the Predictions of Any Classifier},
  booktitle = {Proceedings of the 22nd ACM SIGKDD International Conference on Knowledge Discovery and Data Mining},
  year      = {2016},
  pages     = {1135--1144},
  publisher = {ACM},
  doi       = {10.1145/2939672.2939778}
}
