## Supplementary figures and images for "Machine Learning Models for Predicting Stroke Risk Among Patients with Coronary Heart Disease"

### Fig 1.png

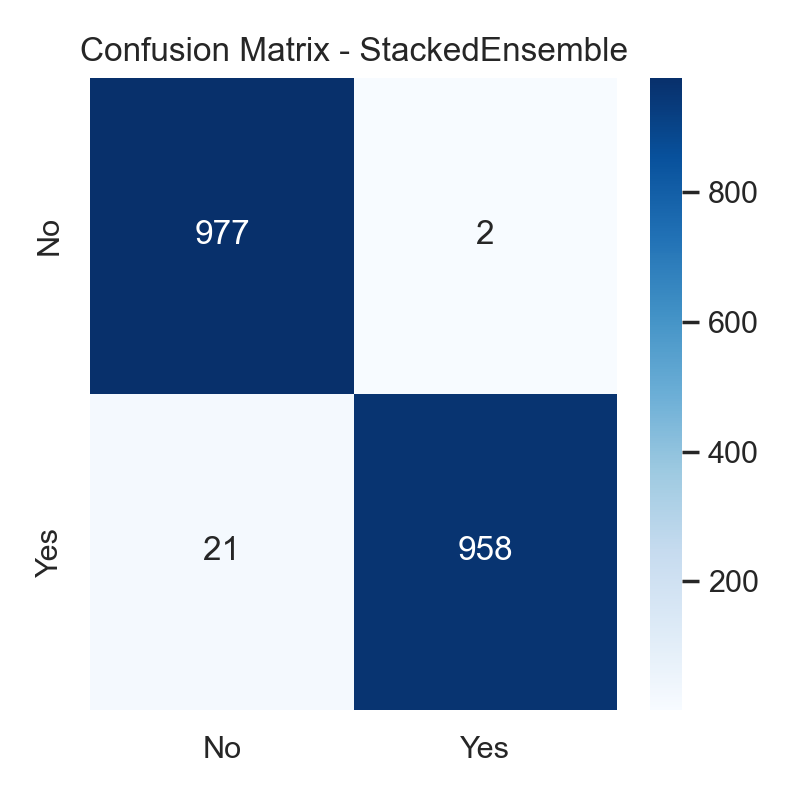

### Fig 2.png

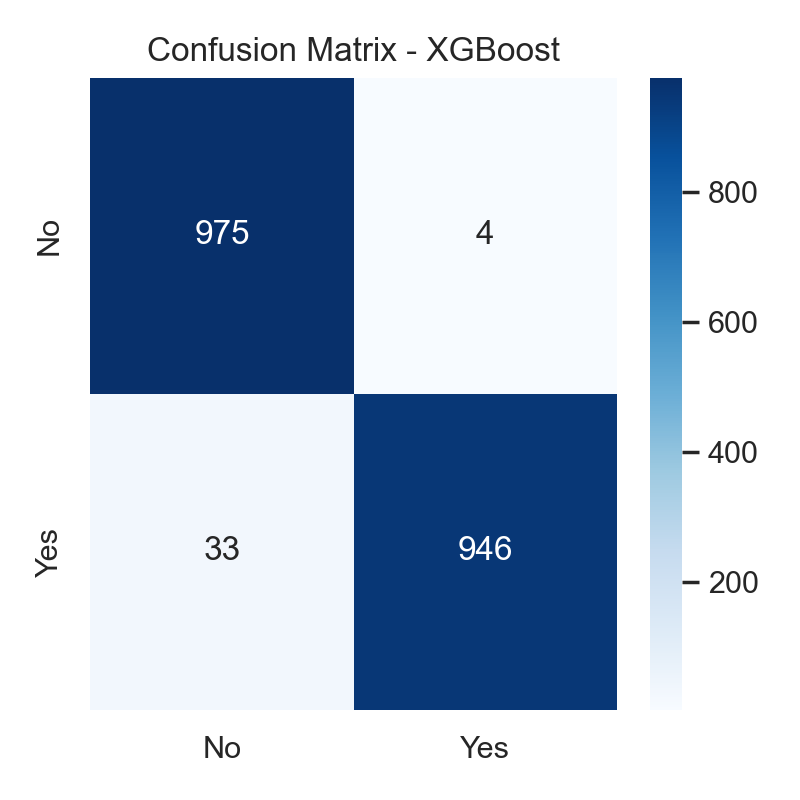

### Fig 3.png

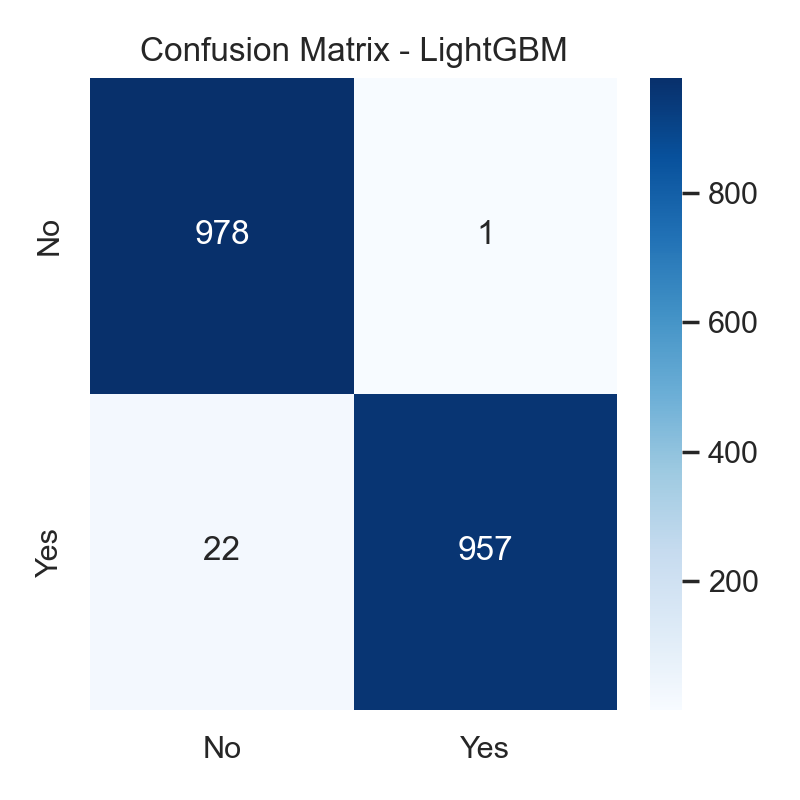

### Fig 4.png

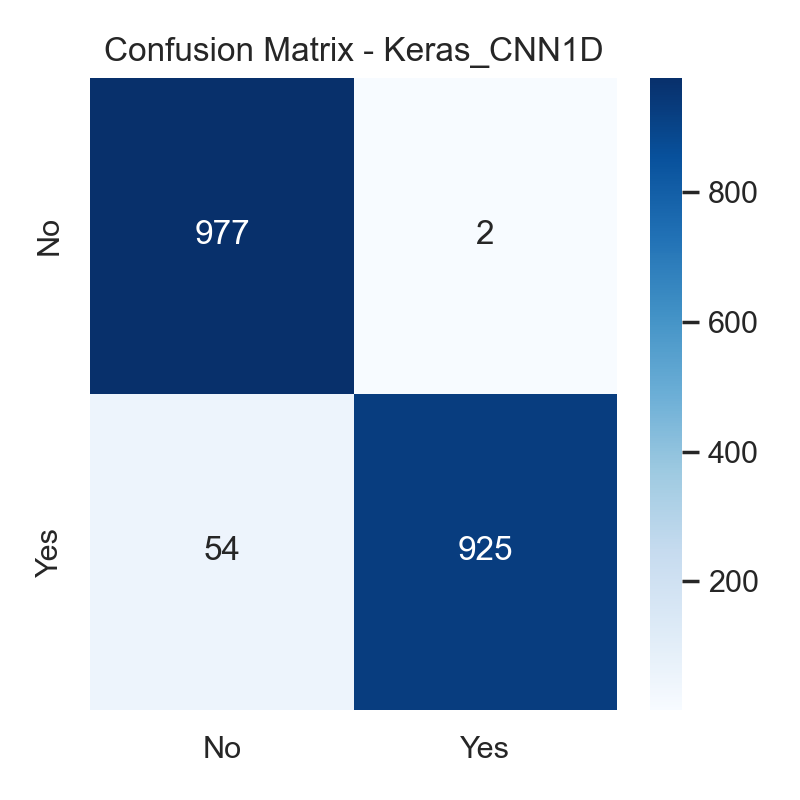

### Fig 5.png

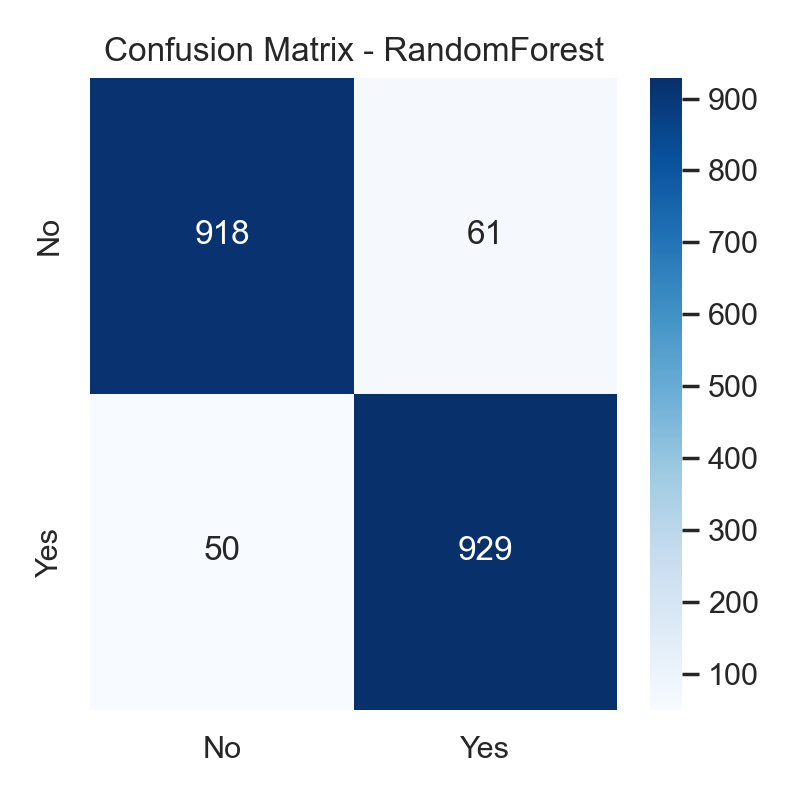

### Fig 6.png

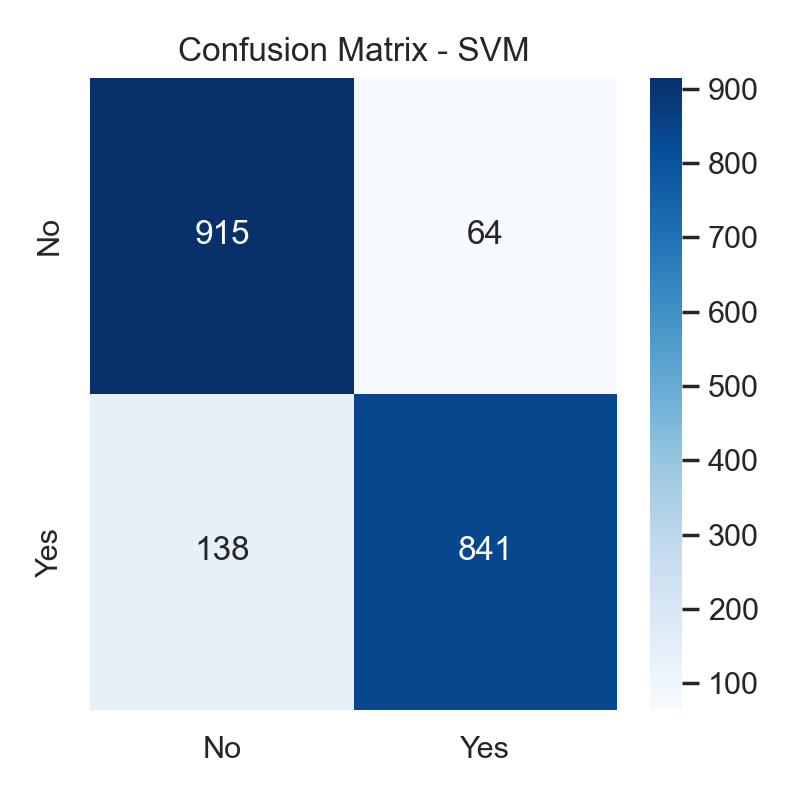

### Fig 7.png

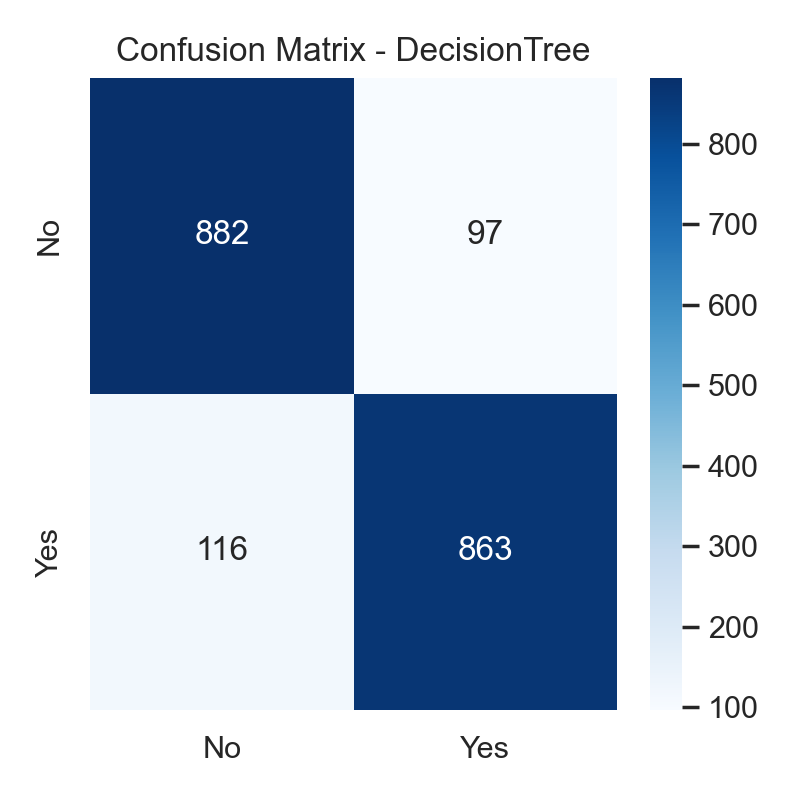

### Fig 8.png

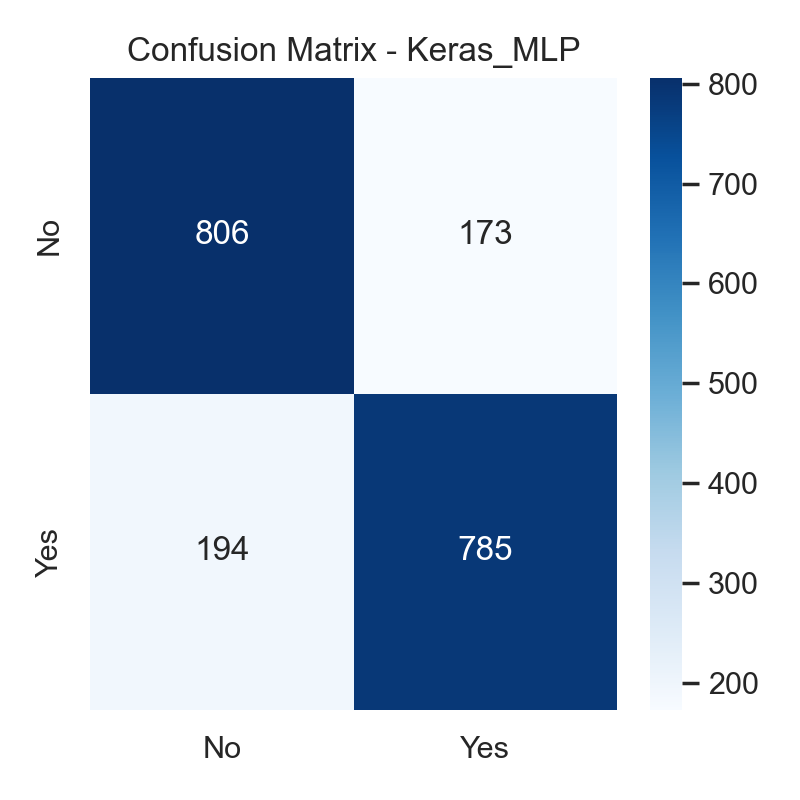

### Fig 9.png

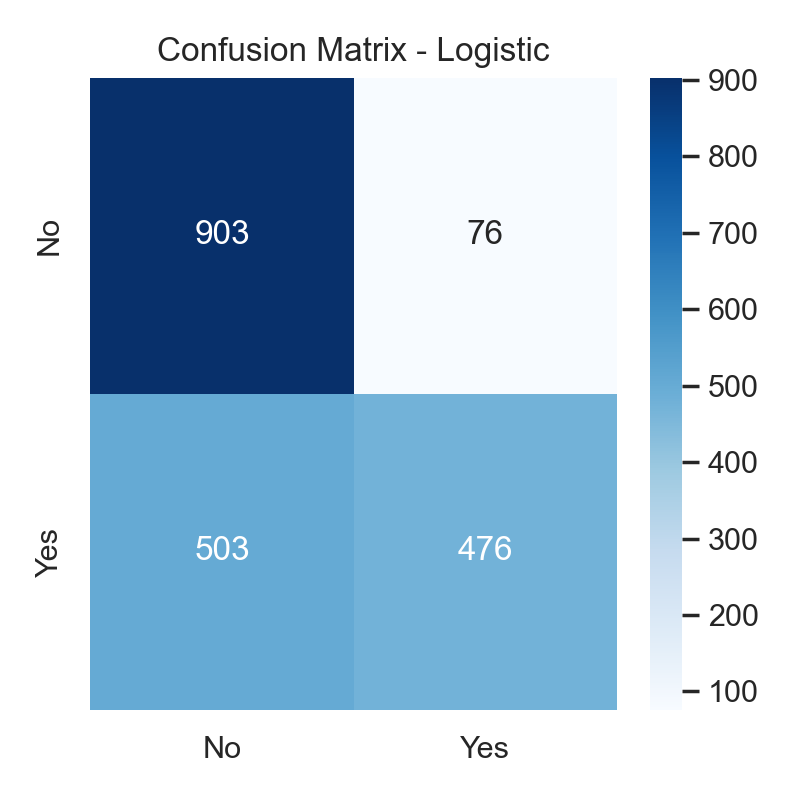

### Fig 10.png

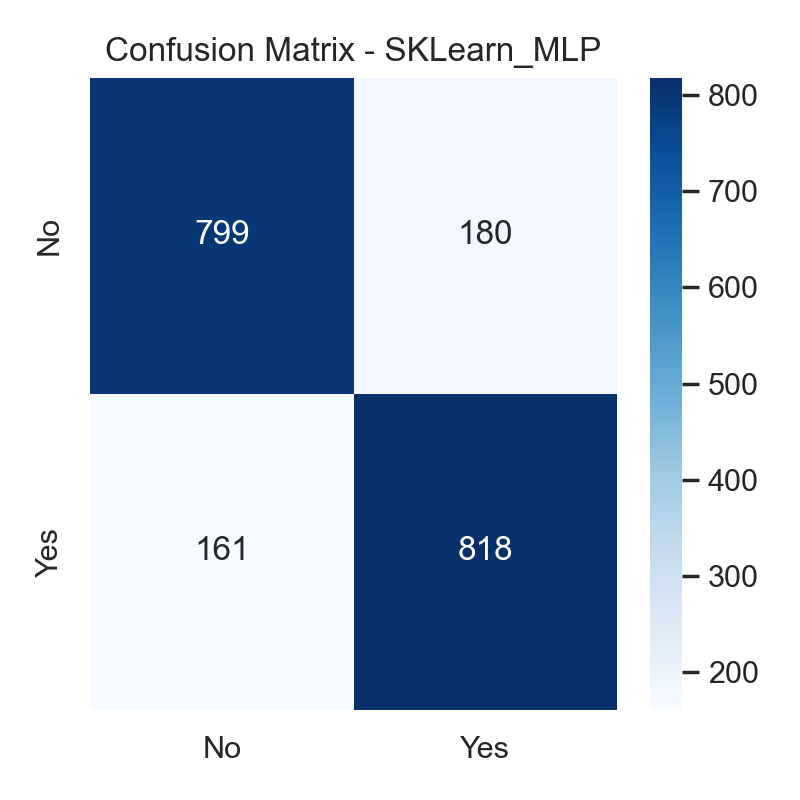

### Fig 11.png

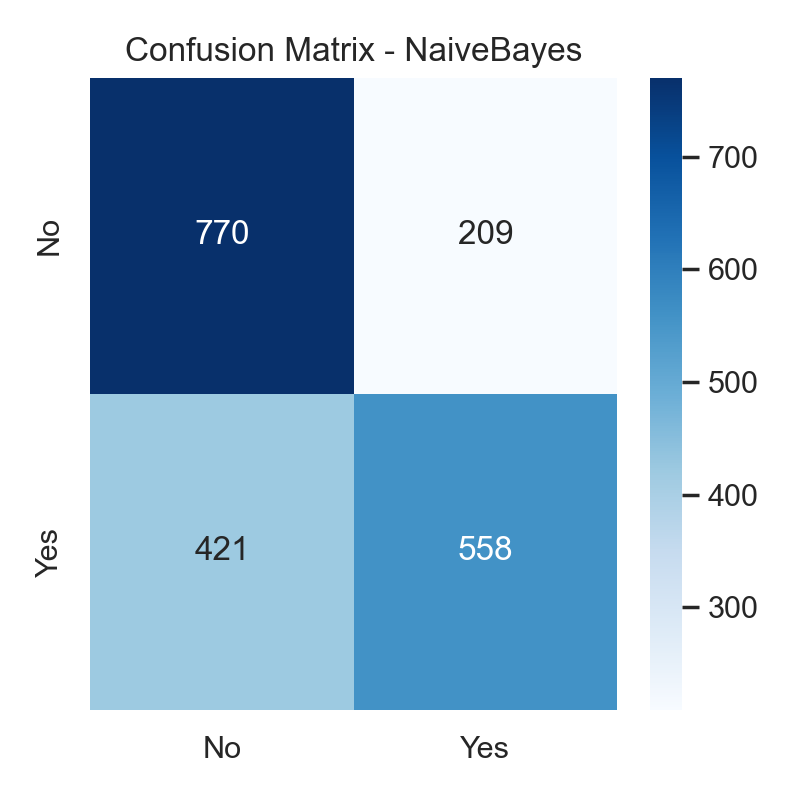

### Fig 12.png

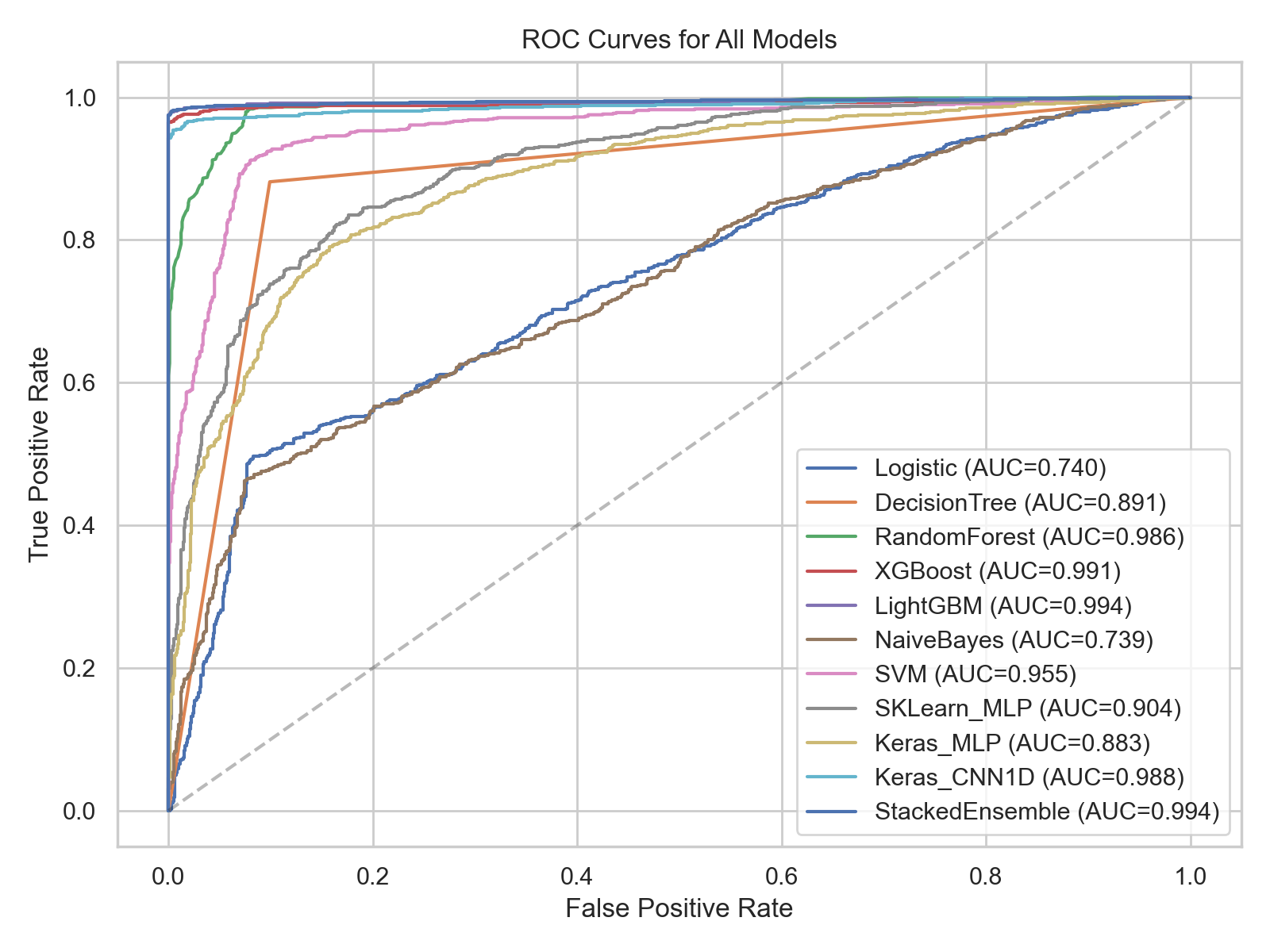

### Fig 13.png

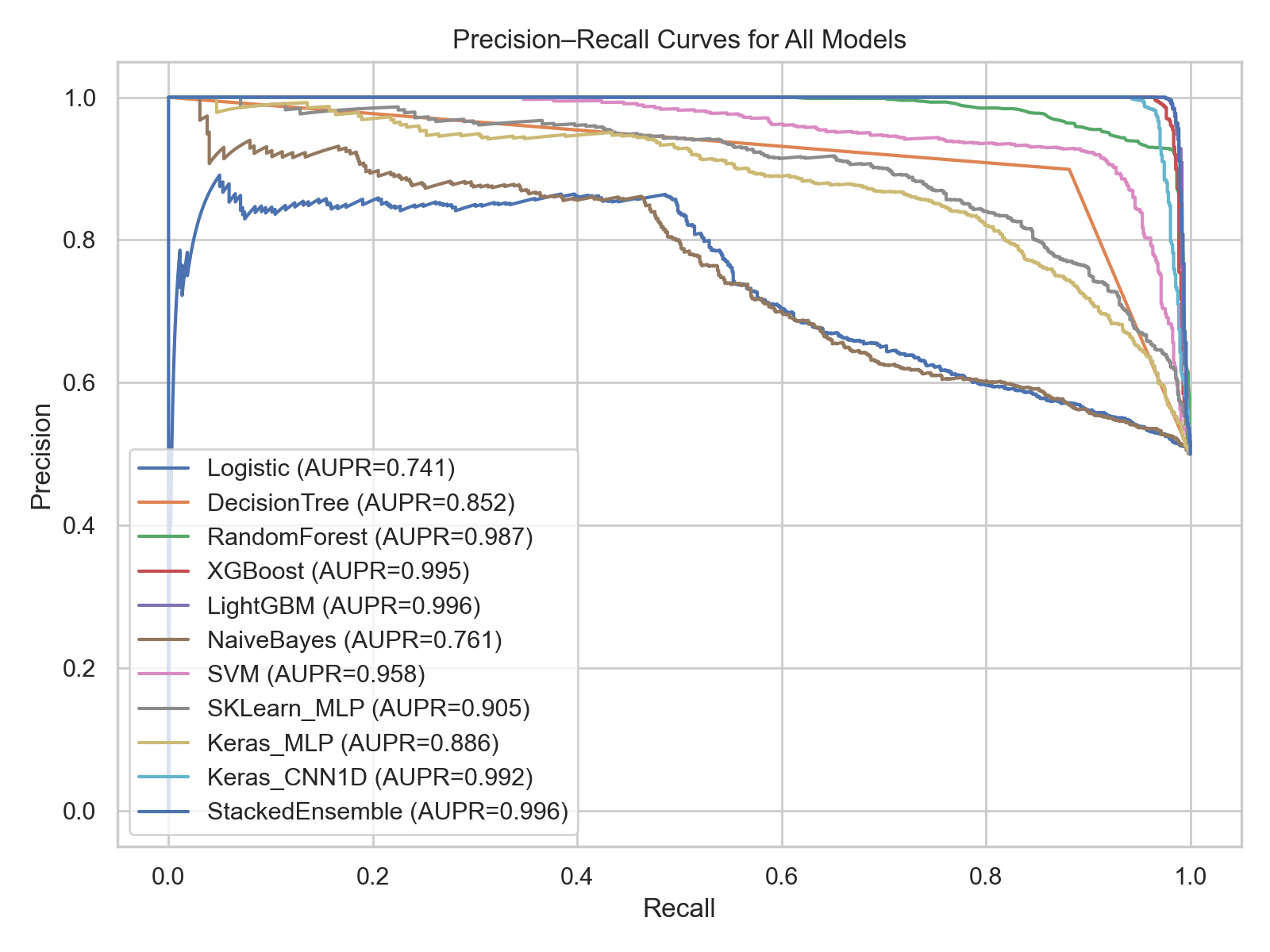

### Fig 14.png

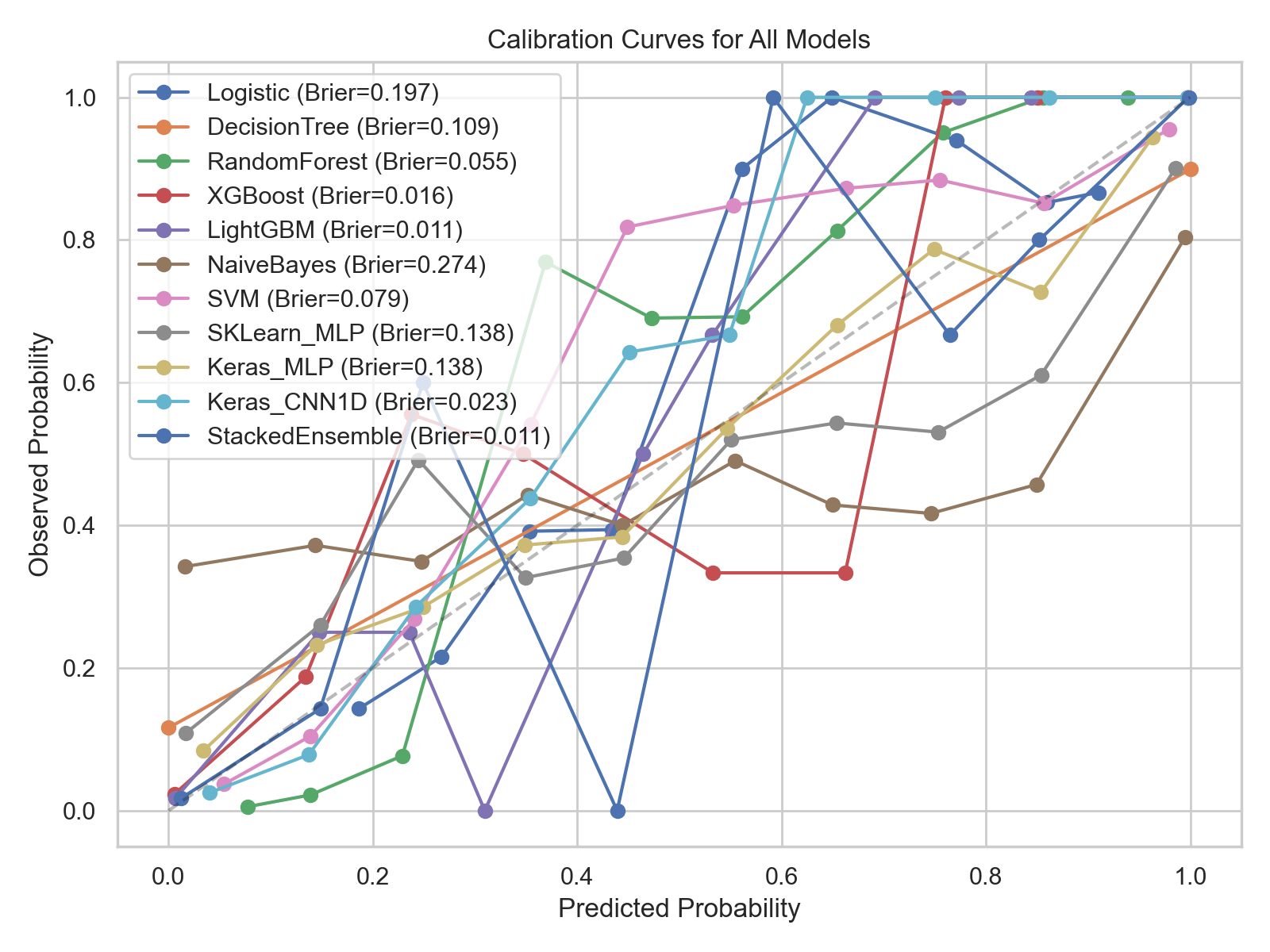

### Fig 15.png

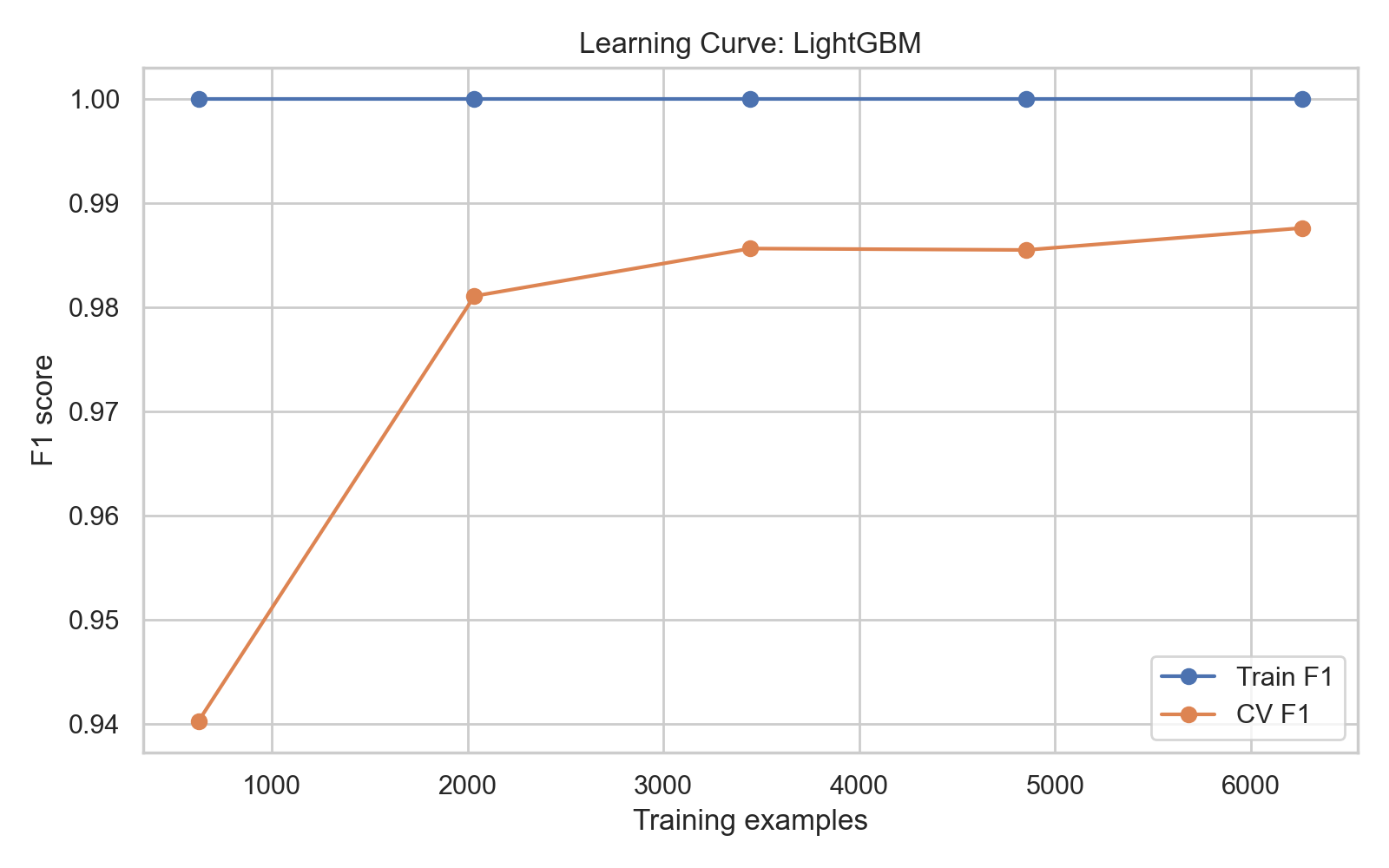

### Fig 16.png

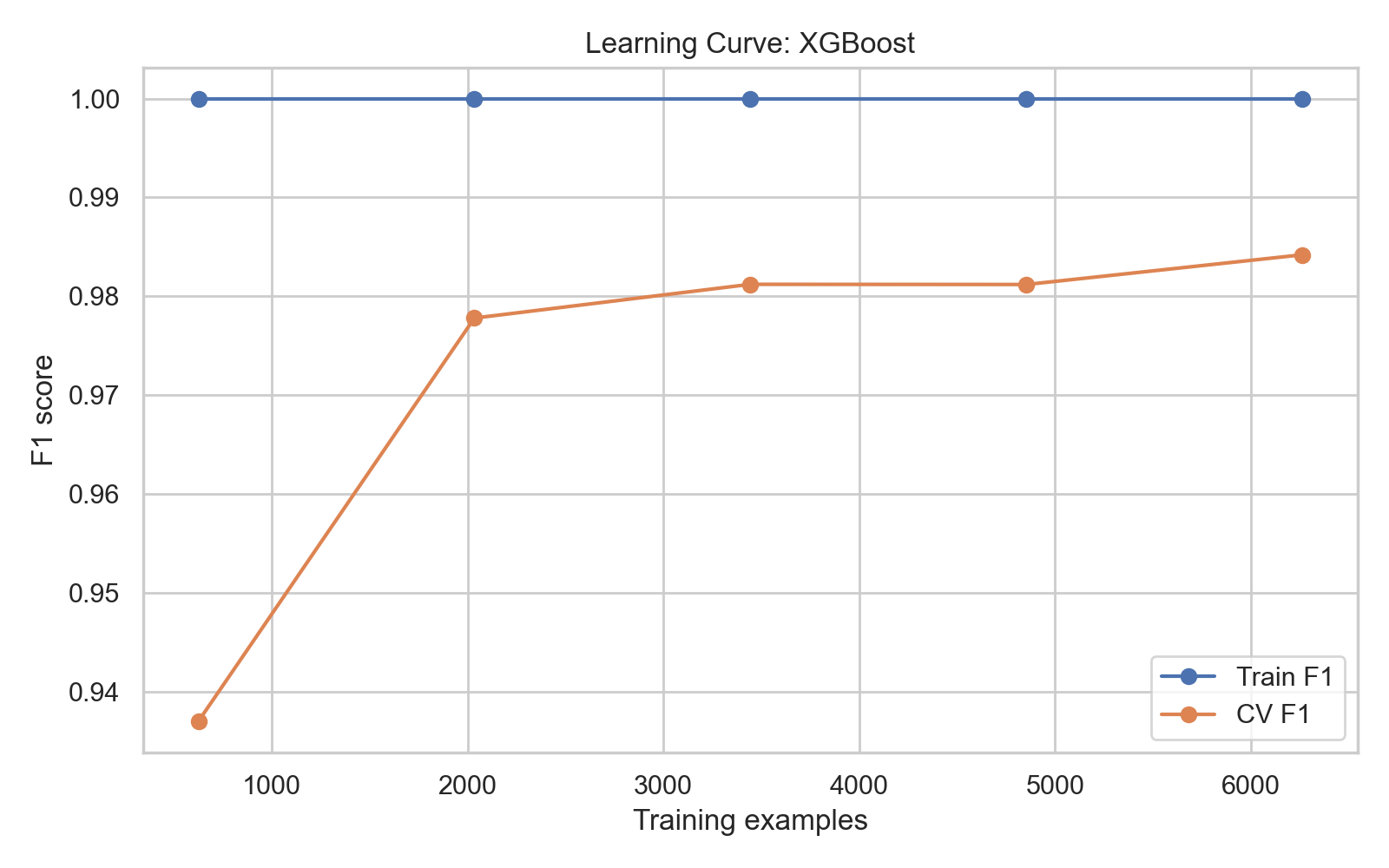

### Fig 17.png

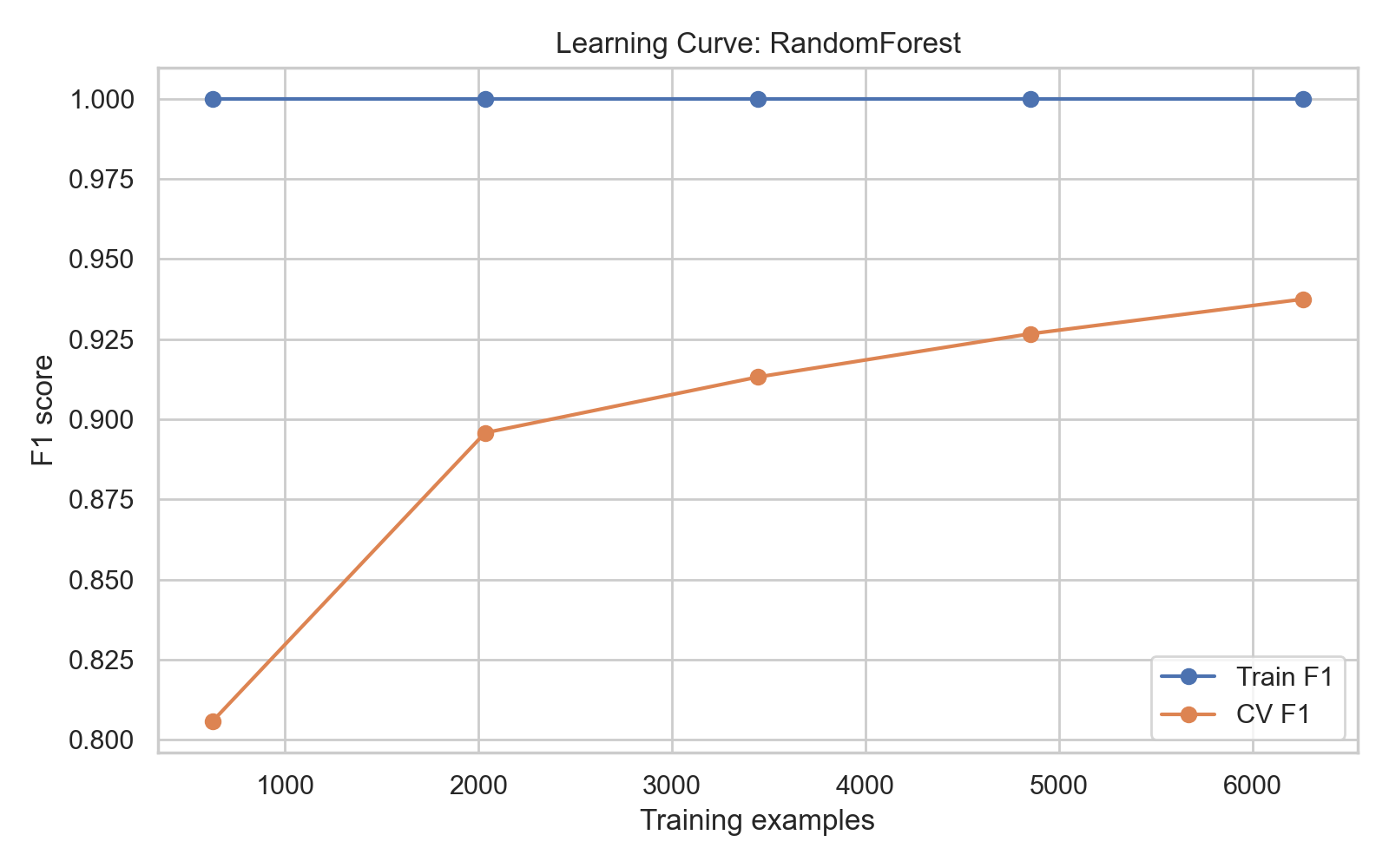

### Fig 18.png

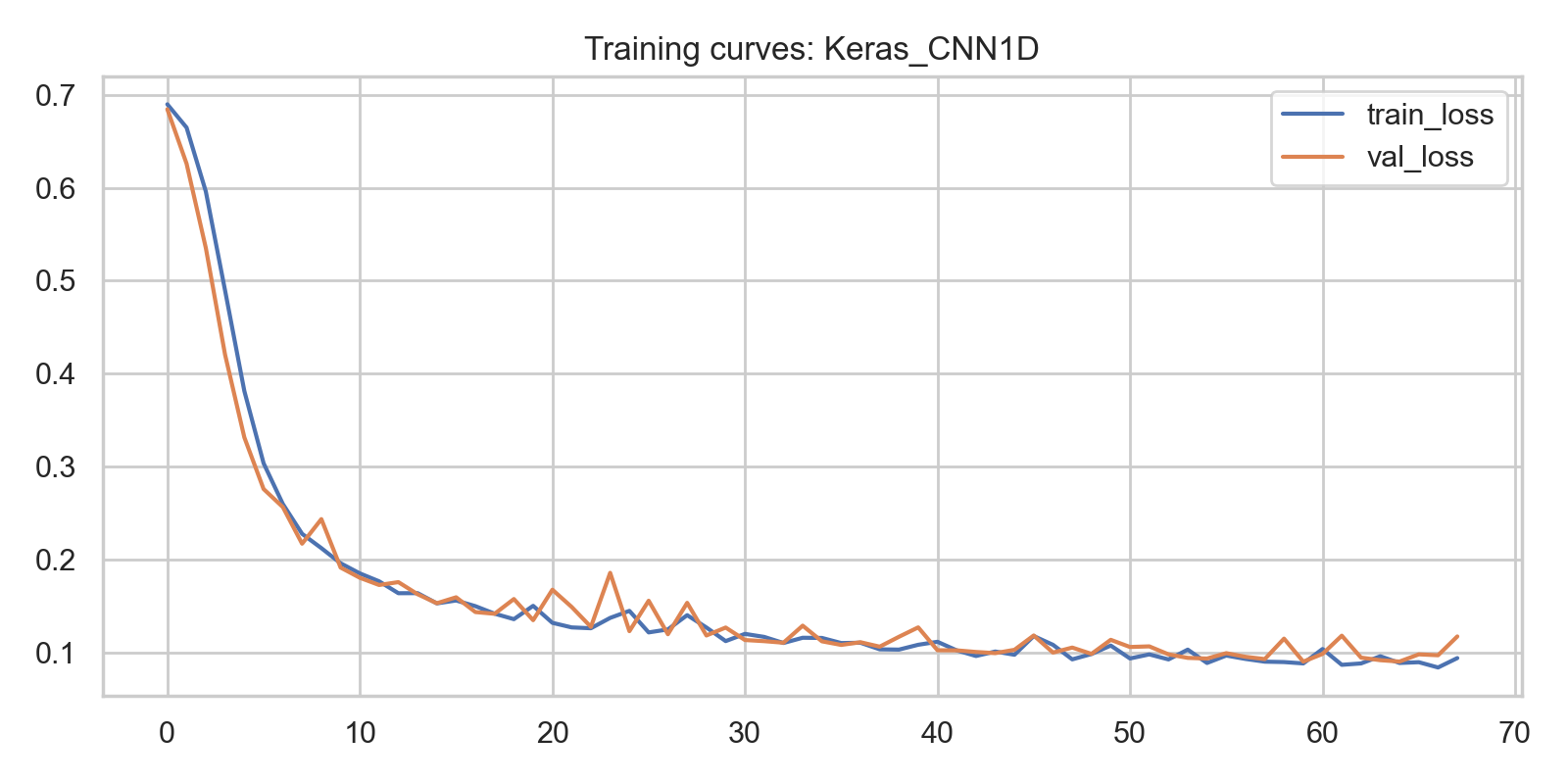

### Fig 19.png

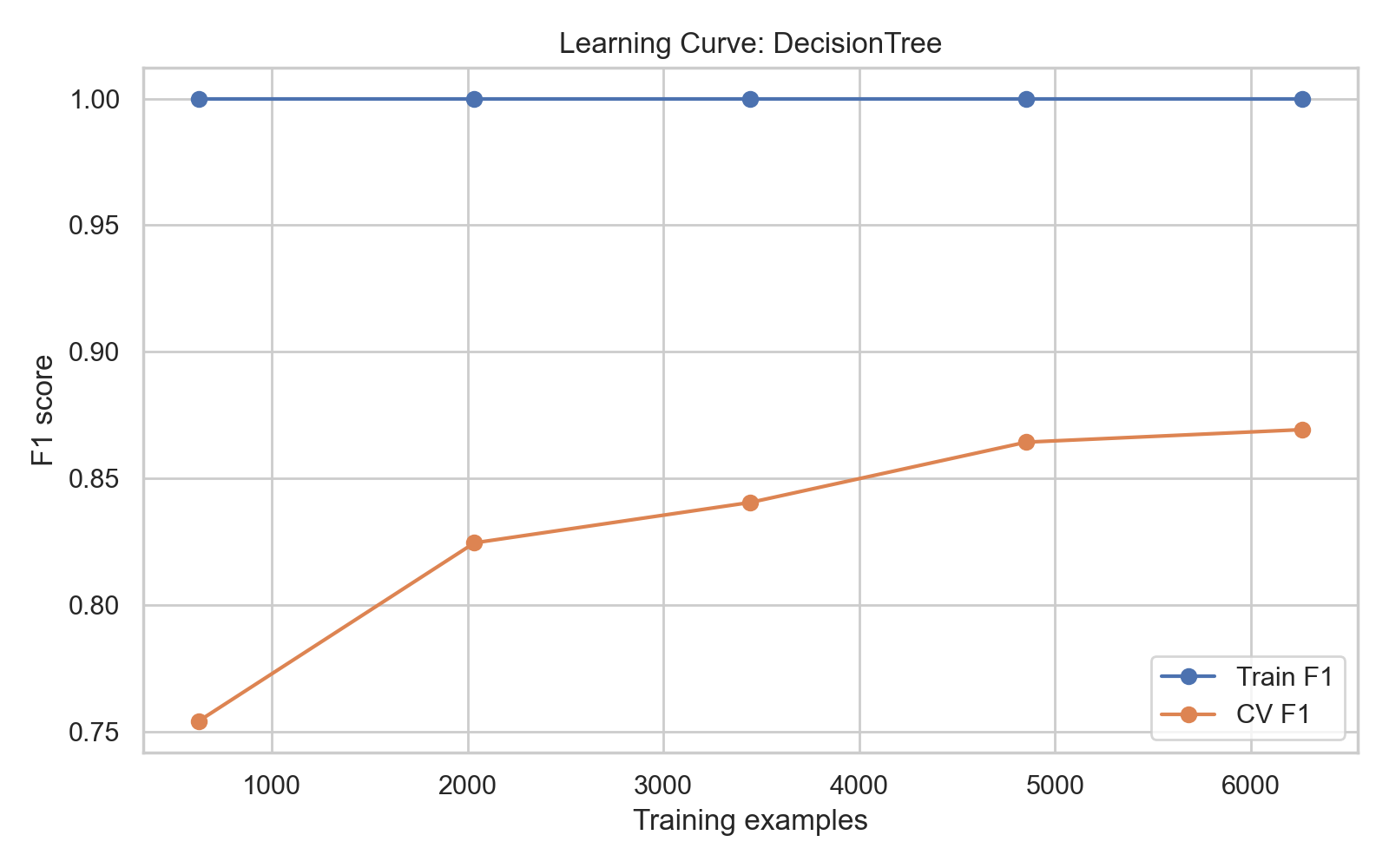

### Fig 20.png

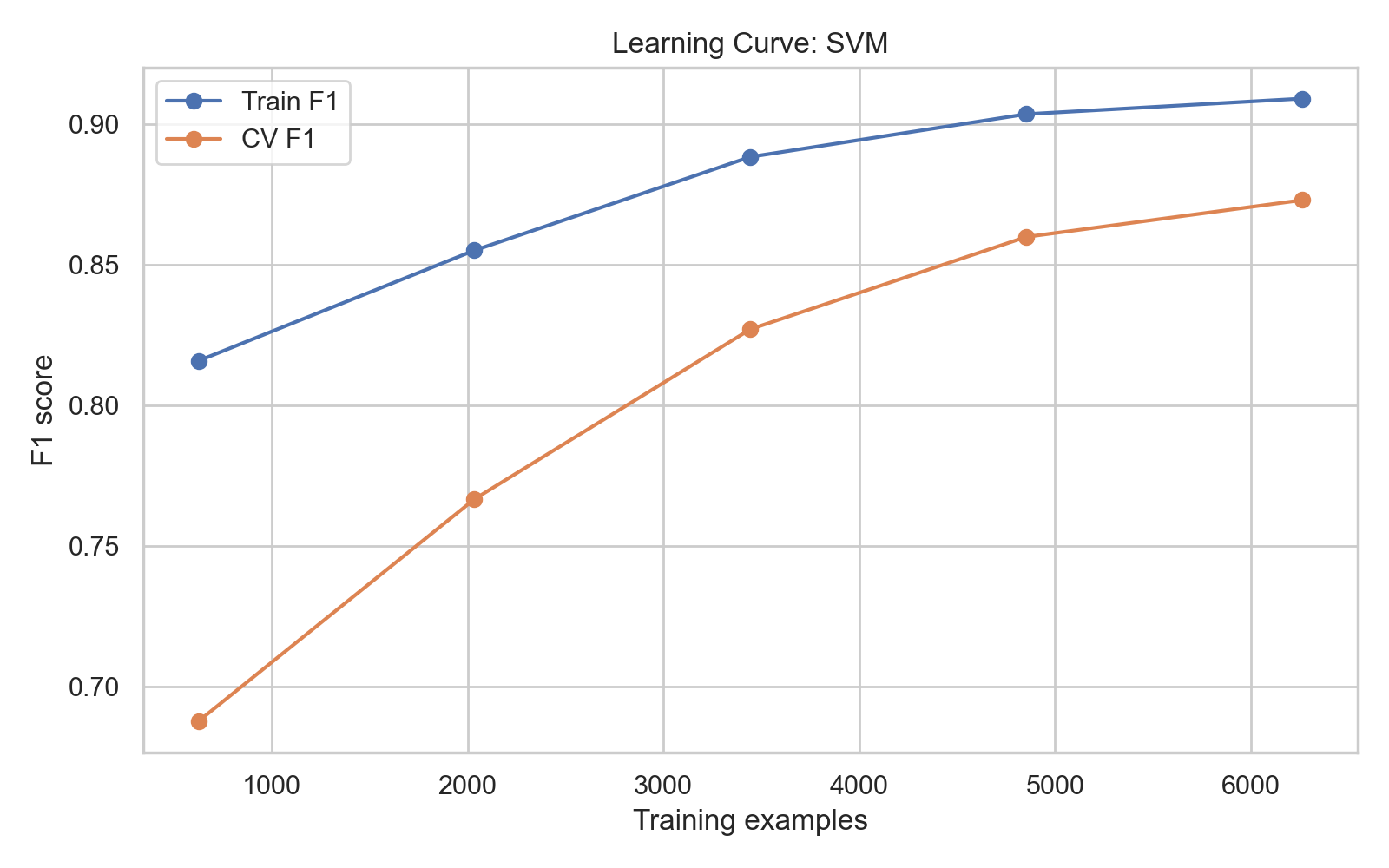

### Fig 21.png

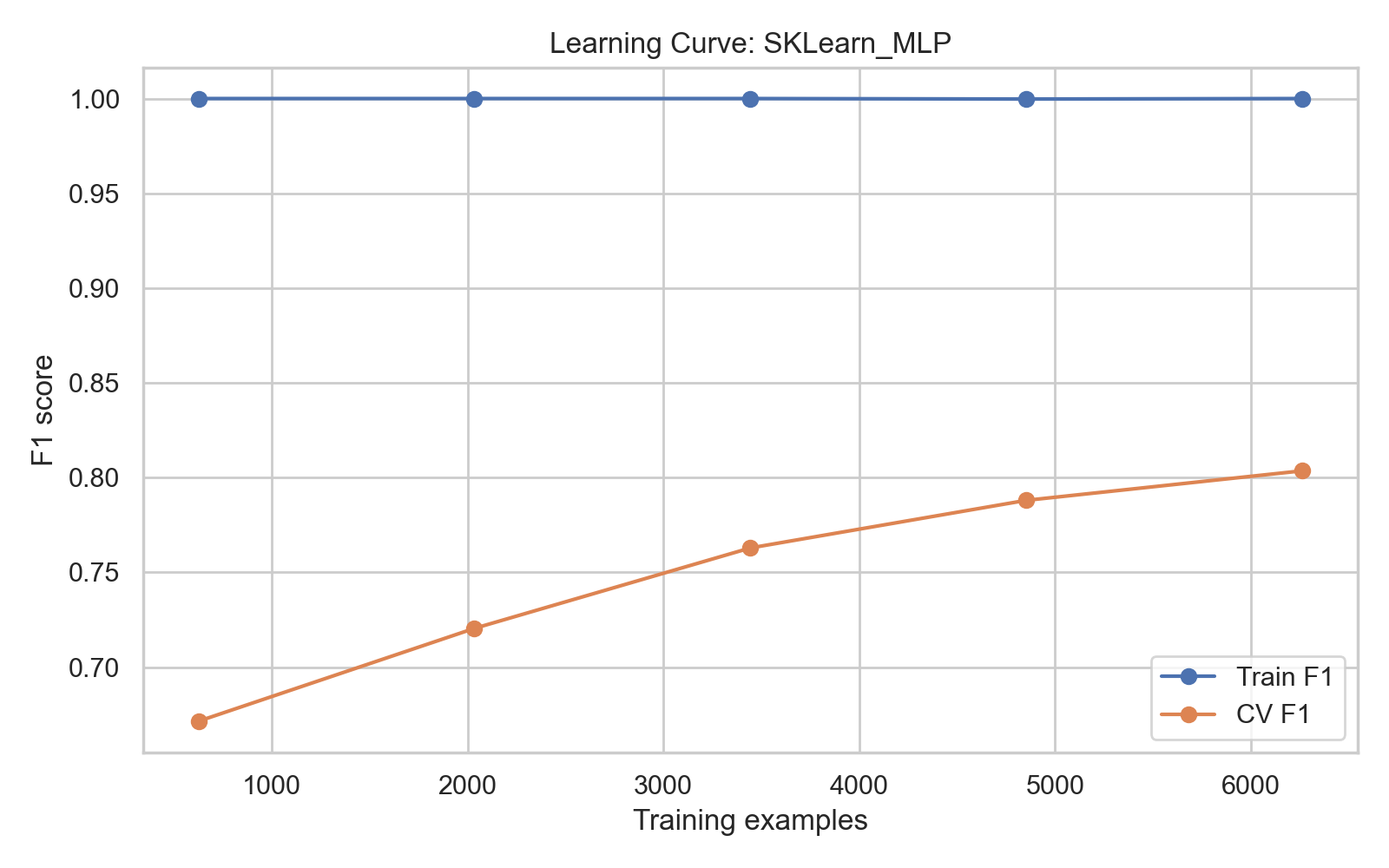

### Fig 22.png

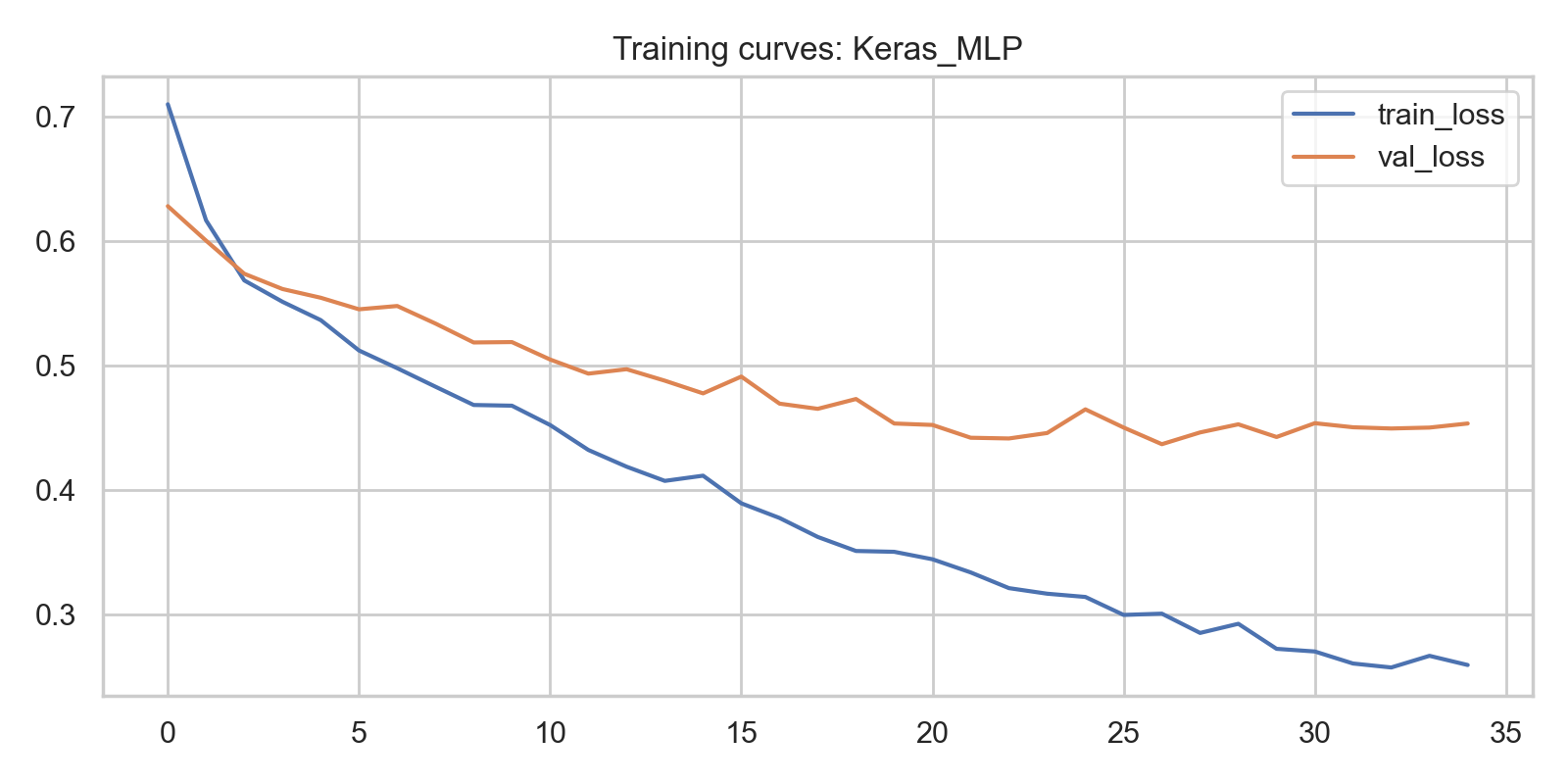

### Fig 23.png

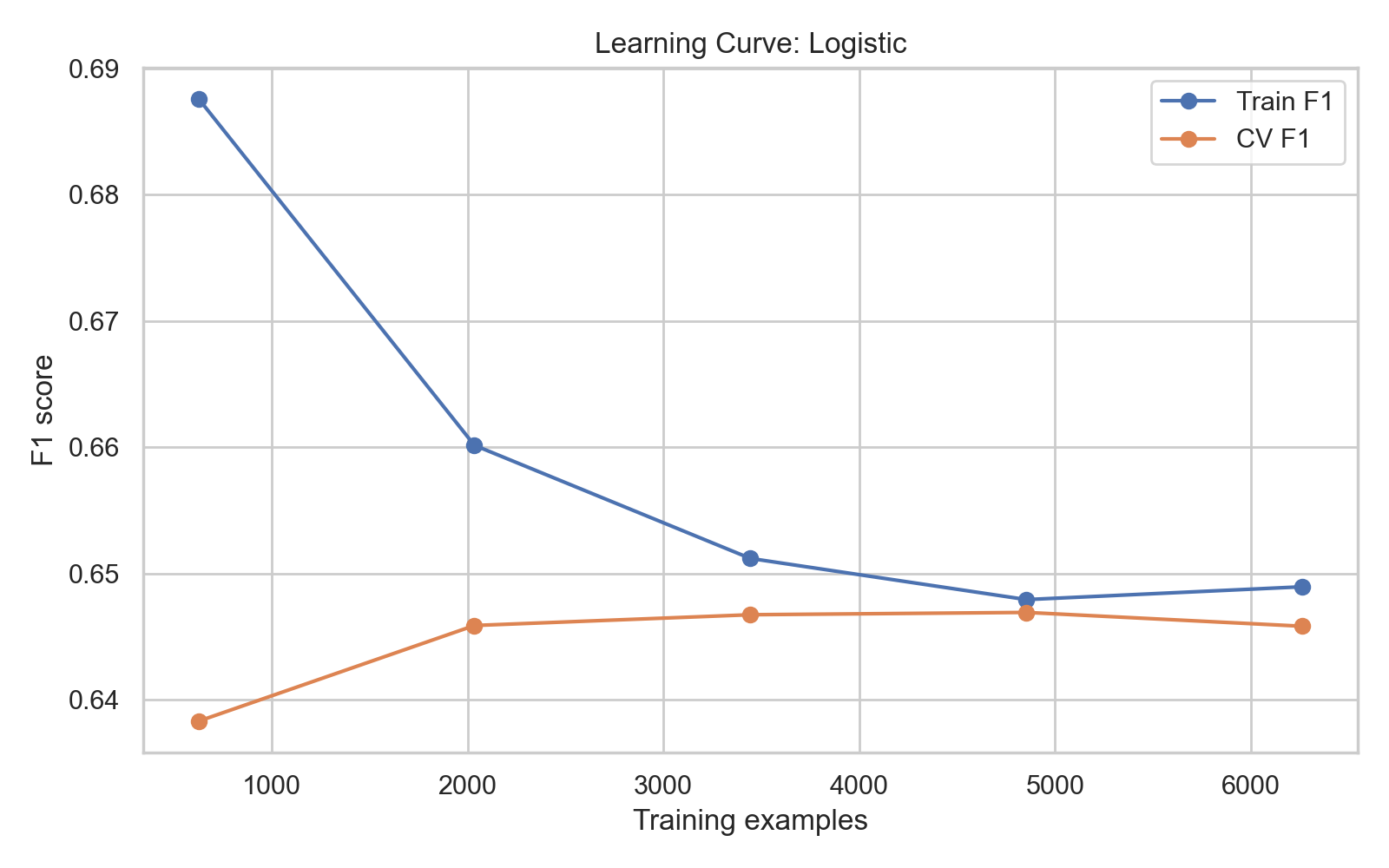

### Fig 24.png

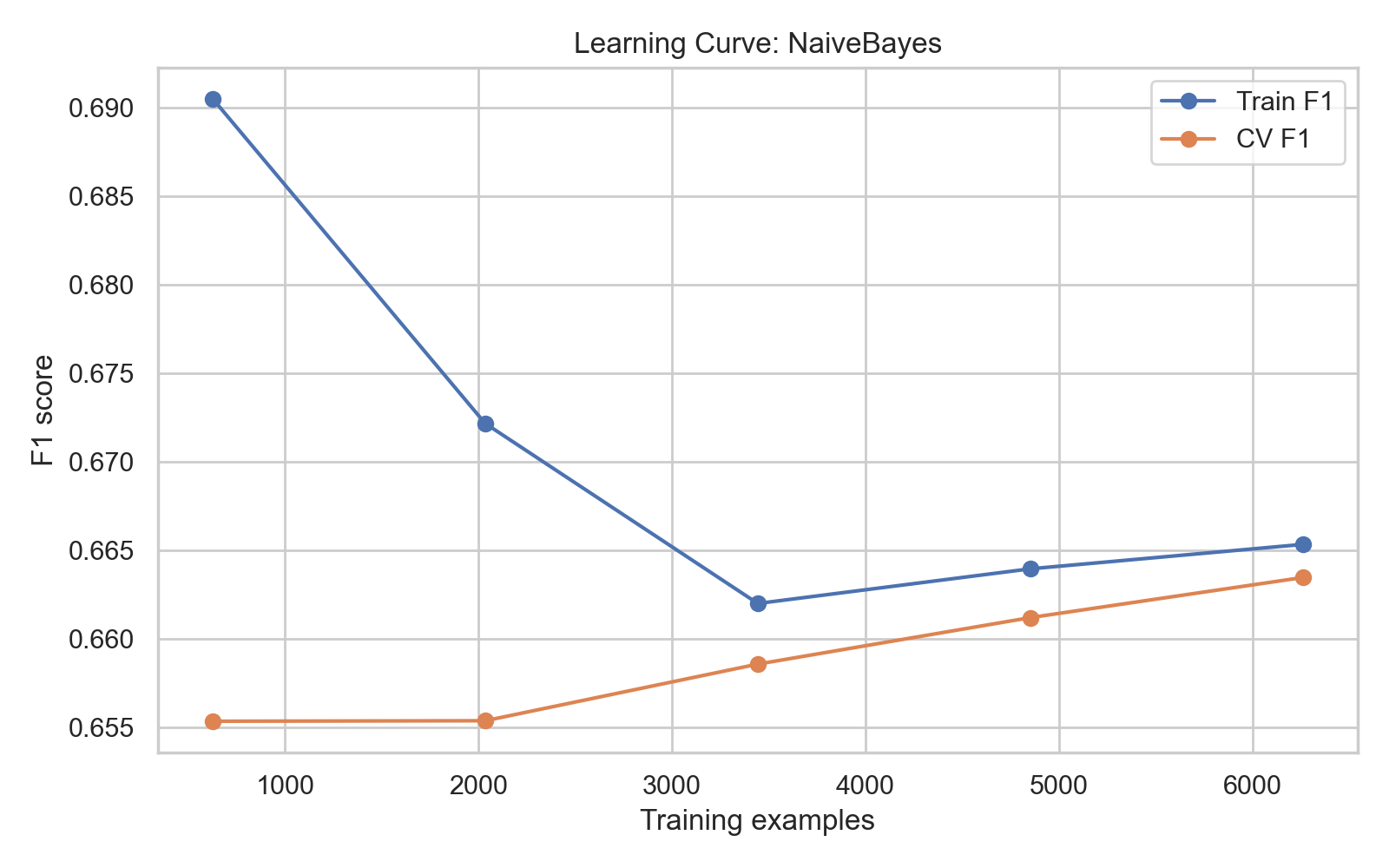

### lime_LightGBM_negative.png

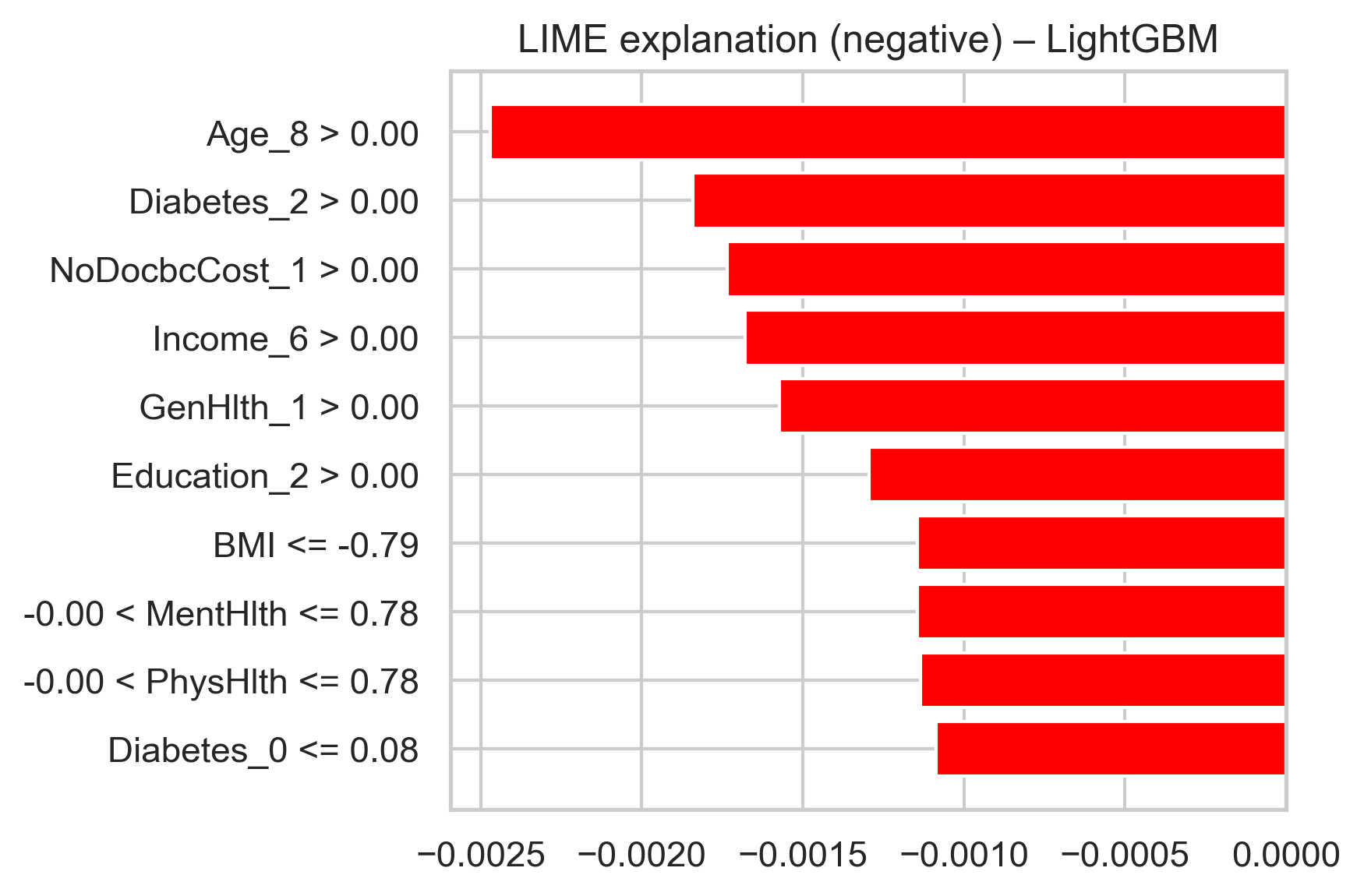

### lime_LightGBM_positive.png

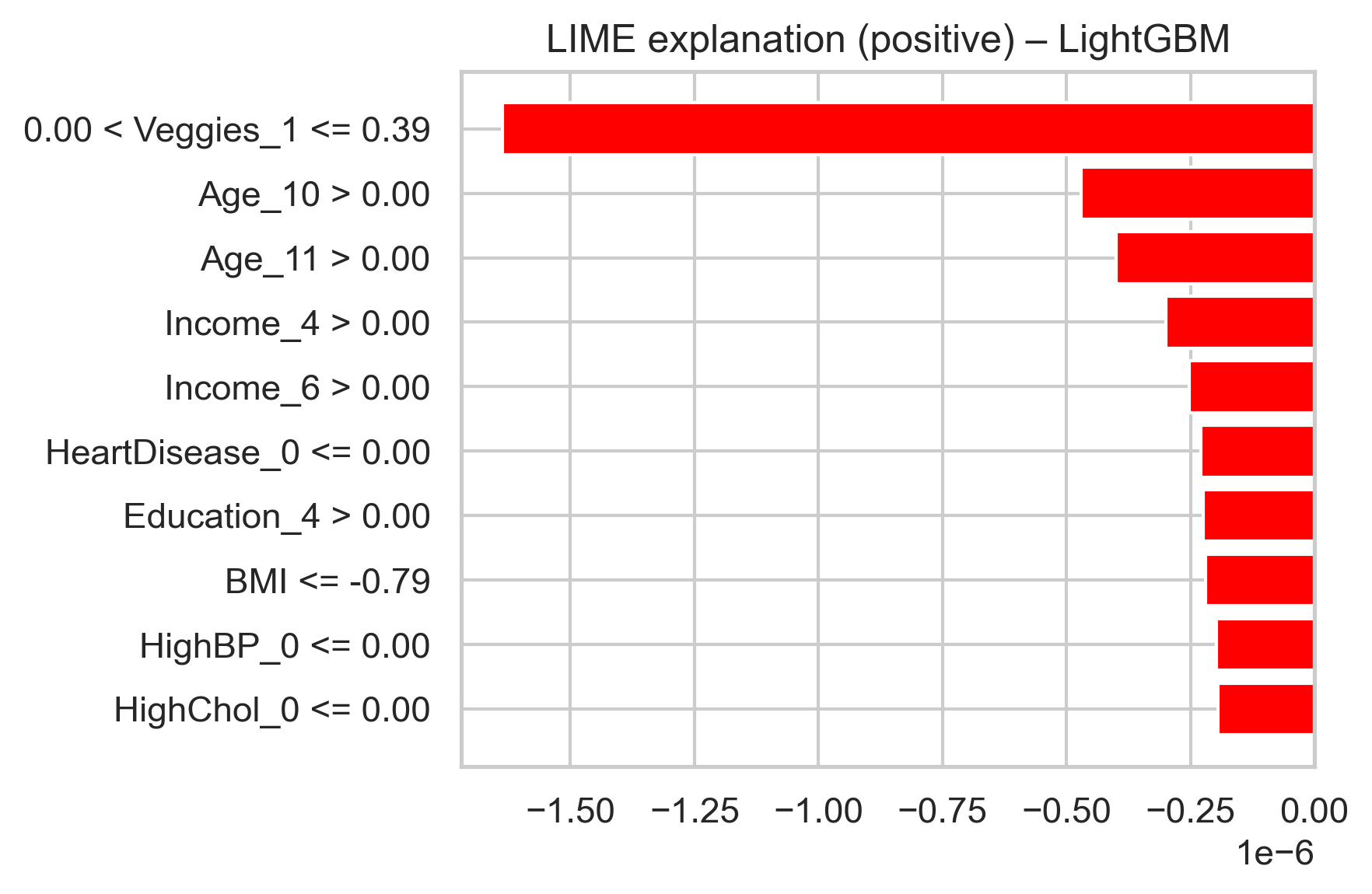

### shap_bar_LightGBM.png

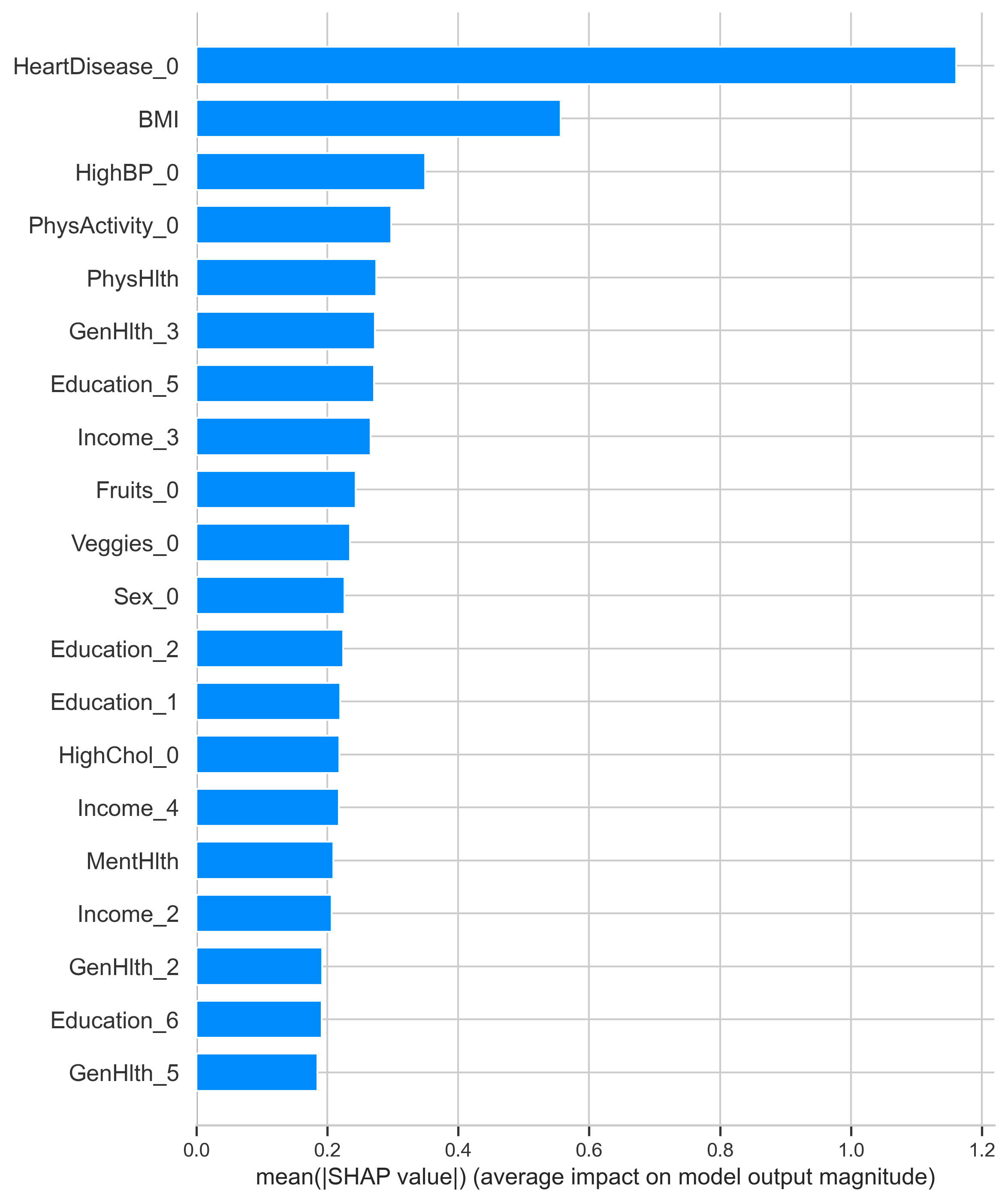

### shap_bar_XGBoost.png

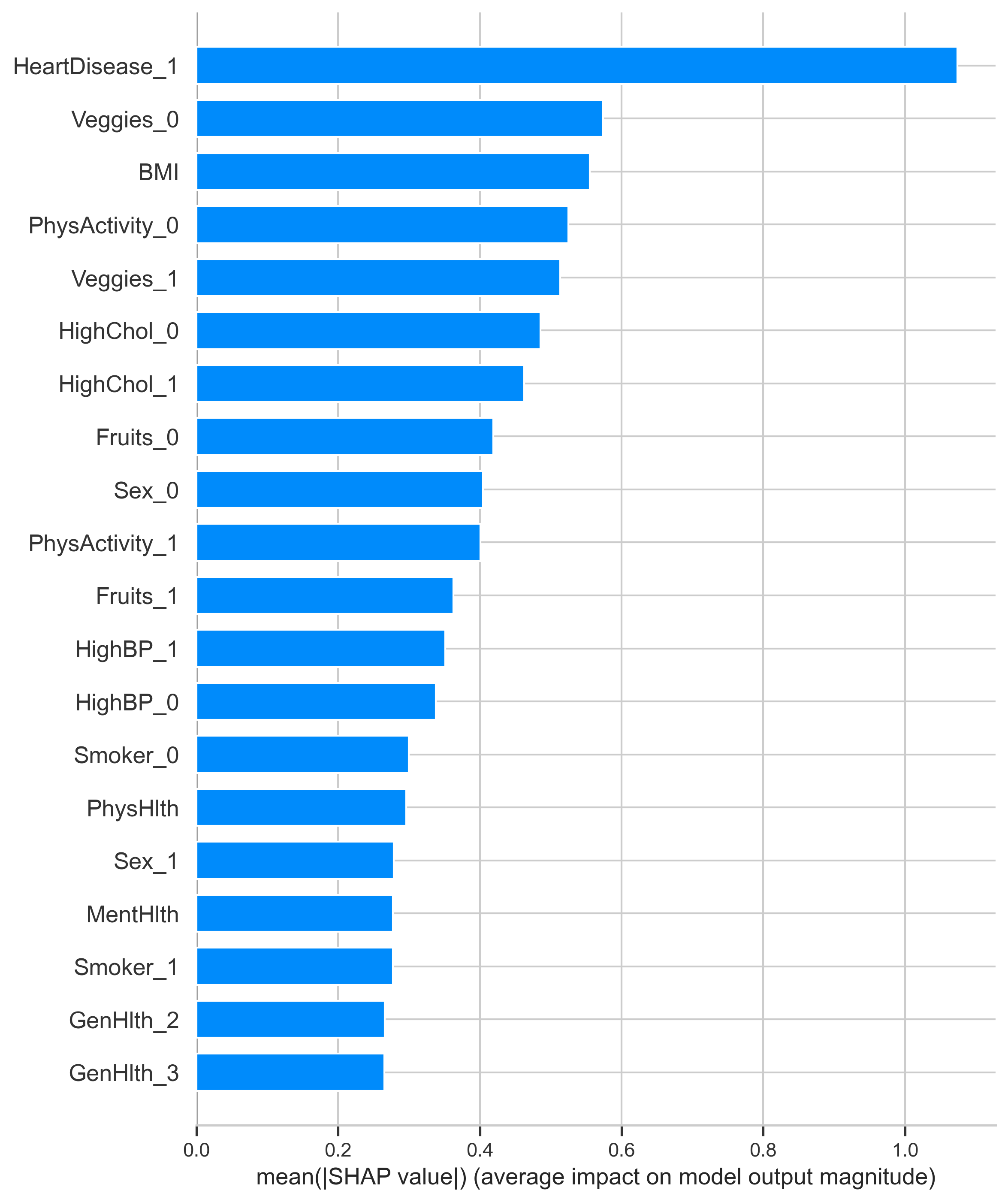

### shap_beeswarm_LightGBM.png

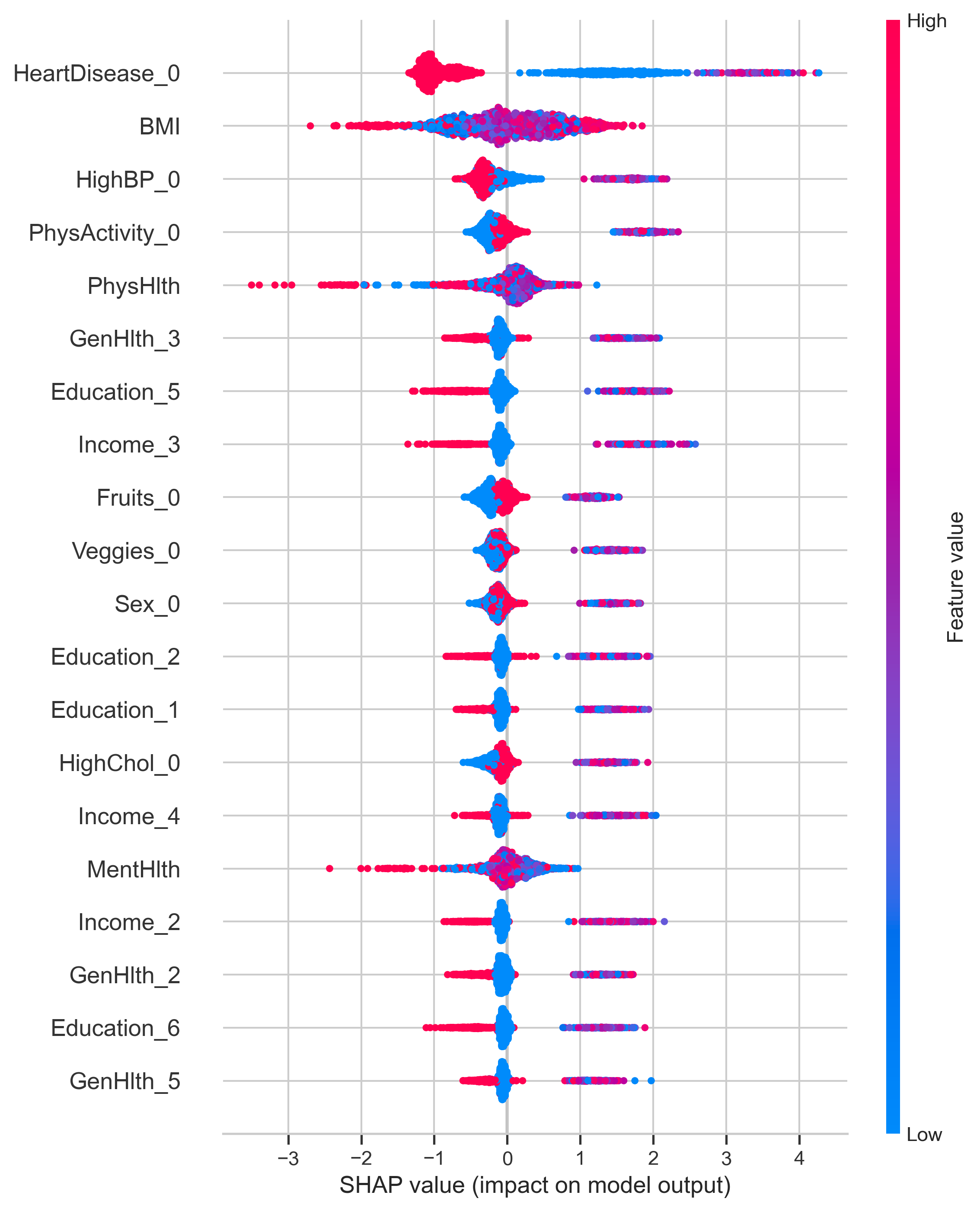

### shap_beeswarm_XGBoost.png

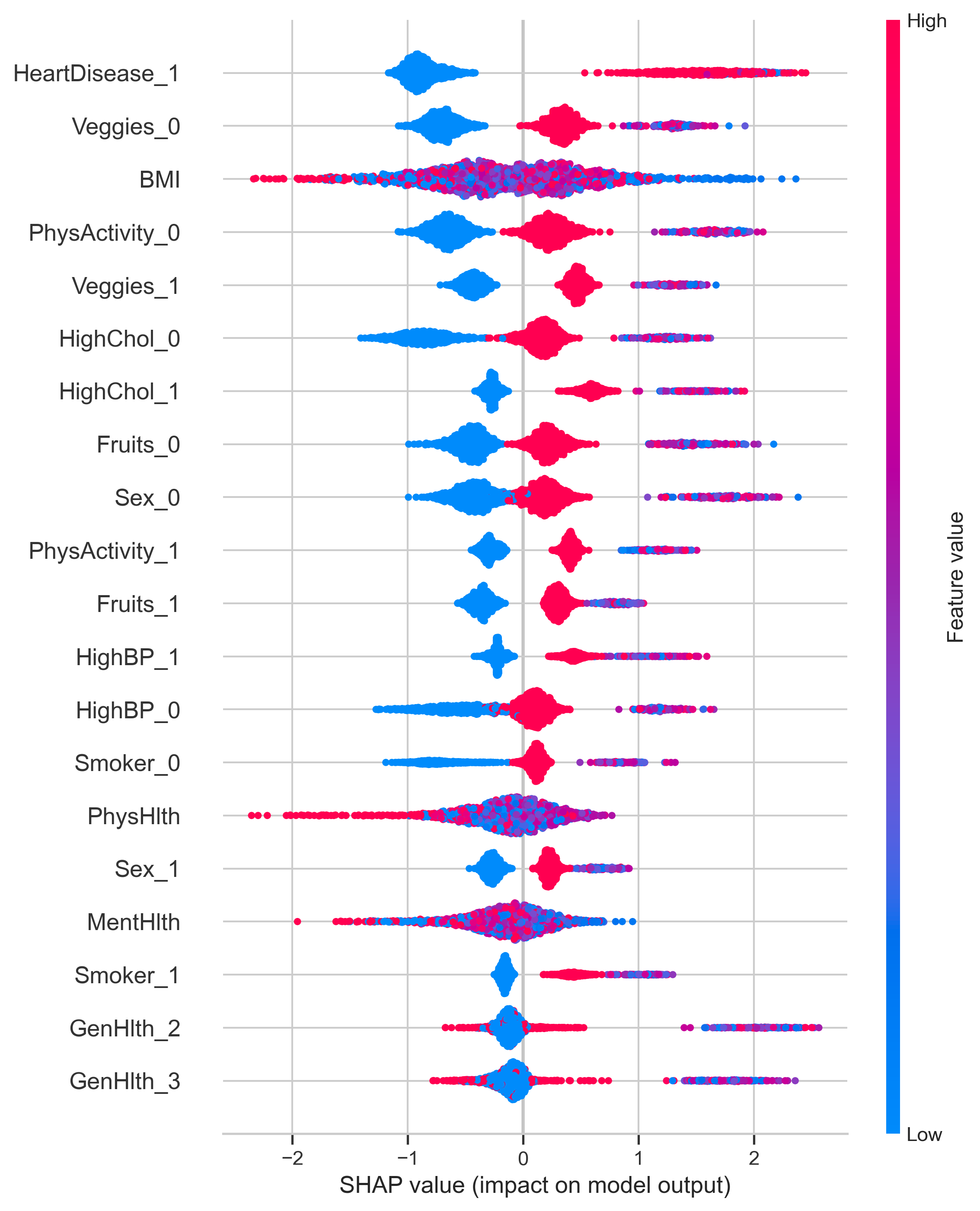

### shap_local_LightGBM_negative.png

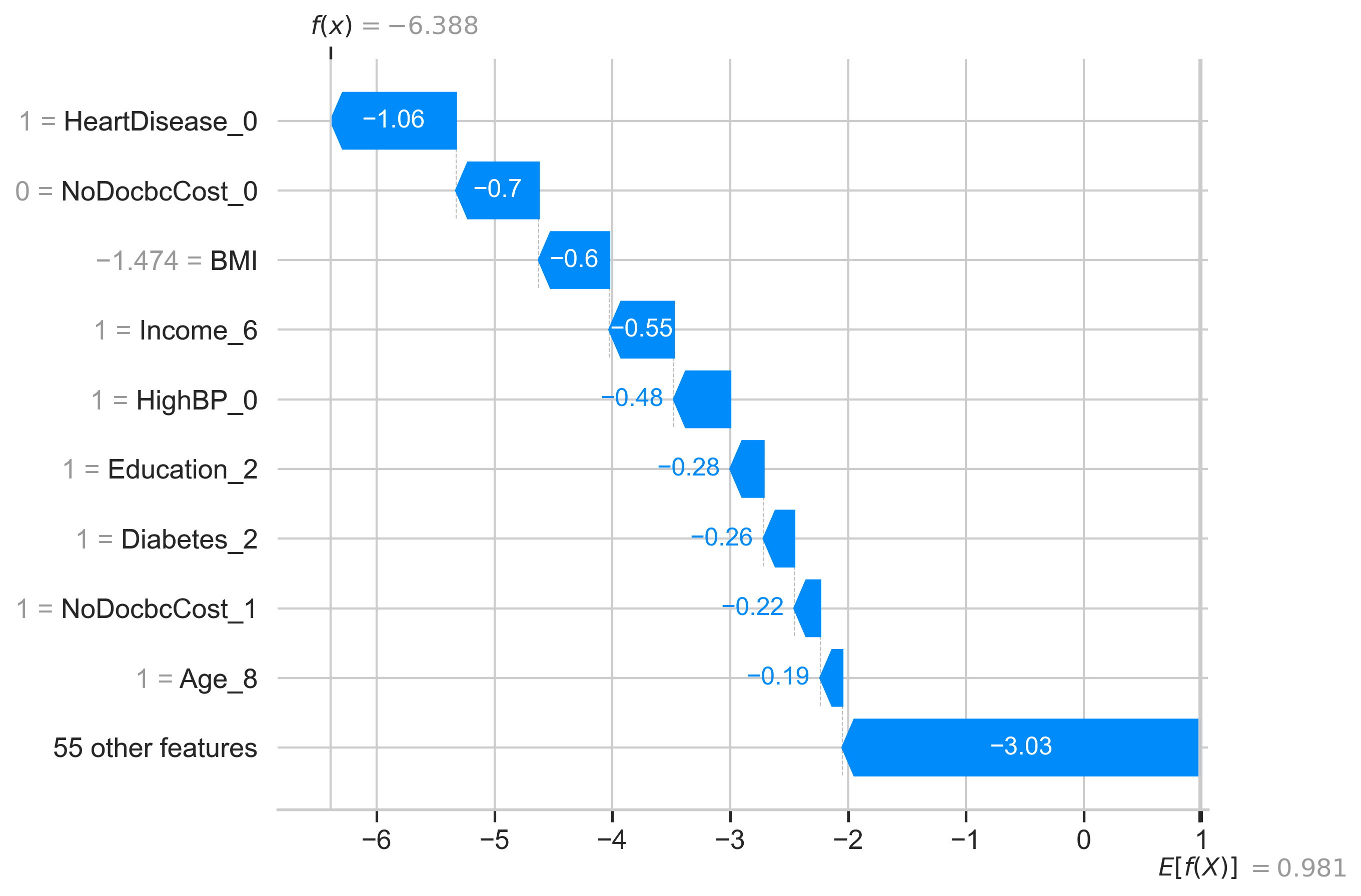

### shap_local_LightGBM_positive.png

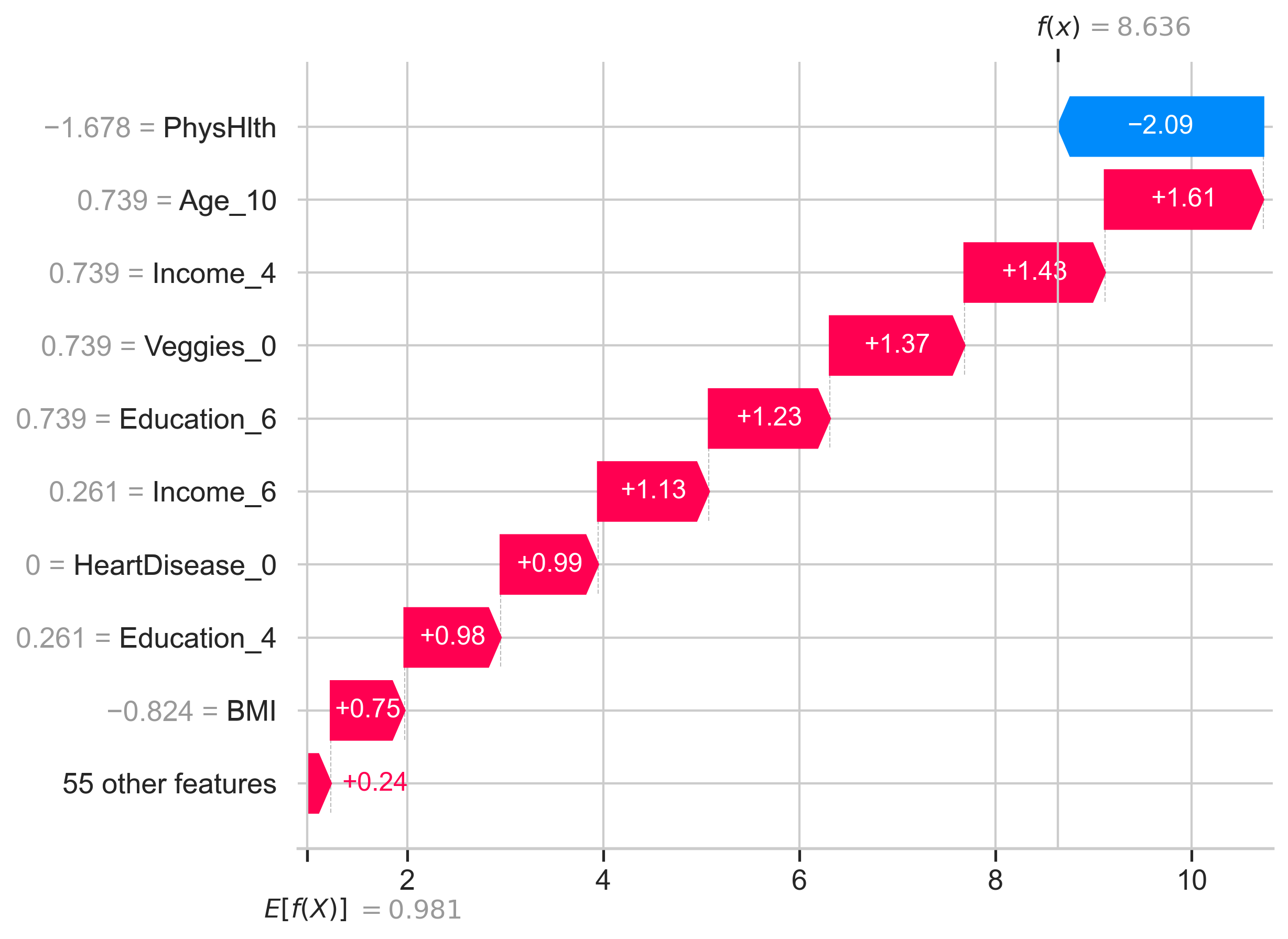

### shap_local_XGBoost_negative.png

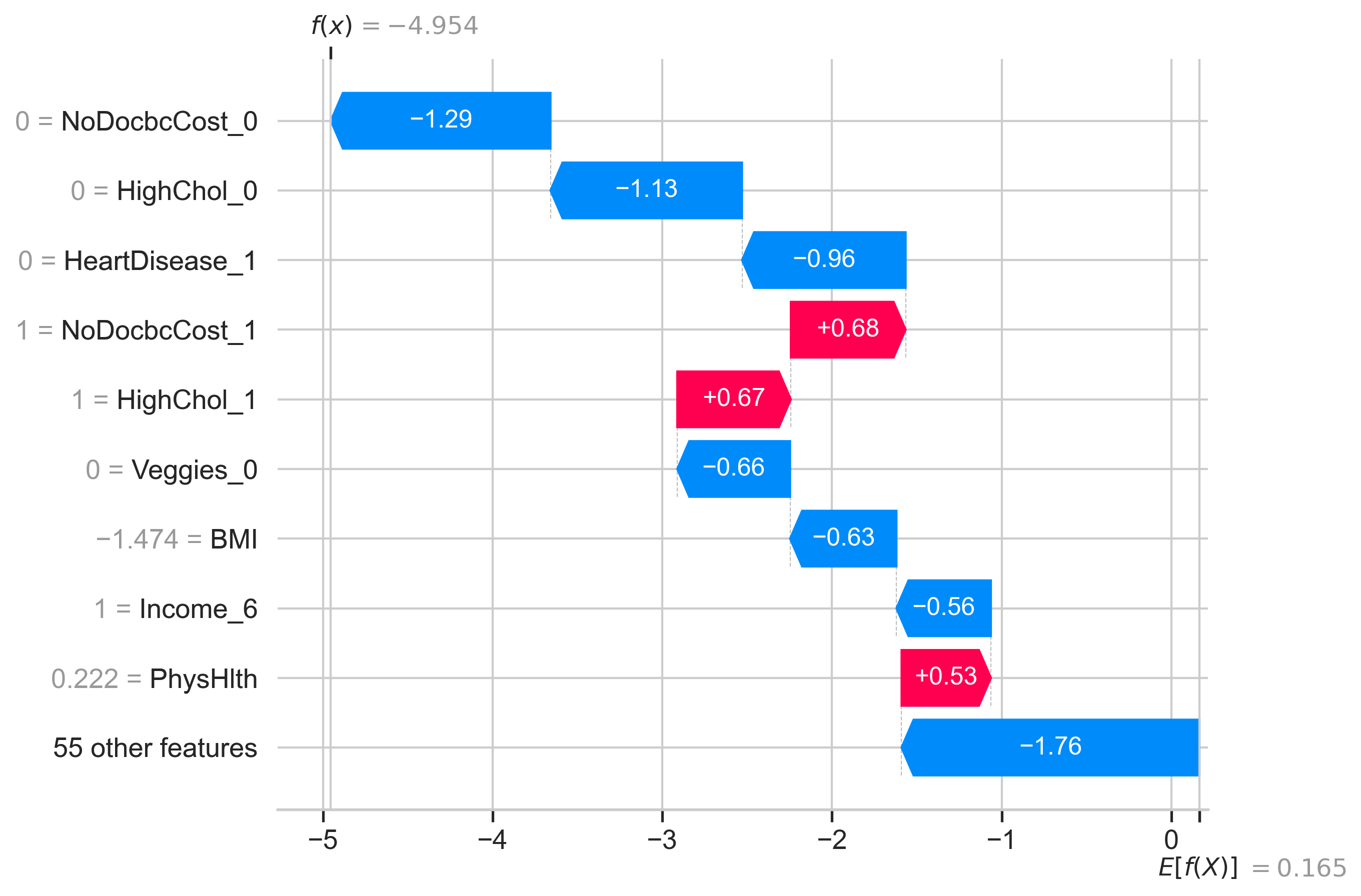

### shap_local_XGBoost_positive.png

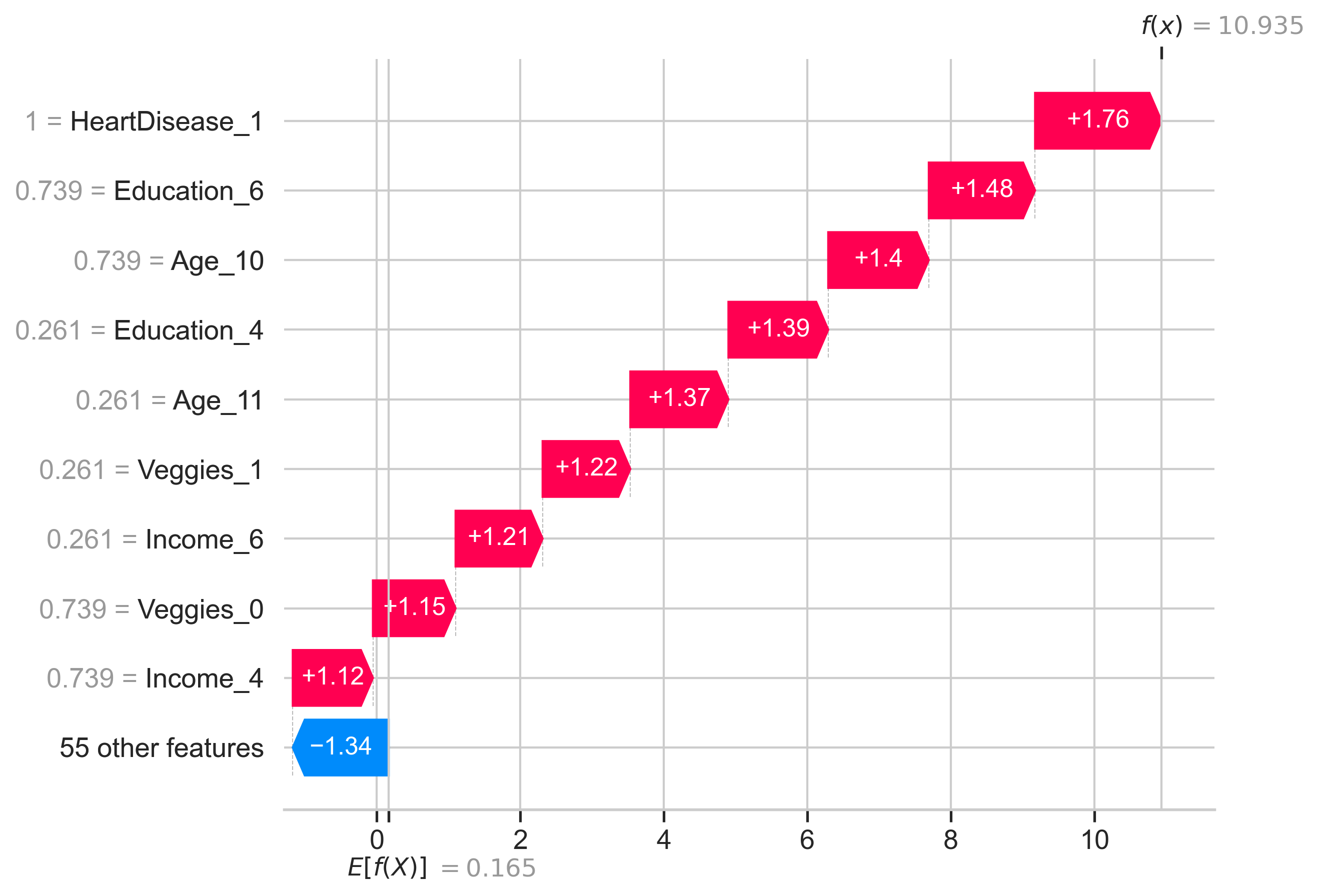

### shap_stacked_bar.png

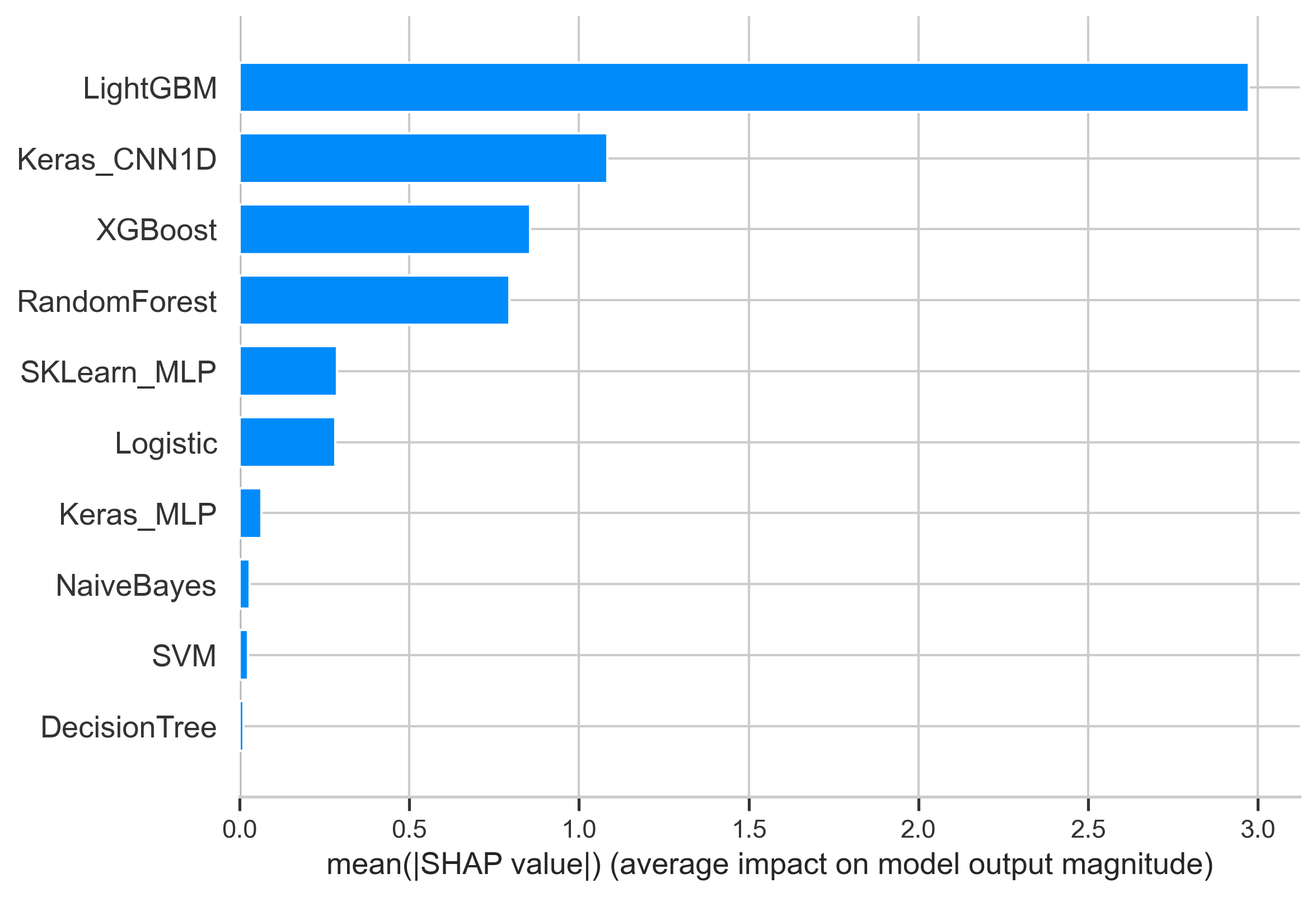

### shap_stacked_beeswarm.png

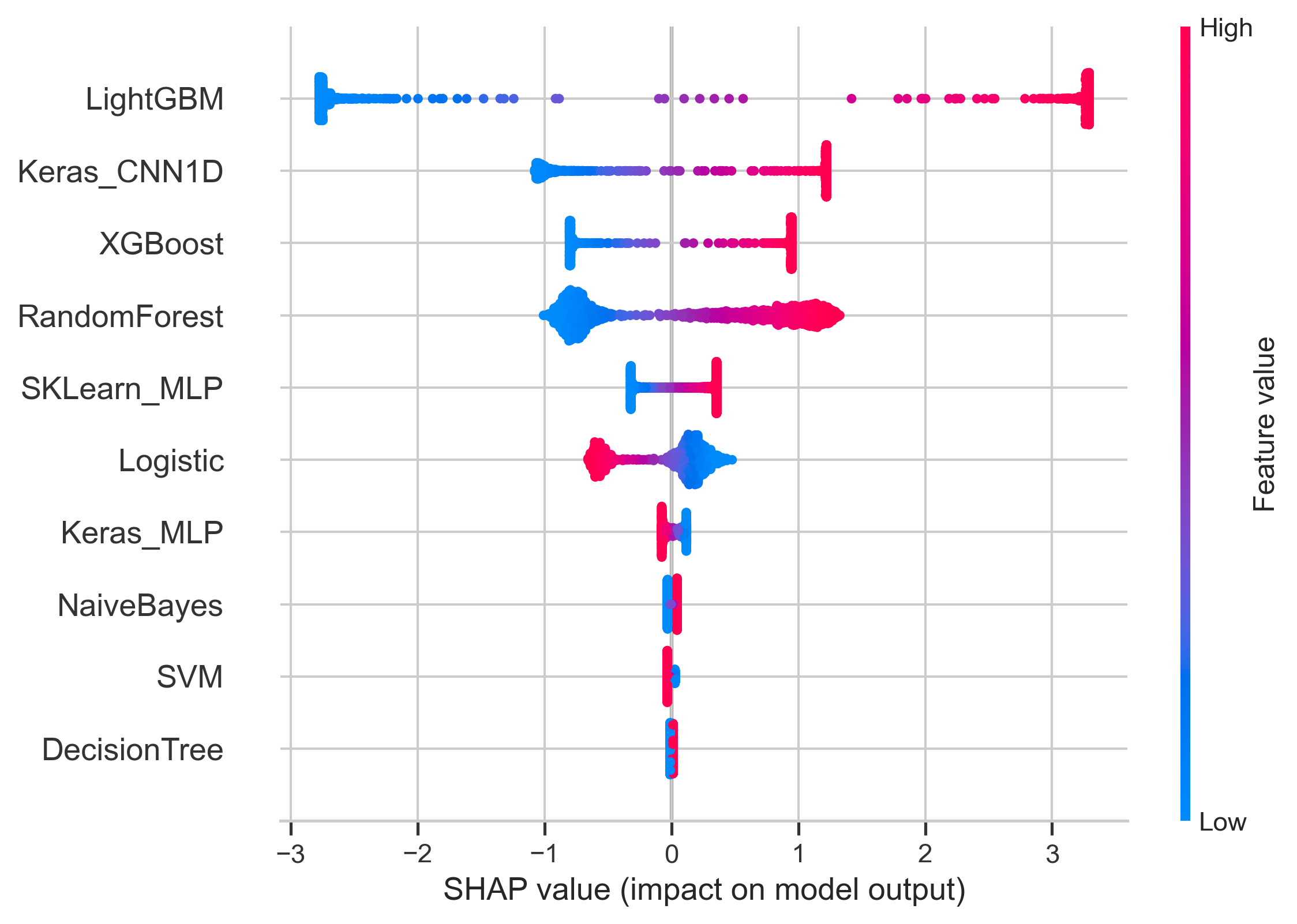
